# Human vs AI Clinical Assessment: Benchmarking a Multimodal Foundation Model Against Multi-Center Expert Judgment on the Mental Status Examination

**DOI:** 10.64898/2026.04.17.26351105

**Authors:** Benson Mwangi, Hammza Jabbar Abd Sattar Hamoudi, Marsal Sanches, Nurhak Dogan, Pooja Chaudhary, Mon-Ju Wu, Giovana B. Zunta-Soares, Jair C. Soares, Andrés Martin, Cesar A. Soutullo

## Abstract

The Mental Status Examination (MSE) is the cornerstone of the psychiatric evaluation, yet validating artificial intelligence (AI) against the inherent variance of clinical judgment remains a critical bottleneck. Here we introduce a multi-center framework to benchmark the open-weight multimodal foundation model Qwen3-Omni against independent expert panels at two sites, UTHealth and Yale. Evaluating 396 classifications across 10 MSE domains and three longitudinal timepoints of increasing symptom severity, we found that experts achieved substantial agreement (Gwet’s AC1 = 0.87), whereas the model achieved only moderate alignment (AC1 = 0.70-0.72). Even as the model’s overall pathology prediction rate approximated the experts’, the aggregate equilibrium masked a profound “clinical reasoning gap”. Specifically, the model systematically over-predicted observable signs (e.g., speech, affect) while notably failing in inferential domains requiring the interpretation of latent mental content (e.g., delusions, perceptions). A 4-bit quantization analysis of the model confirmed this mechanistically: reducing model capacity disproportionately degraded inferential reasoning while preserving perceptual feature extraction. Furthermore, model-to-expert agreement degraded linearly as clinical complexity intensified across longitudinal visits (Accuracy: T0 = 84.8-87%; T1 = 80-82%; T2 = 71-73%), whereas expert consensus remained robust. Notably, model errors increased 2.3-to-3.4 fold where human experts disagreed. These findings establish inter-expert variance as an essential measurable baseline for psychiatric AI, demonstrating that true clinical translation requires models to move beyond multimodal perceptual extraction to achieve higher-order diagnostic reasoning.

## Introduction

The mental status examination (MSE) is the foundational framework through which psychiatrists observe, interpret, and classify a patient’s behavioral and cognitive functioning (Voss and Das 2026; Strub and Black 1993). Despite its centrality to clinical practice, the MSE exhibits substantial inter-rater variability, particularly in domains requiring the subjective inference of latent mental content, such as mood, thoughts, and perceptions (Blaabjerg et al. 2020). These reliability challenges echo broader historical difficulties in psychiatric diagnostics (Regier et al. 2013). Beyond individual expert variance, there is profound structural heterogeneity in how the MSE itself is organized and how its core clinical constructs are conceptualized across both English and international literature (Daza et al. 2025; Sanches 2025). Crucially, however, this variability is not merely statistical noise: It reflects the genuine, domain-specific ambiguity inherent to psychiatric phenomena, constituting a fundamental and measurable property of the clinical examination itself.

Recognizing expert variance as a clinical reality carries profound implications for the development and validation of artificial intelligence (AI) in mental health. Unlike laboratory assays or structural imaging that yield objective and quantifiable metrics, the MSE relies entirely on the real-time human synthesis of complex behavioral signals. Consequently, evaluating any AI-driven diagnostic tool against a single expert panel treating that singular perspective as an infallible “ground truth” is methodologically flawed. A model’s clinical utility must instead be calibrated against the natural distribution of expert consensus, ideally under controlled conditions where diagnostic profiles are known, and symptom progression is rigorously calibrated. A robust evaluation framework must therefore simultaneously benchmark model-to-expert agreement against baseline inter-expert agreement, utilizing prevalence-adjusted statistics to ensure stability against the class imbalances typical of psychiatric datasets. This methodological imperative forms the foundational premise of the present study.

Despite substantial progress in applying AI to mental health ranging from automated depression screening to psychiatric disorder classification (Jiang et al. 2024; Sahili et al. 2025), a critical translational gap remains. Existing paradigms predominantly focus on predicting continuous scores from self-report instruments, such as the PHQ-9 or GAD-7 (Grimm et al. 2025; Teferra and Rose 2023; Wanderley Espinola et al. 2022). The MSE, however, imposes a fundamentally different and compounding cognitive burden. It demands the synchronized integration of visual, auditory, and linguistic streams to classify phenomena across multiple domains simultaneously. Furthermore, it requires clinical reasoning that transcends basic perceptual feature extraction where the model must seamlessly navigate domains anchored in directly observable behavior (e.g., speech, psychomotor activity) alongside domains requiring the extraction of latent mental content from patient narratives (e.g., delusions, obsessions). This dual demand for perceptual synthesis and clinical inference distinguishes the MSE from the single-instrument prediction tasks that have historically dominated psychiatric AI (Librenza-Garcia et al. 2017; Hansen et al. 2025; Sun et al. 2025; Liu et al. 2022). To date, no published study has evaluated a multimodal model’s capacity to generate a full-spectrum, domain-level MSE and benchmarked those outputs against structured expert consensus.

Multimodal foundation models (MFMs) represent a technological convergence that makes this rigorous evaluation possible for the first time. Text-only large language models have demonstrated impressive performance on psychiatric licensing exams and text-based vignettes (Hua et al. 2025; Omar et al. 2024), but they are inherently blind to audiovisual signal appearance, motor behavior, acoustic speech characteristics, and affective displays that are the lifeblood of the real-world MSE. MFMs overcome this sensory deprivation by processing interleaved clinical inputs in a single forward pass, generating both explicit reasoning traces and domain-level determinations without requiring task-specific training (Dai et al. 2025). The recent advent of open-weight MFMs, whose pre-trained parameters are publicly available for independent, local execution rather than locked behind proprietary APIs, is particularly transformative for clinical research. This level of access guarantees the full reproducibility, independent auditing, and community-driven iteration required for medical science (Gao et al. 2025; Passos et al. 2016). However, the theoretical capacity to process multimodal data does not yet transfer into clinical utility. Whether these architectures can achieve meaningful, domain-specific agreement with practicing psychiatrists remains an open empirical question.

Current evaluations of model-assisted psychiatric assessment are constrained by three distinct methodological limitations. First, most rely on a single, fixed reference standard, artificially erasing the clinically informative variance that naturally occurs across institutions (Di Forti et al. 2025). Second, the reliance on aggregate performance metrics such as overall accuracy or F1 scores creates an “aggregate illusion”, masking domain-level heterogeneity and potentially obscuring significant model failures in clinically vital MSE categories (Blaabjerg et al. 2020). Finally, the standard application of Cohen’s *κ* frequently yields highly unstable estimates in psychiatric datasets characterized by low pathology prevalence, necessitating robust, prevalence-adjusted metrics like Gwet’s AC1 (Feinstein and Cicchetti 1990; Wongpakaran et al. 2013). No existing framework integrates multi-center expert benchmarking, MFM evaluation, domain-level error mapping, and prevalence-adjusted statistics all at once.

To bridge these technological and methodological divides, we introduce a multi-center, domain-level benchmarking framework to evaluate the clinical reasoning capabilities of open-weight MFMs demonstrated here using Qwen3-Omni (Xu et al. 2025). Rather than relying on static text or single-rater ground truths, our approach leverages longitudinal clinical simulations (Martin et al. 2020) to precisely isolate model errors across varying stages of diagnostic complexity and symptom severity. By simultaneously evaluating model performance against the natural distribution of multi-center expert consensus, we identify the specific cognitive thresholds where algorithmic inference breaks down. Ultimately, by mapping where AI aligns with and diverges from expert judgment, this work establishes human clinical variance not as an obstacle, but as a definitive, measurable calibration target for the next generation of psychiatric decision-support tools.

To bridge these technological and methodological divides, we introduce a multi-center, domain-level benchmarking framework and apply it to evaluate the open-weight MFM Qwen3-Omni (Xu et al. 2025). We utilized a peer-reviewed dataset of standardized patient videos portraying three distinct psychiatric diagnoses (schizophrenia, obsessive-compulsive disorder, and bipolar disorder) across three longitudinal timepoints of escalating symptom severity (Martin et al. 2020). These controlled simulations provided a known diagnostic trajectory, enabling the precise isolation of model errors to specific MSE domains and clinical stages. By benchmarking the model against independent expert panels from UTHealth and Yale, we simultaneously estimated baseline inter-expert agreement, model-to-expert agreement, and domain-specific error structures. By empirically mapping where AI aligns with and diverges from expert consensus across the full diagnostic spectrum, this work establishes human clinical variance not as an obstacle, but as a definitive, measurable calibration target for the next generation of psychiatric decision-support tools.

## Methods

### Study Design

This study employed a multi-center benchmarking design to evaluate the agreement between the open-weight multimodal foundation model (MFM) Qwen3-Omni (Xu et al. 2025) and independent expert clinical panels at two academic medical centers (UTHealth and Yale). The evaluation framework was structured around three pairwise comparisons: MFM versus UTHealth, MFM versus Yale, and the inter-expert contrast UTHealth versus Yale. The inter-expert comparison served as the definitive baseline against which model-to-expert agreement was calibrated. All analyses were conducted on a shared benchmark dataset of simulated patient videos with known diagnostic profiles and calibrated symptom severity, ensuring that inter-rater disagreements could be strictly attributed to variance in clinical interpretation rather than ambiguity in the underlying clinical material.

### Benchmark Dataset

The benchmark dataset was derived from a peer-reviewed collection of simulated patient (SP) video recordings originally developed for MSE instruction (Martin et al. 2020). The dataset comprises nine video segments (each under 3 minutes) depicting three simulated adult patients, portraying schizophrenia, obsessive-compulsive disorder (OCD), and bipolar disorder. For each diagnosis, segments were recorded at three successive longitudinal timepoints (T0, T1, T2) of escalating symptom severity. The SP cases were scripted using patient composites from clinical experience and performed by professional actors.

Video segments followed a scoring rubric anchored in the ABC-STAMPS framework (Martin et al. 2020). Raters classified the presence or absence of individual MSE items across 10 domains: Affect and Mood (5 items), Appearance, Behavior and Cooperation (8 items), Perceptions (4 items), Speech (4 items), Suicidality (2 items), Thought Content A: Delusions (5 items), Thought Content B: Obsessions (4 items), Thought Content C: Compulsions (4 items), Thought Process A: Coherence (4 items), and Thought Process B: Speed (4 items). Each video yielded 44 binary item-level classifications, totaling 396 observations across the dataset. The overall expert-annotated pathology prevalence was 16.9% (UTHealth) and 17.2% (Yale), reflecting the class imbalance typical of structured psychiatric classification. (Original case scripts, scoring sheets, and answer keys are reproduced in the Supplementary Materials).

### Expert Annotation

The dataset was independently annotated by expert clinical panels at two academic medical centers. At Yale, a multidisciplinary panel (a senior psychiatrist, psychiatric nursing specialist, psychiatry resident, and senior medical student) independently rated each video and utilized iterative consensus resolution to arrive at final scores (pre-consensus inter-rater agreement: 89%, 𝛋 = 0.66) (Martin et al. 2020). At UTHealth, an independent panel (a senior child and adolescent psychiatrist with extensive adult diagnostic experience, a psychiatry fellow, and a psychiatry resident) followed an identical annotation procedure and consensus resolution, blinded to the Yale ratings (pre-consensus inter-rater agreement: 89.9%, 𝛋 = 0.76). Both panels utilized the identical scoring sheet and answer key format. The variation in panel composition addresses whether the benchmarking framework generalizes across rater training levels and disciplinary backgrounds.

### Model and Inference Procedure

We deployed the Qwen3-Omni-30B-A3B-Thinking model (Xu et al. 2025) . This open-weight MFM utilizes a Thinker-Talker Mixture-of-Experts (MoE) architecture (Zhao et al. 2026; Chen et al. 2026). The “Thinker” component processes interleaved text, image, audio, and video inputs to produce high-level multimodal representations, while the “Talker” generates textual or speech outputs. Both employ MoE architectures (Li et al. 2024) to support high concurrency, comprising 30 billion total parameters with approximately 3 billion active parameters per inference step.

The model’s vision encoder - initialized from SigLIP2-So400m (Udandarao et al. 2025) with approximately 543 million parameters processes image and video inputs at a dynamic frame rate aligned with the audio sampling rate. Audio inputs are processed via an Audio Transformer (AuT) encoder (Xu et al. 2025) trained on 20 million hours of supervised data, producing representations at 12.5 Hz. Temporal alignment is maintained through Time-aligned Multimodal Rotary Position Embedding (TM-RoPE), capturing both local and long-range audio-visual dependencies. The “Thinking” variant was specifically selected for its ability to generate explicit intermediate reasoning steps prior to final outputs - a strict requirement for auditable clinical decision-making. Full-precision inference was conducted on a single NVIDIA RTX Blackwell GPU with 96 GB VRAM, using the vLLM backend configured with a maximum context length of 65,536 tokens and a GPU memory utilization of 0.92.

To mechanistically isolate the reliance of clinical inference on higher-order reasoning capacity versus multimodal perceptual encoding, we conducted a model capacity sensitivity analysis using a 4-bit quantized variant of the model. Post-training weight quantization was performed using the llm-compressor (Lin et al. 2025) one-shot pipeline with compressed-tensor serialization. Quantization was applied to linear layers in the thinker/LLM backbone utilizing an INT4, symmetric, group-wise scheme with a group size = 32 and a mean squared error observer. To rigorously preserve multimodal and routing fidelity, we excluded all audio and visual-tower modules, MoE gate network, and the language model (LM) head, maintaining these sensitive components and all activations in 16-bit precision (W4A16). Calibration utilized 256 text samples from mixed-domain corpora (see Supplementary Methods: Model Quantization Mixed Domain Corpora) with a maximum sequence length of 1,024 tokens. This configuration successfully compressed the ∼60 GB BF16 baseline into a ∼19 GB artifact while preserving vLLM compatibility. The quantized run was executed on a CUDA-enabled single-GPU system utilizing accelerate (Gugger et al. 2022) for automatic memory partitioning (allocating ∼80% of available GPU VRAM and host RAM) and temporary CPU offloading.

### Clinical Video Analysis Algorithm

To circumvent the fundamental constraint that full audiovisual clinical videos exceed the model’s single-pass processing capacity, we developed a three-phase clinical video analysis algorithm **(Fig. 1)**. Phase 1, Segments-level observation with rolling memory., Videos were divided into 30-second temporal segments with a 10-second (33%) overlapping sliding window to ensure temporal continuity **(Fig. 1A)**. Both audio and video modalities were enabled. Guided by a structured YAML prompt (see supplementary materials), the model analyzed each segment for behavioral and acoustic evidence relevant to specific MSE items. To maintain continuity, a rolling context mechanism injected a text summary of the two preceding segments into the current prompt. Once three prior segments accumulated, the algorithm compressed earlier observations into a condensed summary (max 250 words) while retaining the two most recent segment outputs verbatim. Notably, this hierarchical memory kept inputs within the MFM processing limits while preserving both the longitudinal trajectory and granular clinical detail. Phase 1 produced structured JavaScript Object Notation (JSON) outputs containing binary determinations, evidence summaries, modality congruence, and evidence strength (see supplementary materials). Phase 2: Cross-segment meta-analysis **(Fig. 1A)**. Following segment processing, accumulated text observations were compiled chronologically and presented to the model in a text-only meta-analysis pass (with no video re-input). This constrained Graphics Processing Unit (GPU) Video Random Access Memory (VRAM) usage while enabling the model to synthesize progression or stability across the full temporal arc. A separate decision-trace prompt generated an auditable clinical reasoning chain, explicitly recording modality contributions and conflict resolutions to output a final binary determination. Phase 3: Longitudinal context transfer **(Fig. 1B)**. For cross-timepoint analysis (T0, T1, T2), the algorithm implemented a cross-visit memory mechanism. Following each timepoint’s meta-analysis, the model generated a 2-3 sentence clinical carry-forward summary. During the subsequent visit (e.g., T1), the prior visit’s summary (T0) was injected into every Phase 1 segment prompt. This ensured subsequent assessments were informed by prior observations, enabling the model to track symptom trajectories and distinguish emergent findings from persistent traits. Longitudinal timepoints were processed strictly sequentially. The complete structured YAML prompts for segment-level analysis, meta-analysis, and decision-trace generation, together with the vLLM inference configuration, are provided in the Supplementary Materials.

**Fig. 1.**
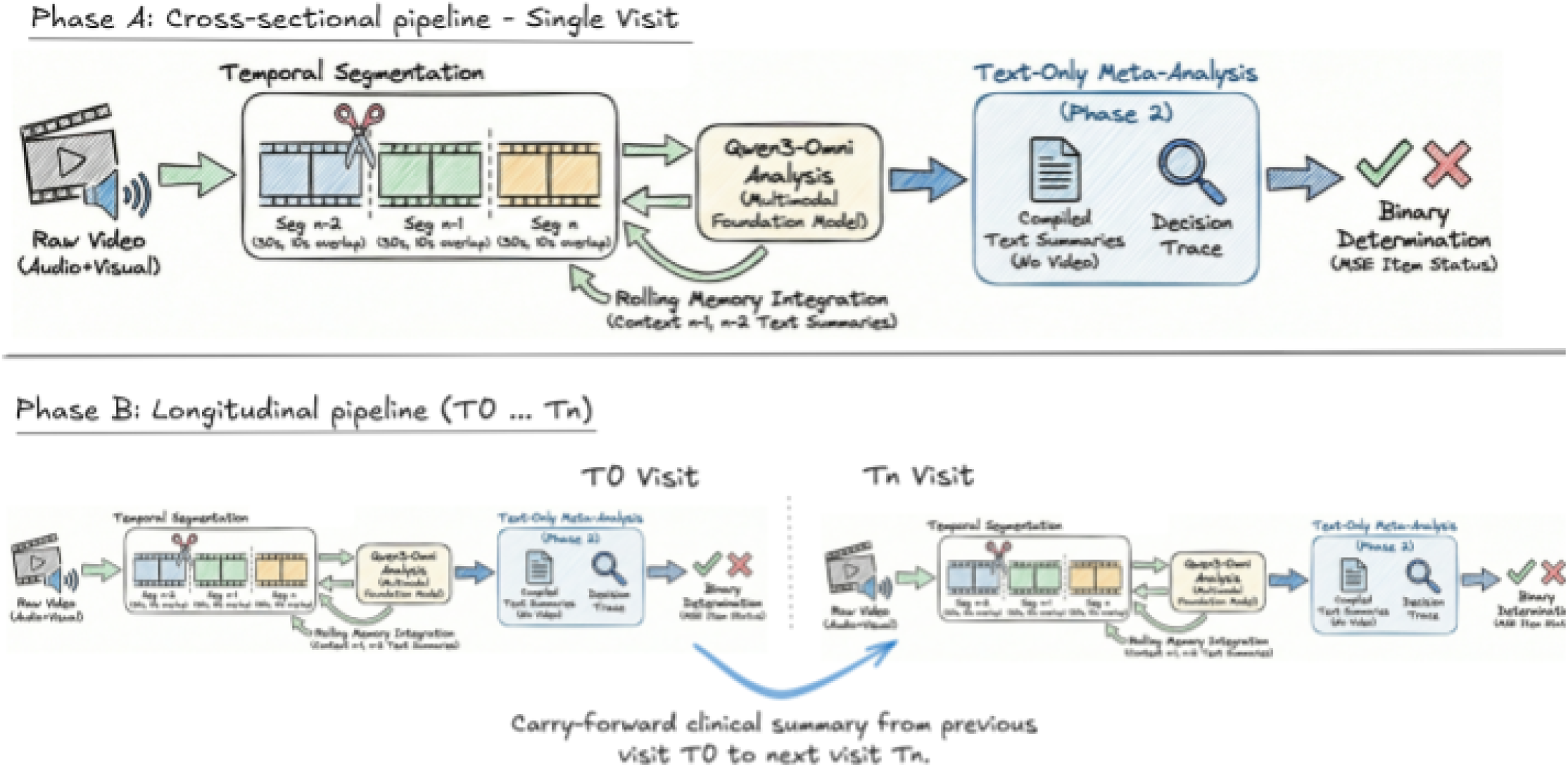
Multimodal clinical video analysis algorithm and evaluation framework. **A**) The cross-sectional inference pipeline. Simulated patient videos are partitioned into overlapping temporal segments and processed by the Qwen3-Omni multimodal foundation model using a rolling memory mechanism. Accumulated segment observations undergo a text-based meta-analysis to generate final Mental Status Examination (MSE) classifications, which arc then evaluated against independent expert consensus panels at UTHealth and Yale. **B**), The longitudinal context transfer mechanism. Across sequential clinical visits of escalating symptom severity (T0, T1. T2). the algorithm generates a structured clinical summary Iron each encounter. This summary is carried forward and injected as prior corniest into the subsequent visit, enabling the model to trade symptom trajectories anil isolating model reasoning errors across distinct psychiatric diagnoses.

### Inference Configuration and Reproducibility

The model operated in instruction-based zero-shot mode; no task-specific training data or few-shot exemplars were provided. Decoding temperature was set to 0.1 to minimize stochastic variation. A fixed random seed (1234) was utilized to ensure reproducibility. Each MSE item was assessed in a single inference pass per video. The reported results strictly reflect the performance of Qwen3-Omni operating within this three-phase temporal segmentation and rolling memory pipeline, which is integral to the system’s output.

### Statistical Analysis

All statistical analyses were conducted on the 396 binary item-level classifications for each of the three pairwise comparisons (MFM vs. UTHealth, MFM vs. Yale, Yale vs. UTHealth). The analytic framework comprised four tiers: 1) Agreement quantification, 2) Directional classification profiling, 3) Systematic bias testing, and 4) Regression modeling of error structures.

### Agreement Metrics

Inter-rater agreement was quantified using Gwet’s AC1 as the primary statistic and Cohen’s 𝛋 as a conventional comparator. Gwet’s AC1 was selected for its mathematical robustness to the prevalence and marginal homogeneity problems, often termed the “kappa paradox” - which produce artificially deflated 𝛋 estimates when pathology prevalence is low (Feinstein and Cicchetti 1990; Wongpakaran et al. 2013; Gwet 2008). Given the dataset’s low overall pathology prevalence (16.9% - 17.2%), this instability was anticipated and is empirically confirmed by the substantial divergence between AC1 and 𝛋 in the model-to-expert comparisons. Agreement benchmarks followed (Landis and Koch 1977) where values below 0.40 were considered poor, 0.40 to 0.60 moderate, 0.60 to 0.80 substantial, and above 0.80 near-perfect.

### Classification Metrics

To characterize the directional error profile of the model, we computed sensitivity, specificity, positive predictive value (PPV), negative predictive value (NPV), F1 score, accuracy, balanced accuracy, and the Matthews correlation coefficient (MCC). MCC, which incorporates all four quadrants of the confusion matrix, was included as a robust measure of classification quality under severe class imbalance (Chicco and Jurman 2020). These directional metrics were strictly applied to the model-to-expert comparisons, as neither expert panel constitutes an absolute ground truth for the inter-expert comparison.

### Confidence Intervals

95% confidence intervals (CIs) for all agreement and classification metrics were estimated using cluster bootstrap resampling (2,000 replicates). To properly account for within-cluster dependence (396 observations nested within 9 videos), video-level resampling (with replacement) was utilized for primary CI estimates. Multiplicity correction was not applied across the three distinct pairwise comparisons, as each tests an independent scientific hypothesis (i.e., establishing a performance ceiling versus testing independent generalizability).

### Stratified Analysis

To isolate spatial and temporal error structures, all metrics were stratified by MSE domain (10 domains), longitudinal timepoint (T0, T1, T2), simulated patient (schizophrenia, OCD, bipolar disorder), and individual video. Domain-level stratification served as the primary analytic lens to identify systematic model divergence from expert consensus. Timepoint-level stratification tested whether model-to-expert agreement degraded as symptom severity increased, and whether inter-expert agreement exhibited comparable degradation.

### Systematic Bias Testing

McNemar’s test (Pembury Smith and Ruxton 2020) was utilized to evaluate systematic directional bias between rater pairs. Net bias was computed as the arithmetic difference between the predicted positive rate and expert-annotated prevalence. A non-significant McNemar’s test indicates that observed positive rate differences are consistent with chance variation rather than systematic over- or under-prediction.

### Regression Modeling of Error Structure

Generalized estimating equations (GEE) with a binomial family and logit link modeled the probability of agreement between rater pairs as a function of MSE domain. The models adjusted for timepoint and subject, utilizing an exchangeable correlation structure to account for within-video clustering (9 clusters). Holm’s step-down procedure was applied to correct for multiplicity across the nine domain contrasts. Because the cluster count (n=9) falls below the conventional threshold for stable GEE robust sandwich estimators (Mancl and DeRouen 2001), small-sample variance constraints inherently limit precision. Consequently, derived p-values are explicitly interpreted as exploratory rather than confirmatory.

### Model Capacity Sensitivity Analysis

To statistically evaluate the contribution of parameter capacity to clinical reasoning, all agreement and classification metrics were recomputed for the 4-bit quantized Qwen3-Omni variant against both Yale and UThealth reference standards. This enabled a direct analytical comparison of error distributions between the full-precision (∼60 GB) and quantized (∼19 GB) models, testing the hypothesis that reduced capacity disproportionately degrades performance in inferential domains relative to observable domains.

### Software

All software code and statistical analyses were implemented in Python 3.11. Confusion matrices and derived classification metrics were computed utilizing scikit-learn (Pedregosa et al. 2011). Gwet’s AC1 was implemented following its original formulation (Gwet 2008), and McNemar’s test was computed through the Scipy python library (Virtanen et al. 2020). GEE models and Holm multiplicity corrections were executed using the statsmodels package (Seabold and Perktold 2010) . Model inference was orchestrated using the vLLM (Kwon et al. 2023) backend via a custom pipeline managing single timepoint execution and longitudinal context transfer.

### Ethics Statement

The standardized patient videos utilized in this study were derived from a peer-reviewed, open-access educational resource published under a Creative Commons Attribution-NonCommercial license (Martin et al. 2020). The original study received institutional review board approval from the Yale Human Investigations Committee (No. 2000024005). Approval for secondary analysis of this data was also obtained from the University of Texas Health Science Center at Houston (UTHealth) Institutional Review Board (No. HSC-MS-25-0671). The videos depict professional actors portraying scripted psychiatric presentations; no real patient data were used. As this study involves the secondary analysis of published educational materials using simulated data, with no personally identifying information collected, it was conducted strictly under the ethical framework established by the original publication.

### Data Availability

The simulated patient video recordings used in this study are available as appendices to the original publication (Martin et al. 2020). The statistical results files (agreement metrics, stratified analyses, and GEE outputs) generated during this study will be deposited in a public repository upon publication.

### Code Availability

The complete analysis prompts and the vLLM setup configuration scripts will be made publicly available in a GitHub repository upon publication.

### Author Contributions

B.M. conceptualized the study, developed the AI software code, performed statistical analyses, and contributed to drafting the manuscript. H.J.A.S.H. conceptualized the study, performed statistical analyses, and contributed to writing the manuscript. C.A.S. conceptualized the study, contributed to the interpretation of statistical data and manuscript writing, and was involved in the MSE scoring. N.D. and P.C. performed the MSE scoring and contributed to manuscript writing and critical review. MJ was involved in study conceptualization, data management and manuscript drafting. AM was primary author on the source clinical paper, and was involved in study conceptualization and drafting of the manuscript. MS,JCS and GBZ were involved in the study conceptualization and drafting of the manuscript.

## Results

### Inter-Expert Agreement

Expert clinical panels at UTHealth and Yale demonstrated substantial consensus across the 396 item-level classifications (Gwet’s AC1 = 0.87, 95% CI [0.79, 0.94]; Cohen’s 𝛋= 0.67, 95% CI [0.54, 0.79]; overall accuracy = 90.7%). The confusion matrix revealed near-symmetric discordant counts (18 vs. 19 items), and McNemar’s test was non-significant (*p* = 0.82, net bias = -0.003), confirming the absence of systematic directional bias between the two academic centers.

Domain-level inter-expert disagreement, however, was not uniform **(Supplementary Fig. 5)**. Thought Process: Coherence exhibited the highest disagreement rate (19.4%), followed by Appearance, Behavior and Cooperation (13.9%), and Affect and Mood (8.9%). Four domains shared an 8.3% disagreement rate (Perceptions, Speech, Obsessions, Compulsions), while Thought Process: Speed (5.6%) and Delusions (4.4%) yielded lower disagreement. Suicidality achieved perfect inter-expert consensus (0% disagreement). Inter-expert agreement also varied by diagnosis, with highest consensus for bipolar disorder (97%) and lower agreement for OCD (89%) and schizophrenia (86%) **(Supplementary Fig. 4a)**. Both institutions exhibited highly congruent pathology prevalence estimates across diagnoses and domains **(Supplementary Fig. 4b, c)**, confirming the absence of systematic institutional thresholding bias. This domain-specific ranking establishes the inherent difficulty gradient of the MSE, providing a calibrated baseline against which model errors can be evaluated.

### Model-to-Expert Agreement

The model achieved moderate agreement with both expert panels: AC1 = 0.70 (95% CI [0.59, 0.78]) and Cohen’s 𝛋 = 0.29 (95% CI [0.15, 0.41]) against UTHealth; and AC1 = 0.72 (95% CI [0.60, 0.82]) and Cohen’s 𝛋 = 0.35 (95% CI [0.16, 0.52]) against Yale. For the UTHealth comparison, model agreement fell significantly below the inter-expert ceiling (model upper bound 0.78 vs. inter-expert lower bound 0.79). For the Yale comparison, the confidence intervals showed only marginal overlap (0.82 vs. 0.79). Notably, the model achieved nearly identical agreement with both independent panels despite variations in panel composition and institutional setting, suggesting its error profile reflects the intrinsic limitations of its own clinical reasoning rather than idiosyncrasies of a specific expert reference standard.

The model’s aggregate error profile was characterized by high specificity (0.86 vs. UTHealth, 95% CI [0.83, 0.88]; 0.87 vs. Yale, 95% CI [0.84, 0.90]) but severely limited sensitivity (0.45 vs. UTHealth, 95% CI [0.32, 0.60]; 0.50 vs. Yale, 95% CI [0.33, 0.69]) **(Fig. 2a, b; Supplementary Fig. 1a, b)**. At the aggregate level, the model’s predicted positive rate (19.4%) nominally exceeded the expert-annotated pathology prevalence (16.9%-17.2%). Yet, McNemar’s test remained non-significant across both comparisons (p = 0.29 and p = 0.36); **(Supplementary Fig. 1c, d)**, indicating that this overall prediction bias was statistically consistent with chance. However, this aggregate equilibrium masked a profound, structured cross-domain mismatch.

**Fig. 2.**
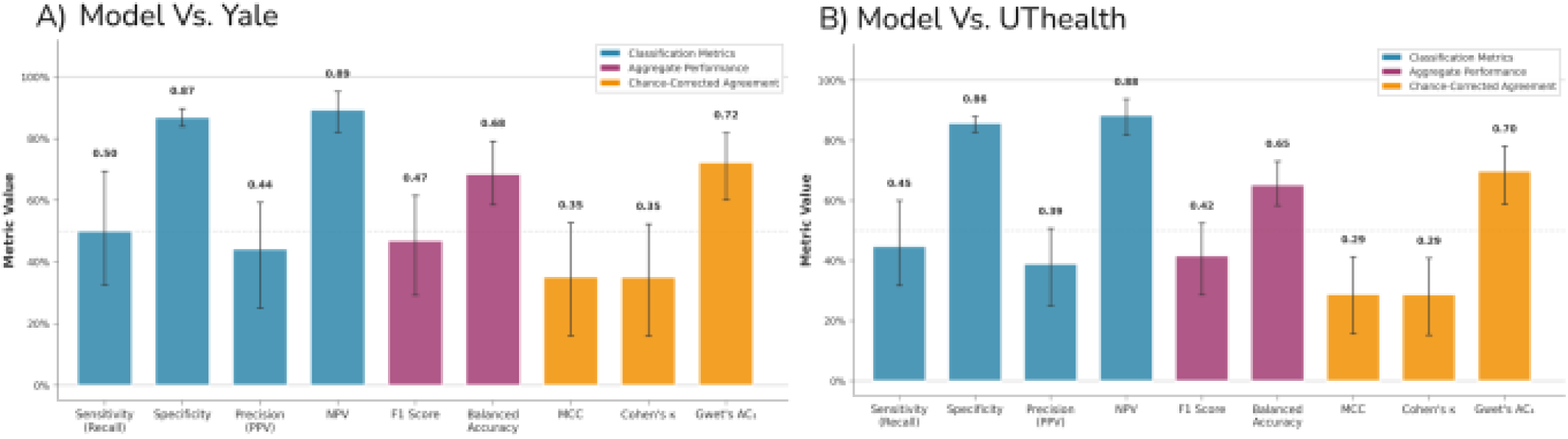
Aggregate classification performance and chance-corrected agreement of the multimodal foundation model. **A**) Performance profile of the model evaluated against the Yale expert consensus. **B**) Performance profile evaluated against the U I Health expert consensus. Across both independent reference standards, the model exhibits a highly consistent aggregate error structure characterized by robust specificity (≥0.86) but severely limited sensitivity (≤0.50). Error bars represent 95% confidence intervals estimated via cluster bootstrap resampling to account far within-video observation dependence.

### Domain-Specific Error Structure

Model errors were structurally organized rather than randomly distributed, exhibiting a highly consistent pattern across both reference standards. Psychiatric classification relies on two distinct evidentiary sources: directly observable behaviors and patient-reported subjective experiences(Clark et al. 2017; Carlat 2023). Following this paradigm, we categorized MSE domains along a continuum from predominantly observable (pathology expressed through visible/audible features, e.g., speech, affect and mood) to predominantly inferential (pathology inferred from verbal reports of subjective experiences, e.g., delusions, perceptions). Transitional domains, such as thought process: coherence, straddle this boundary, as speech disorganization is audible but its clinical significance demands interpretive judgment.

Crucially, in observable domains, the model exhibited high sensitivity paired with low specificity, indicating a systematic over-prediction of pathology **(Fig. 3a, b; Supplementary Fig. 3)**. In affect and mood, sensitivity reached 0.91 against UTHealth and 1.00 against Yale, while specificity collapsed to 0.68 and 0.67, respectively (predicted positive rate: 46.7% vs. expert prevalence: 24.4% and 20.0%). Similarly, in speech, the model achieved sensitivities of 0.78 and 0.88 but specificities of only 0.52 and 0.54 (predicted positive rate: 55.6% vs. expert prevalence: 25.0% and 22.2%).

**Fig. 3.**
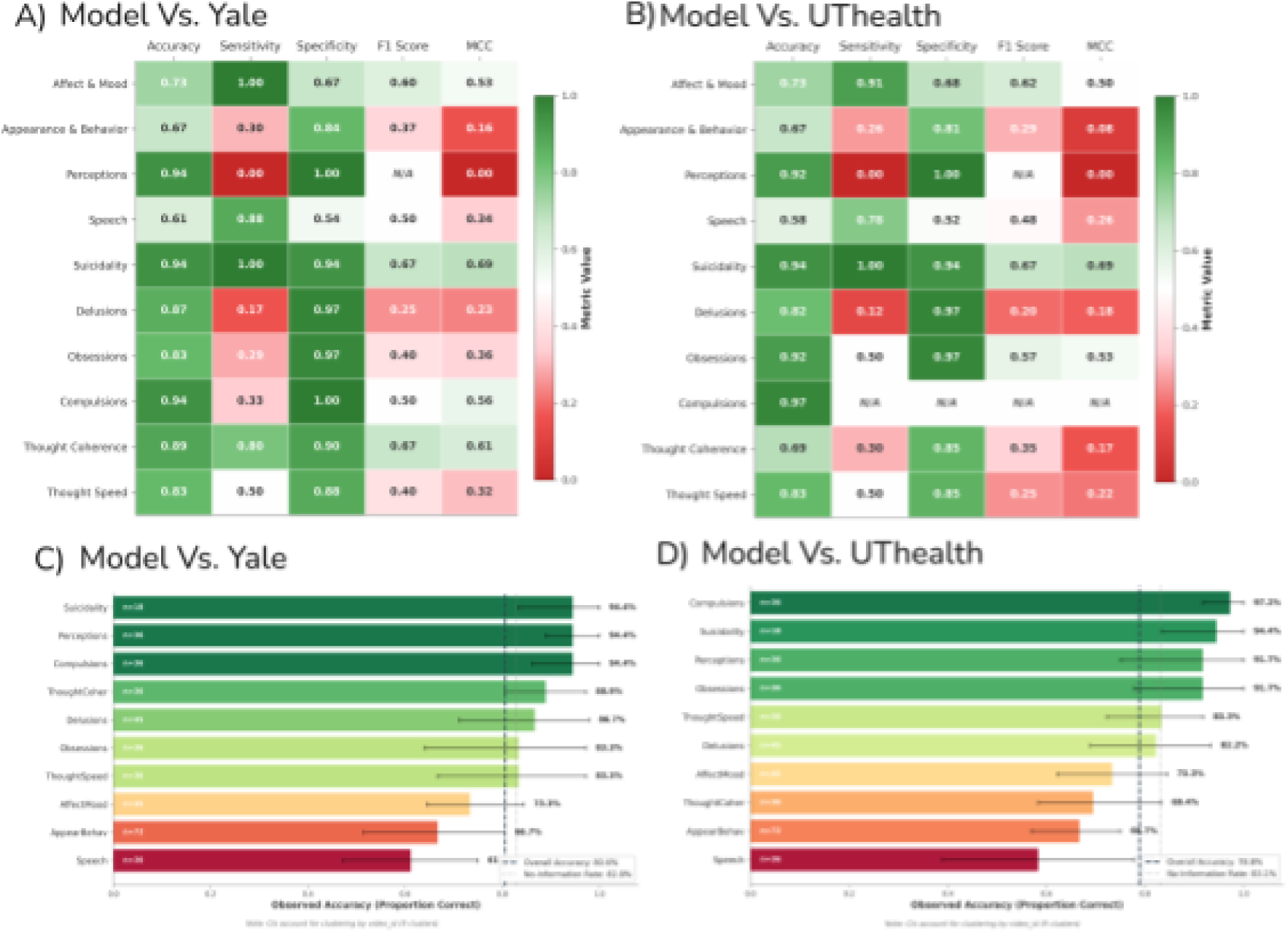
Domain-level error stratification reveals the observable-inferential reasoning gap. **A**) and **B**) Heatmaps detailing multi-metric classification performance across 10 Mental Status Examination (MSE) domains evaluated against the Yale **A**) and UI’Health **B**) expert panels. The model exhibits a distinct diagnostic pattern: systematic over-prediction (high sensitivity, low specificity) in predominantly observable domains (e.g., Affect A Mood, Speech), contrasted with severe under-detection (near-zero sensitivity, high specificity) in inferential domains requiring the interpretation of latent mental content (e.g., Perceptions. Delusions), C and D Domain-specific accuracy distributions against Yale **C**) and UTHealth **D**), Dashed lines denote the aggregate model accuracy and the no-information rale (the baseline accuracy achieved by simply predicting the majority class). Notably, accuracy in several observable domains falls below the no-information baseline due to aggressive over-prediction. Error bars indicate 95% confidence intervals adjusted for within-video clustering.

Conversely, in inferential domains, this pattern inverted: sensitivity approached or reached zero while specificity became near-perfect. The complete failure in the perceptions domain is clinically profound; the model predicted 0.0% positive despite expert-annotated pathology of 8.3% (UTHealth) and 5.6% (Yale), yielding zero sensitivity. The model was unable to infer the presence of hallucinations from the patient’s verbal report, even when explicitly stated in the simulated speech. In Delusions, sensitivity hovered at 0.12 and 0.17 (predicting 4.4% positive against expert prevalences of 17.8% and 13.3%). Obsessions yielded sensitivities of 0.50 and 0.29. Suicidality represented a notable exception to this pattern, achieving 100% sensitivity against both panels. This may reflect the fact that suicidality assessment in this dataset relied on a single, unambiguous verbal cue, making it functionally observable despite its inferential classification.

This cross-domain mismatch mechanistically explains the paradox of low aggregate sensitivity (0.45-0.50) coexisting with net over-prediction (19.4% vs. 16.9%-17.2%): the model aggressively over-predicted observable pathology while markedly under-detecting inferential pathology. Accuracy bar charts confirm this architecture **(Fig. 3c, d)**: speech (61%), appearance and behavior (66.7%), and affect and mood (73.3%) fell below or near the no-information rate, indicating performance inferior to a baseline classifier that simply predicts “absent” for every item. GEE regression indicated that agreement probability varied across domains, though these results should be interpreted as exploratory given the small cluster count (n=9). Following Holm correction, the strongest domain-level effects were observed for compulsions (corrected p = 0.0023) and obsessions (corrected p = 0.0195) against UTHealth, and perceptions (corrected p = 0.0074), thought coherence (corrected p < 0.001), and compulsions (corrected p < 0.001) against Yale **(Supplementary Fig. 2a, b)**. Given the small cluster count (n=9), these p-values should be interpreted with caution. Clinically, an aggregate sensitivity of 0.45-0.50 dictates that the model misses more than half of all expert-identified pathology, rendering it currently insufficient for unsupervised deployment.

### Model Errors Track Expert Ambiguity

To empirically test whether model errors reflect genuine clinical ambiguity, we examined whether error rates covaried with expert consensus strength. Of the 396 item-level classifications, experts agreed on 359 items (90.7%) and disagreed on 37 items (9.3%). When both expert panels agreed, the model’s baseline error rate was 17%. However, when the human experts disagreed, the model’s error rate surged to 41% against the Yale consensus (a 2.3-fold increase) and to 59% against the UTHealth consensus (a 3.4-fold increase) **(Fig. 4a, b)**. This demonstrates that model errors concentrate precisely where clinical phenomena are most ambiguous to human experts, directly validating the study’s methodological premise: expert variance is not noise, but a measurable property of the clinical domain against which AI must be calibrated.

**Fig. 4.**
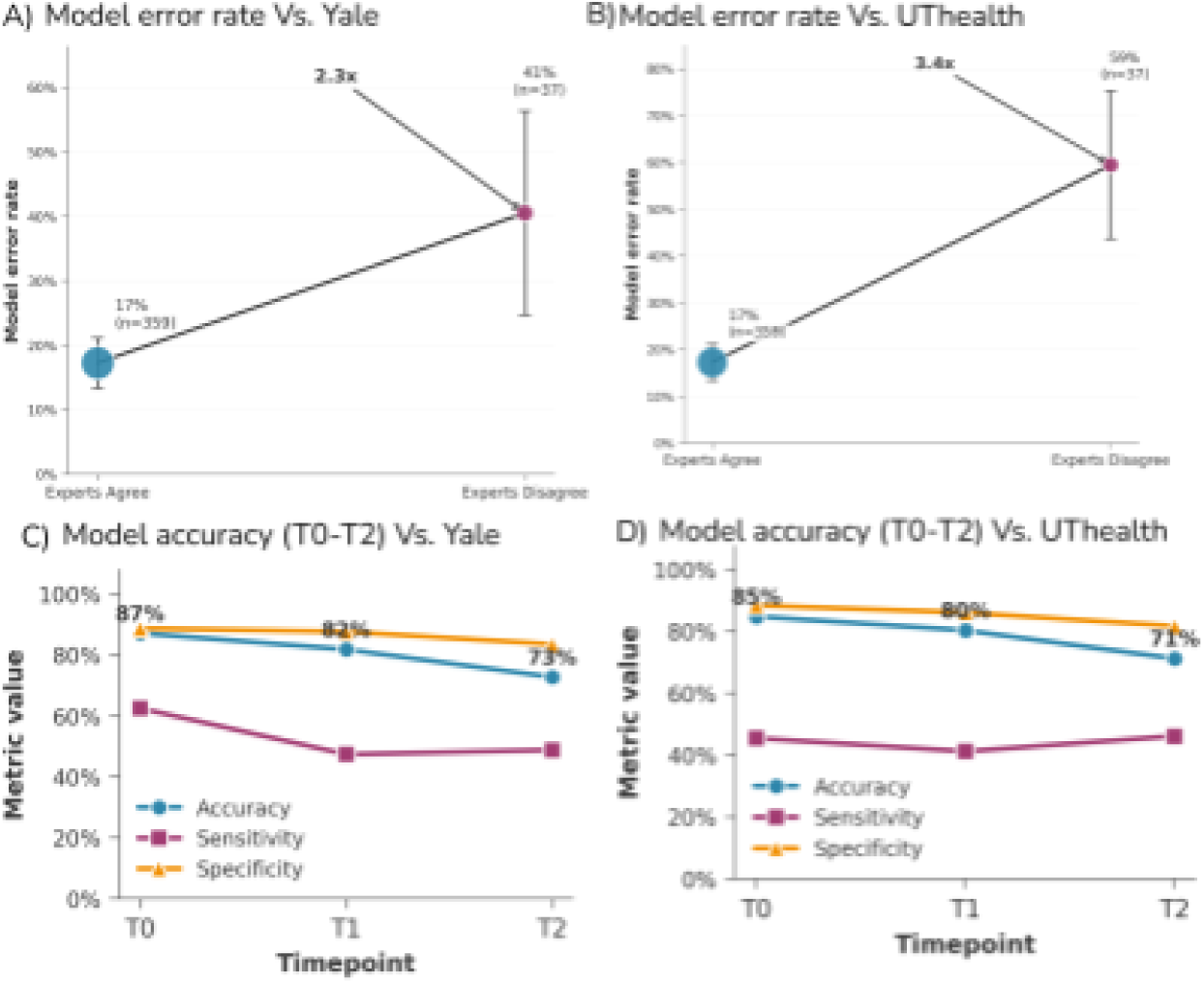
Model errors track export ambiguity and degrade with escalating symptom severity. **A and B**. Model emu rates stratified by inter-expert consensus against the Yale A) and UTHealth **B**) reference standards. The model’s baseline error rate on items with unanimous human agreement (17%) increases substantially (by a factor of 2.3 to 3.4) on items where experts disagree. This demonstrates that algorithmic failures concentrate precisely in areas of inherent clinical ambiguity- **C** and **D**). Longitudinal performance trajectories from baseline (T0) lo maximum Symptom Severity (T2) against Yale (C) and UTHealth (DI. Aggregate accuracy declines monolonically across successive clinical visits. Notably, mode] sensitivity fails to improve despite the higher prevalence of pathology at later timepoints, indicating a specific computational limitation in processing compounding clinical complexity rather than a statistical artifact of changing prevalence. Error bars denote 95% confidence internals.

### Longitudinal Trends in Model-to-Expert Agreement

Model-to-expert agreement declined monotonically as symptom severity escalated across the three longitudinal timepoints. Against UTHealth, accuracy degraded from 84.8% at T0 (pathology prevalence 8.3%), to 80.3% at T1 (prevalence 12.9%), to 71.2% at T2 (prevalence 29.5%). Against Yale, the trajectory was identically structured: 87.1% at T0 (prevalence 6.1%), 81.8% at T1 (prevalence 14.4%), and 72.7% at T2 (prevalence 31.1%) **(Fig. 4c, d)**. This constitutes a stark 13-14 percentage point decline from the mildest to the most severe clinical presentations.

Part of this decline naturally reflects the increased mathematical opportunity for disagreement at higher prevalences, but the model’s sensitivity also failed to improve as more pathology emerged. Against UTHealth, sensitivity remained stagnant (0.45 at T0, 0.41 at T1, 0.46 at T2). Against Yale, sensitivity began higher at T0 (0.63) but deteriorated at T1 (0.47) with only partial recovery at T2 (0.49). The failure to achieve progressively better pathology detection at later timepoints suggests this accuracy decline represents a fundamental limitation in the model’s clinical reasoning under severe conditions, rather than a mere statistical artifact of prevalence.

In contrast, inter-expert agreement proved highly resilient. Yale-UTHealth accuracy was 96.2% at T0, declining only to 87.9% at T1 before stabilizing at 87.9% at T2, despite a further doubling of pathology prevalence. This differential trajectory proves that the model’s performance ceiling is not solely dictated by general task difficulty, but rather reflects a specific inability to navigate increasing clinical complexity-particularly within the inferential domains that dominate severe presentations.

### Diagnosis-Specific Patterns

Model agreement inherently varied across the three simulated patients, each depicting a distinct psychiatric diagnosis. Highest accuracy was achieved for “Robbin” (bipolar disorder; 84.1% vs. UTHealth, 87.1% vs. Yale), followed by “Karthik” (schizophrenia; 78.0%, 78.8%), and “Ben” (OCD; 74.2%, 75.8%). Sensitivity was notably superior for Robbin (0.78, 0.82) compared to Karthik (0.41, 0.43) or Ben (0.20, 0.22). This variation reinforces the observable-inferential hypothesis: bipolar disorder presents prominent observable features highly accessible to audiovisual processing, whereas OCD pathology is predominantly confined to latent thought content that must be inferred verbally. Although this variation may partially reflect case-specific scripting choices inherent to the dataset, the absolute lowest accuracy occurred during the maximum severity T2 timepoints for Ben (OCD; 61.4% vs. UTHealth, 68.2% vs. Yale) and Karthik (schizophrenia; 70.5% vs. UTHealth, 63.6% vs. Yale), cementing that the most challenging barriers for current MFMs are high-severity presentations characterized by inferential pathology (Fig. 5a-c).

**Fig. 5.**
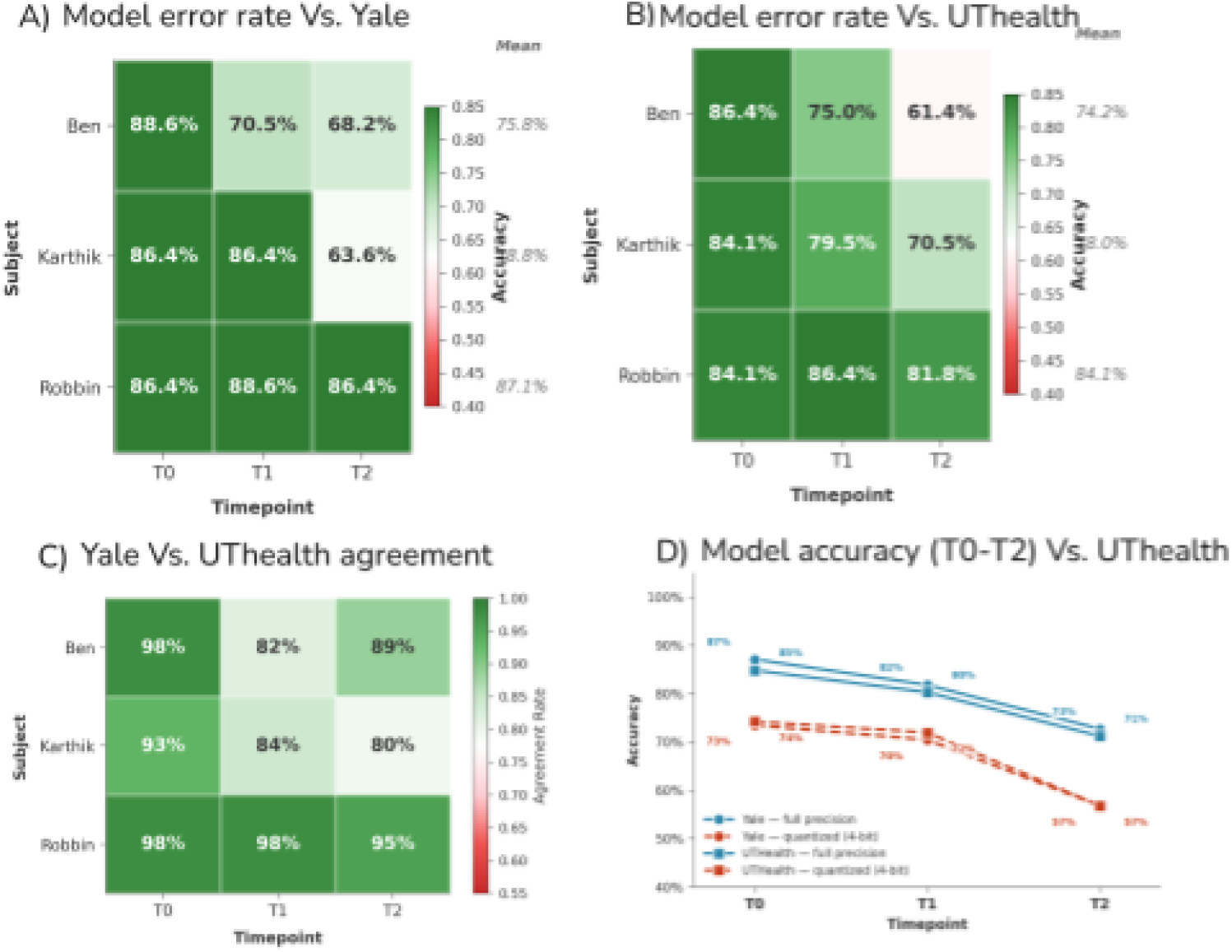
Diagnosis-specific accuracy and the mechanistic impact of model capacity reduction. A and B, Heatmaps displaying model accuracy stratified by simulated patient (diagnosis) and longitudinal timepoint against the Yale A) and UTHealth B) panels. The model achieved highest sustained accuracy for Robbin (Bipolar Disorder), whose pathology features prominently observable motor and affective signs. Conversely, performance degraded most severely for Ben (Obsessive-Compulsive Disorder) and Karthik (Schizophrenia) at the maximum severity timepoint (T2), where pathology relies heavily on latent thought content. C) Inler-expert agreement (Yale vs. UTHealth) stratified by patient and timepoint, establishing a highly robust consensus baseline even at maximum symptom severity. D) Model capacity sensitivity analysis. Evaluating the 4-bil quantized architecture against the fill I-precision baseline reveals a catastrophic decline in aggregate accuracy’ across all timepoints for both reference standards. This capacity-induced degradation disproportionately crippled inferential reasoning capabilities, confirming; that full-precision performance on complex clinical inference relies heavily on parameter capacity rather than basic multimodal perceptual encoding.

### Error-Case Reasoning Trace Audit

To investigate the mechanistic drivers of model failure, a senior psychiatrist conducted a targeted qualitative audit of the model’s reasoning traces, focusing on false-negative cases across clinically high-stakes domains. In inferential domains requiring the interpretation of latent mental content (e.g., perceptions, delusions), errors were almost exclusively driven by interpretive divergence. For instance, the model successfully transcribed simulated patients’ explicit verbal reports of hallucinatory experiences, yet its reasoning engine failed to conceptually map this linguistic content onto the clinical taxonomy of a perceptual abnormality. The traces were structurally coherent, but they exposed a fundamental cognitive bottleneck: the model can process clinical syntax but struggles to execute the higher-order deductions required to synthesize raw narratives into complex psychiatric classifications.

This pattern of interpretive divergence notably extended into predominantly observable domains, revealing a profound miscalibration of clinical thresholds. When analyzing complex behavioral states, psychomotor agitation, or subtle diagnostic phenomena like motor tics, the reasoning traces demonstrated that the model frequently attended to the correct physical movements and anatomical regions. However, it systematically mischaracterized their clinical significance, repeatedly dismissing definitive signs of motor pathology as being “within normal limits” or paradoxically mislabeling pressured speech as “low prosody”. These findings confirm that the model’s failures are not driven by simple perceptual blindness, but rather by an immature diagnostic thresholding mechanism. The model successfully captures the raw phenomenological data but lacks the nuanced, expert-level clinical judgment necessary to differentiate benign behavioral variations from true psychiatric acuity.

### Model Capacity Sensitivity Analysis

The quantized model exhibited severe degradation relative to the full-precision architecture across both reference standards. Evaluated in detail against the Yale panel (n = 396), overall accuracy collapsed from 80.6% to 66.8% (a 13.7 percentage point drop). Gwet’s AC1 plummeted from 0.72 (95% CI [0.60, 0.82]) to 0.44 (95% CI [0.27, 0.59]), shifting the model from substantial to merely moderate agreement. Cohen’s 𝜅 similarly degraded from 0.35 to 0.24. This declining trajectory was identically mirrored against the UTHealth reference standard, where quantized accuracy plummeted across all longitudinal timepoints, ultimately collapsing to 57% at maximum symptom severity.

Mechanistically, the quantized model’s error profile inverted relative to the baseline. Sensitivity artificially inflated from 0.50 to 0.71, while specificity collapsed from 0.87 to 0.66,indicating that the compressed model exhibited a reduced specificity in pathology prediction. The predicted positive rate surged from 19.4% to 40.3%, vastly overshooting the expert-annotated prevalence of 17.2% and McNemar’s test became highly significant (*p* < 0.0001), confirming marked systematic over-prediction (compared to the full-precision model’s calibrated predictions at *p* = 0.36).

Crucially, this capacity-induced degradation was completely asymmetrical across MSE domains. Inferential domains suffered a large mean accuracy decline of 19.2 percentage points (thought process: coherence dropped from 88.9% to 57.1%; suicidality from 94.4% to 61.1%; delusions from 86.7% to 64.4%). In marked contrast, observable domains exhibited a much narrower mean decline of 10.5 percentage points (speech from 61.1% to 50.0%; appearance and behavior from 66.7% to 48.6%; affect and mood from 73.3% to 71.1%). This differential degradation is consistent with the hypothesis that full-precision performance on inferential domains depends more heavily on model parameter capacity than performance on observable domains. When this cognitive capacity is constrained via quantization, domains requiring clinical inference are disproportionately crippled, while those reliant on basic audiovisual feature extraction remain relatively intact.

## Discussion

This study introduces a multi-center benchmarking framework that simultaneously estimates inter-expert agreement, model-to-expert agreement, and domain-specific error structures using prevalence-adjusted statistics. Applied to the open-weight multimodal foundation model (MFM) Qwen3-Omni, we drive three principal findings. First, the model achieves moderate agreement with expert panels at two independent academic centers (AC1 = 0.70-0.72), falling significantly below the inter-expert ceiling (AC1 = 0.87) but demonstrating robust consistency across both reference standards. Second, model errors are not randomly distributed; they are structurally organized along a clinically interpretable continuum, succeeding in observable domains but failing significantly in inferential domains. Finally, and most notably, model errors track expert ambiguity itself. When human panels agree, the model errs on 17% of items. When experts themselves disagree, the model’s error rate surges to 41-59% (a 2.3 to 3.4 fold increase). These findings reframe expert variance from statistical noise into a measurable calibration target, establishing that aggregate performance metrics obscure clinically critical, domain-level failures.

### The Observable-Inferential Reasoning Gap

The dichotomy between observable and inferential MSE domains provides a unifying mechanistic explanation for the model’s error profile, its longitudinal degradation, and the differential impact of model capacity reduction. In observable domains such as speech and affect, the model demonstrates high sensitivity but low specificity. This pattern is consistent with highly competent perceptual feature extraction coupled with an overly liberal diagnostic threshold. In inferential domains such as delusions, hallucinations, and obsessions sensitivity approaches zero.

The complete failure in the perceptions domain is particularly instructive. The model successfully transcribed the simulated patient’s verbal reports of hallucinatory experiences, yet it failed to classify this linguistic content as a perceptual abnormality. This isolates the computational deficit: the model can process speech at the syntactic level but cannot yet execute the higher-order inferential reasoning step required to map a patient’s subjective narrative to a structured clinical taxonomy. This mapping is a complex cognitive operation that clinicians hone through years of supervised psychiatric training (Carlat 2023; Nordgaard et al. 2013). Notably, converging lines of evidence solidify this interpretation. The 4-bit quantization sensitivity analysis demonstrated that when the model’s reasoning capacity was artificially constrained, inferential domains suffered nearly twice the accuracy degradation (19.2 percentage points) compared to observable domains (10.5 percentage points). These findings are consistent with the interpretation that inferential task performance depends more heavily on the reasoning capacity preserved by full-precision weights, though alternative explanations, such as differential sensitivity to quantization noise in low-prevalence domains, cannot be excluded. The longitudinal degradation pattern further aligns with this axis. As symptom severity escalated from T0 to T2, the proportion of complex inferential pathology grew, causing model-to-expert agreement to decline monotonically by 13-14 percentage points while inter-expert consensus remained robustly stable. This differential trajectory indicates that the model’s performance ceiling is not merely a function of general task difficulty, but a specific architectural limitation in navigating compounding clinical complexity.

These findings expose a critical blind spot in prior multimodal AI research, which has largely focused on predicting continuous clinical scores for self-report instruments like the PHQ-9 or GAD-7 (Grimm et al. 2025; Jiang et al. 2024; Chen et al. 2025). Such tasks are core extraction problems based on perception as they do not require the model to reason about latent mental content. Similarly, text-only large language models have demonstrated encouraging results on psychiatric licensing examinations (Hua et al. 2025; Omar et al. 2024), but these benchmarks present clinical information in a pre-processed textual format, artificially removing the burden of real-time audiovisual processing. The MSE imposes both demands concurrently. Our results suggest that while current MFMs can meet the perceptual demand, they have only partially bridged the inferential reasoning gap. This finding echoes emerging evidence across the medical AI literature demonstrating that while foundation models excel at statistical pattern recognition often achieving expert-level performance on structured medical licensing examinations, they struggle acutely with the contextual causal and multi-step logical deduction required for true clinical judgment (Savage et al. 2024; Goh et al. 2024; Thirunavukarasu et al. 2023). Noticeably, the clinical significance of these domain-specific errors varies considerably. False negatives in inferential domains such as suicidality, delusions, and hallucinations carry immediate safety implications that far exceed the consequences of false positives in observable domains such as speech. Any clinical deployment pathway must therefore weight error types according to their downstream clinical consequences, not merely aggregate accuracy.

### Expert Variance as a Reusable Benchmark Property

A central methodological contribution of this study is formally decoupling true algorithmic error from genuine clinical ambiguity. Our empirical data demonstrates this separation: the 2.3 to 3.4 fold surge in model errors on items lacking expert consensus proves that the model failures concentrate precisely where psychiatric phenomena are most ambiguous to human experts. This observation establishes expert variance not as statistical noise, but as a structured, measurable calibration target that dictates the inherent difficulty ceiling of specific clinical presentations. This empirical reality necessitates a fundamental shift in how reference standards are constructed for psychiatric AI decision support tools. Relying on a single expert panel treated as an infallible “ground truth” - a pervasive standard in current literature (Di Forti et al. 2025) is methodologically flawed. Such designs force a binary evaluation that inevitably conflates true algorithmic failure with legitimate clinical disagreement. By measuring model outputs against a single point-estimate rather than the natural distribution of valid clinical opinions, single-panel evaluations artificially deflate performance estimates in domains where human consensus is inherently limited. Furthermore, the empirical divergence between Gwet’s AC1 and Cohen’s 𝛋 in the model comparisons (gaps of 0.37-0.41) provides a concrete, real-world demonstration of the “kappa paradox” in clinical AI evaluation (Aykut et al. 2026; Wang et al. 2026). This underscores that reliance on Cohen’s 𝛋 alone can mathematically mischaracterize model performance by up to 0.41 points under the severe class imbalances ubiquitous in psychiatric classification (Feinstein and Cicchetti 1990; Wongpakaran et al. 2013). By demonstrating consistent model-to-expert agreement across two panels with distinct compositions (multidisciplinary at Yale versus psychiatrist-only at UTHealth), this framework proves its robustness and transferability as a reusable standard for evaluating future models across institutional divides.

### Limitations

Several limitations warrant consideration. The benchmark dataset utilizes simulated patient videos performed by professional actors. Even as this guarantees the controlled conditions and known diagnostic trajectories necessary for precise error isolation, actor-driven portrayals may manifest more stereotypical than naturalistic clinical presentations (Subodh 2012). Crucially, real-world psychiatric patients frequently lack insight or actively conceal inferential pathology (e.g., guarding delusions), suggesting that the large algorithmic under-detection observed in these explicit simulations likely represents a conservative estimate of the model’s true clinical reasoning gap. Furthermore, the model evaluated these encounters in a strict informational vacuum, devoid of the referral context, psychiatric history, or medication status that routinely anchors human clinical judgment, which may artificially depress model performance relative to real-world practice. Additionally, while our pipeline incorporates longitudinal memory across visits, it may not fully capture the dynamic evolution of mental status within a single encounter (e.g., a patient shifting from guarded to disorganized), a trajectory experienced clinicians routinely track. Model performance must eventually be validated in real-world encounters featuring heterogeneous camera angles, audio degradation, and diverse patient demographics. Additionally, the current dataset is modest in scale (9 videos, 396 classifications) and restricts evaluation to three specific diagnoses among a narrow adult age range, with each diagnosis represented by a single patient. Because MSE presentation and interpretation can differ profoundly across cultural contexts and developmental stages (e.g., pediatric or geriatric populations), this inherently confounds diagnosis-level and subject-level effects and limits broad generalizability. Because both expert panels utilized iterative consensus resolution, the final ratings represent negotiated positions, potentially overestimating the agreement that would be observed on strictly independent ratings. Finally, each MSE item was assessed in a single inference pass. Although a fixed random seed (1234) and low decoding temperature (0.1) were used to reduce stochastic variation, the resulting outputs are not strictly deterministic, and no test-retest reliability was estimated. Additionally, this study evaluates a single multimodal foundation model without a head-to-head comparator. The absence of a text-only baseline or competing commercial model (e.g., Gemini 2.5 Pro) limits the ability to attribute observed performance patterns specifically to Qwen3-Omni versus the broader class of current-generation MFMs. The framework itself, however, is model-agnostic and designed for reuse across successive model generations. Furthermore, while the temperature was set to 0.1 to minimize stochasticity, foundation models remain inherently prompt-sensitive. Future work will include a text-only baseline to explicitly quantify the incremental value of multimodal processing over transcript-based inference alone, and to ensure the algorithm’s rolling-memory summarization phase does not inadvertently dilute the subtle linguistic cues required for inferential reasoning. Finally, because the simulated patient videos and their associated scoring rubrics (Martin et al. 2020) are widely utilized, publicly available educational resources, we cannot definitively exclude the possibility of data contamination within the foundation model’s vast, web-scraped pre-training corpora. However, the model’s highly specific, domain-structured failure modes strongly suggest it was engaging in *de novo* clinical inference rather than recalling memorized ground truths.

### Translational Implications and Future Directions

The observable-inferential error structure identified here dictates a staged translational pathway for AI-assisted psychiatric assessment. Observable domains, where the model demonstrates high sensitivity, are primed for near-term integration as clinician-facing decision-support tools. Analogous to AI screening systems in radiology, these models could continuously flag potential affective or psychomotor abnormalities for human review. Conversely, inferential domains require fundamental computational advancement before clinical deployment. Targeted supervised fine-tuning (SFT) on expert-annotated datasets heavily enriched with latent thought-content labels represents the primary remediation pathway. To ensure clinical safety, this must be coupled with preference alignment techniques such as reinforcement learning from human feedback (RLHF) or direct preference optimization (DPO) specifically designed to penalize hallucinated observations and calibrate the model’s inferential thresholds against psychiatric consensus. Retrieval augmented generation (RAG) approaches that dynamically supply models with clinical reasoning exemplars at inference time may offer a complementary solution. Ultimately, architectural innovations that formally decouple perceptual encoding from clinical reasoning engines may prove necessary to strengthen inferential capabilities independently.

Future extensions of this work must evaluate naturalistic clinical videos to verify the generalizability of the observable-inferential axis across broader diagnostic categories with varied demographic profiles, such as major depressive disorder and, post-traumatic stress disorder among others. Head-to-head comparisons with commercial closed-source multimodal models will quantify the current competitive landscape, while systematic clinician auditing of decision traces at scale will isolate whether inferential failures stem from reasoning deficits, perceptual limitations, or hallucinated observations. Tracking these metrics longitudinally across successive foundation model generations will provide the field with a definitive, empirical measure of our progress toward truly intelligent, AI-augmented psychiatric care.

## Supporting information

case scripts, prompts and other supplements.

## Acknowledgments

This work was supported by the NECMHR01 grant from the Texas Child Mental Health Care Consortium (TCMHCC).

