## Supplemental materials for "Human vs AI Clinical Assessment: Benchmarking a Multimodal Foundation Model Against Multi-Center Expert Judgment on the Mental Status Examination"

**Supplementary materials**

**Supplementary Fig. 1 Aggregate confusion matrices and directional bias analysis.** **A** and **B**, Item-level confusion matrices evaluating the multimodal foundation model against the Yale **A**) and UTHealth **B**) expert reference standards (N = 396 classifications per panel). The matrices visualize the model’s baseline error profile, characterized by robust specificity (accurately identifying true negatives in 86-87% of cases) but fundamentally constrained sensitivity (identifying true positives in only 45-50% of cases). **C** and **D**, McNemar’s test for systematic directional bias against Yale **C**) and UTHealth **D**). The bar charts contrast the raw counts of false positives (diagnostic over-prediction) against false negatives (missed pathology). Although the model exhibits a nominal tendency to over-predict pathology (43 vs. 34 errors for Yale; 47 vs. 37 errors for UTHealth), McNemar’s test remains non-significant across both independent panels (p = 0.36 and p = 0.29, respectively).

**Supplementary Fig. 2 Generalized estimating equation (GEE) models of domain-specific prediction accuracy.** **A)** and **B)**, GEE logistic regression results evaluating the effect of individual Mental Status Examination (MSE) domains on the probability of correct model prediction against the Yale **A**) and UTHealth **B**) expert reference standards. To account for within-video observation dependence, models utilize an exchangeable correlation structure (n=9 clusters) and are adjusted for longitudinal timepoint and simulated patient (subject). The ‘Appearance & Behavior’ domain - representing the largest observation subset - serves as the statistical reference category. Odds ratios (OR) > 1 indicate a higher likelihood of accurate model prediction relative to the reference domain. Adjusted-values reflect Holm’s step-down procedure to rigorously correct for multiple comparisons across the nine domain contrasts (p < 0.05, ** p < 0.01, *** p < 0.001).

**Supplementary Fig. 3 Proportional distribution of classification outcomes across MSE domains.** **A** and **B**, Stacked bar charts detailing the relative frequencies of true positives (hits), true negatives (correct rejections), false positives (over-predictions), and false negatives (misses) across the 10 Mental Status Examination domains, evaluated against the Yale (**A**) and UTHealth (**B**) expert panels. This visual stratification explicitly maps the model’s bidirectional error structure. In predominantly observable domains (e.g., Affect & Mood, Speech), the error profile is heavily driven by false positives (light red), confirming systematic diagnostic over-prediction. Conversely, in inferential domains requiring the interpretation of latent mental content (e.g., Perceptions, Delusions), errors are entirely dominated by false negatives (dark red) with true positive detections (dark green) approaching zero. This granular, domain-level outcome distribution visually corroborates the “observable-inferential reasoning gap”, demonstrating that the model’s computational failures are mechanistically distinct depending on the cognitive demands of the clinical category.

**Supplementary Fig. 4 Inter-expert agreement and pathology prevalence distributions across clinical diagnoses and domains.** **A)**. Inter-expert agreement (Yale vs. UTHealth) stratified by simulated patient diagnosis (n = 132 classifications per diagnosis). Maximum consensus was achieved for the bipolar disorder presentation (97%), structurally aligning with its predominantly observable clinical features, compared to high but slightly lower consensus for obsessive-compulsive disorder (89%) and schizophrenia (86%), which rely more heavily on inferential thought content. **B)**. Overall pathology prevalence rates annotated by each expert panel, stratified by diagnosis. Both institutions exhibited highly congruent prevalence estimates, confirming the absence of systematic institutional thresholding bias at the subject level. **C)** Radar chart detailing domain-specific pathology prevalence. The highly overlapping geometrical footprints demonstrate that both independent academic centers applied a fundamentally identical diagnostic schema across the 10 Mental Status Examination (MSE) domains. This tight inter-institutional alignment further validates the human consensus as a robust, objective baseline against which multimodal foundation model errors can be rigorously calibrated.

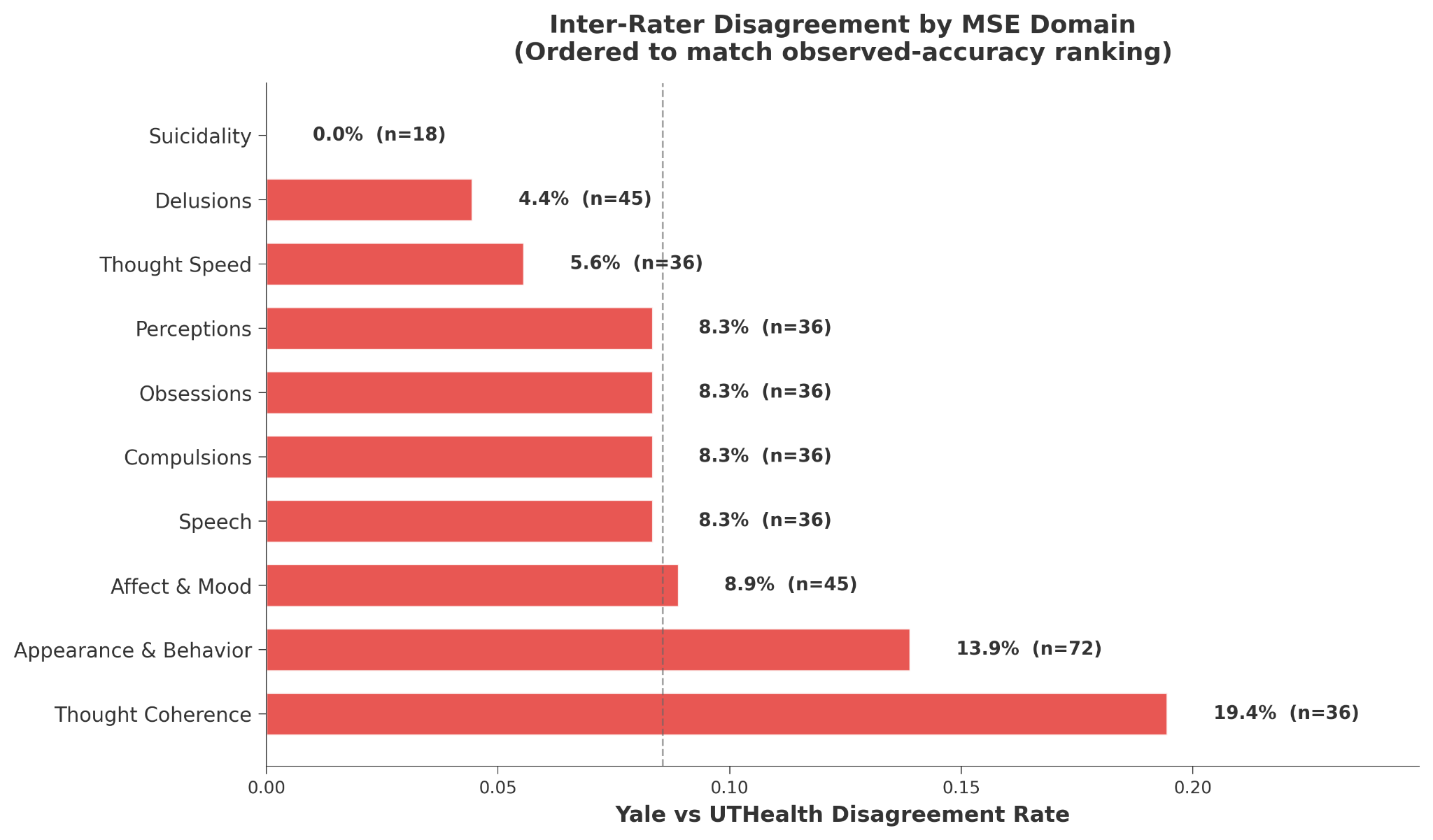

**Supplementary Fig. 5 Inter-expert disagreement rates across Mental Status Examination domains.** Horizontal bar chart detailing the raw disagreement rates between the Yale and UTHealth expert clinical panels across the 10 MSE domains (n = 396 total classifications). Domains are intentionally ordered to mirror the foundation model’s observed-accuracy ranking (from lowest to highest model accuracy). This visual alignment demonstrates that human inter-rater disagreement was most pronounced in domains requiring complex interpretive judgment of disorganized clinical presentations (e.g., Thought Coherence, 19.4%) and broad behavioral synthesis (e.g., Appearance & Behavior, 13.9%), establishing an inherent clinical difficulty gradient. Conversely, highly specific, high-acuity domains achieved near or perfect human consensus (e.g., Suicidality, 0.0%).

**Model quantization mixed domain corpora**

Post-training weight quantization was calibrated on a deliberately mixed text pool combining one broad-coverage English corpus with four mental-health–oriented resources, all loaded from the Hugging Face Datasets hub using the public training splits: WikiText-2 (wikitext, configuration wikitext-2-raw-v1, split train) for general-domain syntax and fluency; Mental Health Condition Classification (sai1908/Mental_Health_Condition_Classification, train); Sentiment Analysis for Mental Health (btwitssayan/sentiment-analysis-for-mental-health, train); Mental Health Counseling Conversations (Amod/mental_health_counseling_conversations, train); and Mental Health Chatbot (heliosbrahma/mental_health_chatbot_dataset, train). For each source we iterated a shuffled index list and attempted to collect up to ⌊256/5⌋ = 51 usable snippets (equal per-source budget before pooling). Rows were converted to single plain-text strings using the field templates in the quantization script (e.g., paired patient/therapist turns or question–answer pairs, depending on the dataset); snippets shorter than 50 characters after stripping were discarded, and accepted text was truncated to 2 000 characters. The combined pool was shuffled again, and calibration used the first min(pool size, 256) examples as a Hugging Face Dataset with a single text column; with the above filters, the documented pipeline yields on the order of approximately 230 retained snippets (hence fewer than 256 if some sources or rows fail quality checks). During calibration, sequences were processed with a maximum length of 1 024 tokens
