## Supplementary material for "Human vs AI Clinical Assessment: Benchmarking a Multimodal Foundation Model Against Multi-Center Expert Judgment on the Mental Status Examination": case scripts, prompts and other supplements.: A. Case Script Ben OCD.docx

**Appendix A**
Ben (Obsessive Compulsive Disorder): Standardized Patient Case

Date: 2/2019

Primary Case Author: Doron Amsalem, MD, Andres S. Martin, MD, MPH

Secondary Case Author: Asaf Jacobs, MS4, Robert Krause, DNP, APRN-BC

Standardized Patient Educator: Robert Krause, DNP, APRN-BC, Andres S. Martin, MD, MPH

Name of Case: Ben - Obsessive Compulsive Disorder (OCD) Scenario

Name of educational and or assessment activity: Mental status exam (MSE) video-based assessment tool

Patient Name: Ben

Chief Complaint: **At his initial visit (T0):** Ben is concerned that he will get ill due to environmental, food, and household contaminants. He also fears that others will have accidents unless things are in the right place (fears traffic signals are not up-to-date). Reports that the degree of occupation with the thoughts during the day is ‘relatively low’ and that ‘it’s all under control’.

**Second visit (T1):** Ben is becoming more preoccupied with the possible health issues caused from the food/beverage contaminants. He is spending a lot of his time researching these contaminants and it is causing him to be late to work. He begins to develop somatic obsessions, worried that he has kidney and liver failure from the chemicals in commercial foods. He does not feel balanced or in control. He hopes the therapist can refer him for a physical workup.

**Third visit (T2):** Ben endorses feeling even less stable and in control than in his previous visit. He has avoided going to work out of fear of saying something inappropriate and has missed deadlines due to compulsive cleaning of his office. Ben is suffering from stomach pain and is now convinced that he has a tumor in his stomach. He asks the therapist to refer him for an MRI or PET scan, convinced that he has a malignant tumor. Moreover, he is afraid that his wife never loved him and believes that her lack of love, combined with the contaminants, have caused this tumor.

Most likely Diagnosis and Differential with rationale from history and/or physical exam:

Ben suffers from obsessive-compulsive disorder (OCD). His symptoms progressively worsen at each visit (i.e.: minimal symptoms at initial visit, severe symptoms at third visit). His thoughts regarding staying safe and healthy turn into obsessions by his second visit and transform into frank somatic delusions by his third visit. More specifically, he displays pathological doubting, somatic obsessions, and environmental obsessions. In addition, he compulsively cleans and obsessively checks for danger. Ben also suffers from physical tics (both neck and eyes) and wrings his hands anxiously. With all the constant worry that Ben displays, general anxiety disorder (GAD) would have to be in the differential as well.

Challenge question: Can learners identify key elements in the mental status exam (MSE), and do their abilities vary according to level of clinical experience and to didactic interventions they are exposed to?

Domains: Check all that apply

- Professionalism
- Communication and Interpersonal skills
- Medical History
- Physical exam
- Shared Decision Making
- Patient Education

X	Clinical Reasoning

- Documentation
- Handoff
- Presentation

X	Other: psychiatric history and clinical exam

Type and level of learner: Medical, nursing and physician associate students; other learners for whom competence on the MSE could be relevant (p. ex. psychiatric technicians, social workers, supporting staff at psychiatric clinics).

Case Objectives: please list specific objectives for each of the domains you have checked above.

Learners viewing the videos should be able to:

1. Identify key components of the MSE, specifically those pertaining to OCD

2. Differentiate obsessional thoughts from compulsive behaviors

3. Distinguish obsessional from delusional thought content, based on the level of associated insight

| SETTING: outpatient, in patient, ED, home, nursing home, rehab, group etc. | Outpatient clinic |
| --- | --- |
| PATIENT PROFILE: Information about the “patient” that helps select an SP and helps the learner get an understanding of them as a person. SP will know more information about the patient than learner will ever ask but allows SP to portray a fully developed patient personality. If none of the items below are particulars for the case please write “all may be used.” | |
| Age range | ~ 40 years old |
| Religious/spiritual background | Not Specified |
| Sex (e.g., male, female, intersex, transwoman, transman) | Male |
| Sexual Orientation (e.g., heterosexual, lesbian, gay, bisexual, pansexual, queer, asexual) | Heterosexual |
| Gender expression (e.g., man, woman, gender queer) | Man |
| Race/ethnicity: | Caucasian |
| Physical description (e.g., BMI, height range) | Height: 6’1-6’3, BMI: 19-24 |
| Physical limitations | None |
| Patient appearance (e.g., disheveled, hospital gown, business casual, casual) | T0 and T1: Business casual (dress pants and button-down)  T2: Casual (Khaki pants and long -sleeve shirt) |
| Moulage + location (e.g., none, bruises, scars, body piercing, tattoos) | None |
| Affect (e.g., pleasant, cooperative) | Euthymic, pleasant and cooperative |
| Family group (e.g., who is family, who they live with) | Lives with his wife Laura and two children (Ben Jr., 14 Sarah; 8) |
| Education | Not specified |
| Level of health literacy | Knowledgeable about his psychiatric condition and adherent to medication and routine visits |
| **Employment, if any - present and past, noting any current stresses** | Works as an engineer, often at different construction sites. He is in charge of making sure that all the building codes are up to date. He worries that the buildings he is working in are unsafe (constantly checking the building codes). He also misses various deadlines at his job due to his inability to get his work done (spending his time compulsively cleaning his office). In addition, he is worried of offending female coworkers by “saying something inappropriate,” which makes him avoidant at the office. |
| Home/homeless - type of dwelling, number of stories, owned or rented | Not specified |
| Financial situation- any current stresses | No current financial stressors |
| Insurance Status (e.g., un/under/insured, public/private, HMO/PPO) | Not specified |
| Habits (i.e., diet, exercise, caffeine, smoking, alcohol, drugs) | Very aware of ‘anything I put in my body’; reads food labels carefully |
| Activities (i.e., hobbies, sports, clubs, friends) | Not specified |
| Typical day - what is the usual daily routine | Conscientious and timely worker, but his increasing preoccupations have more recently made him arrive late on more than on occasion: very unusual for him, and a further source of embarrassment and anxiety. |

| CASE INFORMATION | |
| --- | --- |
| Chief Concern: What the patient will say when greeted by the student. The patient’s primary reason for seeking medical care often stated in his/own words. | T0: “I am feeling good lately. Everything is under control. I am just here for a routine visit.”  (He has a history of OCD, but currently has minimal concerns regarding his level of functioning. He watches what he eats and stays away from certain chemicals, fearing that they could lead to cancer or Alzheimer’s, but he has been this meticulous for many years.)  T1: “I don’t feel so good. I definitely had a relapse. I don’t feel balanced. A lot of my worries have gotten worse. I am having trouble controlling everything.”  (He is worried about the contaminants in his seltzer/food and believes that it is has caused him kidney damage. He also has been second guessing himself at work because he feels the buildings are not safe/up to code.)  T2: “Everything has gotten worse. I don’t feel stable. I don’t feel like I have control. I have not been able to go to work lately. I have a tumor inside my stomach, I need an MRI or PET scan”  (He is now convinced that he has developed a tumor in his stomach from all the contaminants. He also worries that his wife never loved him and just settled for him. He is beginning to avoid going to work altogether). |
| Additional Concerns: Other, if any, concerns the patient has today (i.e., symptoms, requests, expectations, etc.) that will become part of set agenda. | T0: He has concerns regarding environmental, food, and household contaminants but states that his degree of occupation is ‘relatively low.’ He is concerned that the traffic lights on his way to work are ‘screwed up’, so he waits a few extra seconds at the traffic before proceeding. These thoughts/concerns are not interfering with his daily responsibilities and he feels stable.  T1: He is worried about the contaminants in his seltzer/food and believes that it is has caused him kidney damage. He is feeling bloated, nauseous, and complains of burning when he urinates. He asks the therapist to refer him for further work up. He also has been second guessing himself at work because he feels the buildings are not safe/up to code.  T2: He is now convinced that he has developed a tumor in his stomach from all the contaminants. He also worries that his wife never loved him and just settled for him. He is beginning to avoid going to work altogether. His main request is for the therapist to refer him for a PET or ‘whole-body CT’ scan. |
| **THE PATIENT STORY: The SP will be asked to tell their symptom story and the personal and emotion impact for each of their concerns. You will want to write this is the patient voice. The symptom story should be able to answer this question: “Tell me more about [chief concern/additional concern], starting at the beginning and bringing me up to now.”**  **The personal context should be able to answer questions concerning the broader personal/psychosocial context of symptoms, especially the patient beliefs/attributions.**  **The emotional context should be able to ask how are you doing with this, how does this make you feel, how has this affected you emotionally? IMPACT: How has this affected your life? How has this been for your family?** | The conversations between Ben and his doctor take place at an outpatient clinic over three visits/appointments. During his first visit (**baseline, T0)**, Ben presents with **‘minimal symptoms’** and is not too concerned regarding his level of functioning. His second visit takes places three months later **(T1)**, where Ben experiences an exacerbation of his disorder; he presents as anxious and worried (i.e.: **‘medium symptoms’**). The third visit takes place one month later **(T2)**, with Ben displaying **‘severe symptoms’,** including somatic delusions.  First Visit (baseline, T0): Ben arrives at his doctor’s office and begins by sharing some good news regarding a possible promotion at work. He seems a bit nervous about the increase in responsibility, but overall states that the promotion would be a “good thing.” When asked about his current symptoms, Ben states that he feels stable: “My hands are good. I’m not chaffed, I’m not raw. I have it pretty under control.” He does endorse a few symptoms which he describes as “the usual stuff.” These include being careful with what he eats, such as avoiding phthalates, since “they can cause cancer and are linked to Alzheimer’s.” He also states that the traffic lights on his way to work are “totally screwed up” and that “it is an accident waiting to happen.” Therefore, in order to avoid getting into an accident, he waits “an extra 10 seconds once the light turns green” before proceeding. But he describes these symptoms/actions as the “usual stuff, nothing [he] can’t deal with.” In addition, he seems content with his marriage, stating that things with his wife are going well. He mentions that his son just turned 14, which is the age when he first experienced his symptoms. He endorses “watching [his son] more intently,” looking for symptoms, but states that “so far he is a normal 14-year-old boy.” Ben does display **neck and eye tics** throughout the conversation (excessively blinking and tilting his head towards his shoulders), but all his other previous symptoms seem to be under control (i.e.: excessive handwashing etc.). Even his concerns regarding his diet and the traffic lights, while present, have not affected his day to day functioning (i.e.: he still goes to work and is succeeding). Overall, Ben is pleasant and cooperative throughout the interview, displays minimal symptoms, and seems content at work and at home.  Second Visit (T1): Ben arrives for a second visit 3 months later. When asked how he is doing he states “not so good, I definitely had a relapse. I don’t feel balanced. A lot of my worries have gotten worse. I’m having trouble controlling everything.” He is then asked what he is trying to control and states that he recently found out that the seltzer he has been drinking daily is contaminated with chemicals that can “cause kidney failure and possibly cancer.” He seems convinced that he suffered some kidney damage from this contamination, “I just know it has affected my kidney and liver. I have to pee more often and it burns. I need a work up.” Here, Ben is displaying pathological **obsessions** regarding **contamination** (i.e.: chemicals in the seltzer) and **somatic illness** (kidney failure). Although he fears being ill, he is not certain that he is sick; he is still displaying some self-questioning and doubt about his physical symptoms, these are still obsessions and have not yet turned into overt delusions. As the conversation progresses, Ben endorses spending an excessive amount of time online, researching the ill effects of the chemicals he ingested, which was causing him to be late for work. In addition, when he does get to work, he constantly checks the buildings he is going into, believing that they are not up to code. “I am constantly second-guessing going into the buildings. I do not feel safe in them.” Ben’s **compulsive checking** of these buildings, as well as his internet searches regarding the chemicals, are affecting his day-to-day functioning and are no longer just obsessions. Overall, Ben seems agitated and anxious throughout the visit. He displays **akathisia**, uneasily sitting in his chair, and has **neck and eye tics** throughout the conversation. While he is not displaying any delusions or psychotic features, his obsessions have turned into **pathological doubting** and led to **compulsions** in the form of repeated checking, organizing, and ordering and reordering. His symptoms are affecting his ability to function and are causing him a lot of anxiety and distress.  Third Visit (T2): Ben comes for an appointment one month after his last visit. On initial appearance, it is apparent that his **akathisia** is worse than before (he is anxiously moving his right foot up and down and is nervously wringing his hands). In addition, his **grooming and hygiene are inappropriate** (messy hair, wrinkled shirt, etc.) and he is displaying more prominent **neck and eye tics**. He states that he is not feeling good; “Everything has gotten worse. I don’t feel stable and I don’t feel like I have control.” He further explains that he has not been able to go to work lately, after missing deadlines due to his constant **compulsive cleaning** of his desk and computer. Moreover, he is afraid of interacting with female coworkers out of fear that he will say something inappropriate (an **obsession** in the form of **sexual/forbidden thoughts**). Ben’s current symptoms/actions seem to be affecting his marriage as well. He endorses “driving [his wife] crazy” and begins to wonder if she was ever in love with him. “She did not want to kiss me on the first date. She probably never loved me and just settled for me.” As the conversation continues, Ben’s **speech** becomes **pressured** as he complains of having bloating and nausea, due to a tumor in his stomach. He asks the therapist to schedule further workup (PET scan, ‘whole-body CT scan’). The therapist follows this statement by asking if these symptoms may be from the anxiety he is experiencing. Ben denies any possibility that it is from anxiety and is fully convinced that he has a tumor. “This is above and beyond anxiety. It is a tumor and it is probably malignant.” He believes that the tumor was caused by the contaminated beverage and his insecurities with his wife, stating that “they are probably connected.” Ben’s **contamination/environmental obsessions** as well as his **somatic obsessions** have turned into **somatic delusions** (i.e.: he is not merely worried; he is *certain* that he has a tumor). Overall, Ben’s symptoms have become very severe. He can longer function at work and he is beginning to frustrate his wife/family. His obsessions are turning into delusions and he is fully preoccupied with these thoughts. |
| HISTORY OF PRESENT ILLNESS: Although some of the HPI will be given in the patient’s symptom story, the learners will expand the story during the direct question section. Below describe the detailed history, usually about the chief concern, which the student must develop in order to make a useful assessment of the problem: | |
| Onset (when; gradual or sudden) | gradual |
| Setting (what was going on or where was patient when symptoms first noticed?) | T0: He has no ‘new’ concerns or symptoms. His concerns regarding what he puts in his body started years ago. His OCD symptoms began when he was 14 years old.  T1: Ben began compulsively cleaning and obsessively worrying about his health a couple of months after his first visit. It was triggered/exacerbated shortly after finding out that his food/drinks were contaminated.  T2: Four months after his first visit, Ben began worrying that he had a malignant tumor in his stomach. His obsessions and compulsive checking/cleaning caused issues at work and with his wife, which further exacerbated his anxiety/fears that he had cancer. |
| Duration (how long) | His symptoms have been present on and off since the age of 14. |
| Time relationships (frequency, constant or intermittent) | These obsessions occur daily (constant and frequent) |
| Location | N/A |
| Radiation | N/A |
| Quality | N/A |
| Amount | N/A |
| Aggravated by what | External stressors and unpredictability |
| Relieved by what | Following a strict diet (contaminants), double checking everything, feeling safe |
| Associated with what |  |
| Attitude (what does the patient think is the problem, and how does he/she feel about it) | T0: He thinks he suffers from OCD since he was 14 years old. He believes that the best way to manage it is by adhering to his medication and scheduling routine check-ups with his therapist. While he understands that his symptoms have gotten out of control in the past, he currently feels stable and is happy with his progress.  T1: Patient feels unstable and is worried that he has caused damage to his kidneys from consuming contaminants. At the same time, he knows that he has a history of worrying a lot/obsessing over things and understands that there is a possibility that a lot of his symptoms are due to anxiety.  T2: Patient continues to feel out of control but no longer believes that his symptoms are due to anxiety. He is convinced that he has a tumor and no longer connects his fears/obsessions to his OCD. |
| Overall course |  |
| REVIEW OF SYSTEMS: Significant positives and negatives | |
| Past medical history |  |
| Medication allergies (Name and reaction) | NKDA |
| Environmental allergies (Name and reaction) | None |
| Illnesses | None |
| Vaccinations | None |
| Surgeries | None |
| Accidents/ injuries/ trauma | None |
| Hospitalization | Voluntary psychiatric hospitalization at 19yo |
| Inclusive sexual and reproductive history | |
| Sexual practices  Sexual partners  Protection: Use of safer sex practices  Use of birth control if appropriate  Risk of intimate partner violence | Sexually active with his wife. |
| Ob/GYN HISTORY | Age of onset of menses  Age of menopause  Number of pregnancies  Number of live births  Number of miscarriages  Number of abortions |
| Medications | Prescription/dose/reason: serotonin-selective reuptake inhibitor (SSRI) antidepressant: fluoxetine (high dose) + atypical antipsychotic: risperidone (low dose). Both agents were prescribed to help him with his anxiety and OCD.  Over the counter/dose/reason: None  Herbs/supplements/dose/reason: None  Other: |
| Immunizations | - Tetanus - Flu - Hepatitis - Pneumovax - HPV - Other |
| Tobacco products:   - Cigarettes - Cigar - Pipe - Chew - E-cigarettes | - Never - Past- year started/year quit - Current   - Quantity   - # of years |
| Alcohol   - Beer - Wine - Liquor - Other | - Never - Past- year started/year quit - Current   - Quantity   - # of years |
| Drugs   - Weed - Cocaine - Heroin - Meth - Other - IV - Inhalants - Other | - Never - Past- year started/year quit - Current   - Quantity - # of years |
| Diet (describe) | Adheres to a strict diet, avoiding phthalates. |
| Exercise (describe) | Not specified |
| List any other important social history or information important to this case |  |
| Family history |  |
| Mother, Father, Siblings, Grandparents, and other significant findings. | Not specified |
| Physical Exam- List exam maneuvers expected for this case and any abnormal findings that SP will simulate. (tenderness, hyper-hypo reflex, rebound, weakness etc. )  None | |
| PHYSICAL EXAM FINDINGS |  |
| 1) Written in layman’s terms |  |
| 2) General appearance- affect, appearance, position of patient at opening (i.e. sitting, laying down, holding abdomen etc.) | T0: Well-groomed and well dressed, wearing dress pants and a button-down shirt. He is sitting in a chair at the doctor’s office. He is alert and organized. He is cooperative and spontaneous, with good eye contact. Neck and eye tics are noticeable.  T1: Well-groomed and well dressed, wearing dress pants and a button-down shirt. He is sitting in a chair at the doctor’s office. He seems a bit restless (wringing his hands and nervously shaking his legs). Neck and eye tics are noticeable as well. He is cooperative.  T2: Dressed in khaki pants and a Henley long sleeve shirt. Hair is a bit disheveled. He is sitting in a chair at the doctor’s office. Once again, he seems a bit restless (wringing his hands and nervously shaking his legs). Neck and eye tics are noticeable as well. |
| 3) Vital signs | N/A |
| 4) Specific findings and affect | Ben has noticeable tics (forceful blinking and slight twisting of the head and neck). |
| 5) Response to certain physical movements | N/A |
| DIAGNOSIS AND DIFFERENTIAL |  |
| Diagnosis with support from positive and negative history and PE findings | Ben seems to suffer from OCD. His symptoms progressively worsen at each visit (i.e.: minimal symptoms at initial visit, severe symptoms at third visit). His thoughts regarding staying safe and healthy turn into obsessions at his second visit and transform into delusions by his third visit. More specifically, he displays pathological doubting, as well as somatic and environmental obsessions. In addition, he compulsively cleans and obsessively checks for danger. Ben also suffers from physical tics (both neck and eyes) and wrings his hands anxiously. |
| Differential with support from positive and negative history and PE findings | Another possible diagnosis is general anxiety disorder. Ben is constantly anxious throughout the day. He micromanages many aspects of his life and is afraid that things will constantly go wrong. |
| MANAGEMENT OR DIAGNOSITIC PLAN | Ben should continue taking his current medications and should schedule another routine visit with his therapist. |
| PROFESSIONALISM ISSUES OR CHALLENGES: | N/A |
