## Supplementary material for "Human vs AI Clinical Assessment: Benchmarking a Multimodal Foundation Model Against Multi-Center Expert Judgment on the Mental Status Examination": case scripts, prompts and other supplements.: B. Case Script Robbin BD.docx

**Appendix B**
Robbin (Bipolar Disorder): Standardized Patient Case

Date: 2/2019

Primary Case Author: Doron Amsalem, MD, Andrés S. Martin, MD, MPH

Secondary Case Author: Asaf Jacobs, MS4, Robert Krause, DNP, APRN-BC

Standardized Patient Educator: Robert Krause, DNP, APRN-BC, Andrés S. Martin, MD, MPH

Name of Case: Robbin – Bipolar Disorder Scenario

Name of educational and or assessment activity: Mental status exam (MSE) video-based assessment tool

Patient Name: Robbin

Chief Complaint:

T0: Her partner Bobby left her a few weeks ago and she has been feeling tired and sad since. She endorses sleeping up to 16 hours a day and having trouble going to work due to a lack of motivation.

T1 (a few weeks later): Robbin feels that she is doing better and going to work, but feels like she “is just going through the motions.” She is sleeping less but still feels tired all the time. She also feels guilty that she is not doing enough for her daughter Jamie. “I am not doing what I am supposed to be doing as a parent and it is not fair to her.” Robbin is worried that her inability to be happy makes her a bad mother. She also endorses passive suicidal ideation, stating that she would not go through with it because of her daughter, but that she has had “thoughts.”

T2: Robbin has no current complaints, stating that she “feels great.” She is in a rush to get back to work and wants the doctor to hurry up and finish the patient visit.

Most likely Diagnosis and Differential with rationale from history and/or physical exam: Robbins seems to be suffering from a mood disorder, most likely bipolar disorder (BD). In her first visit, she is very depressed, exhibiting psychomotor retardation, a flat affect, hypersomnia, and extreme sadness. In her second visit, her physical energy is better, but she continues to display depressive symptoms (anhedonia, fatigue, guilt, passive SI, etc.). It is possible that during the second visit (T1), Robbin is displaying some mixed symptoms, which is allowing her to have enough physical energy to contemplate what is making her so depressed and suicidal. During her third visit, she exhibits grandiose delusions, pressured speech, mood lability, and inappropriate sexual behavior. She also endorses euphoria and a decreased need for sleep. The switch to mania that is seen during this visit (due to the SSRI) is a good indicator that Robbin suffers from BD.

Challenge question: Can learners identify key elements in the mental status exam (MSE), and do their abilities vary according to level of clinical experience and to didactic interventions they are exposed to?

Domains: Check all that apply

- Professionalism
- Communication and Interpersonal skills
- Medical History
- Physical exam
- Shared Decision Making
- Patient Education

X	Clinical Reasoning

- Documentation
- Handoff
- Presentation

X	Other: psychiatric history and physical exam

Type and level of learner: Medical, nursing and physician associate students; other learners for whom competence on the MSE could be relevant (p. ex. psychiatric technicians, social workers, supporting staff at psychiatric clinics).

Case Objectives: please list specific objectives for each of the domains you have checked above:

1. Identify key components of the MSE, specifically those pertaining to bipolar disorder.

2. Differentiate between depressive, mixed, and manic symptoms, including their degrees of severity.

3. Distinguish between different types of delusions and thought process abnormalities that are often seen during manic episodes.

| SETTING: outpatient, in patient, ED, home, nursing home, rehab, group etc. | Outpatient clinic |
| --- | --- |
| PATIENT PROFILE: Information about the “patient” that helps select an SP and helps the learner get an understanding of them as a person. SP will know more information about the patient than learner will ever ask but allows SP to portray a fully developed patient personality. If none of the items below are particulars for the case please write “all may be used.” | |
| Age range | ~50 years old |
| Religious/spiritual background | Not Specified |
| Sex (e.g., male, female, intersex, transwoman, transman) | Female |
| Sexual Orientation (e.g., heterosexual, lesbian, gay, bisexual, pansexual, queer, asexual) | Lesbian |
| Gender expression (e.g., man, woman, gender queer) | Woman |
| Race/ethnicity: | Caucasian |
| Physical description (e.g., BMI, height range) | 5’4 - 5’6, BMI: 25-30 |
| Physical limitations | None |
| Patient appearance (e.g., disheveled, hospital gown, business casual, casual) | T0: Slightly disheveled and dressed too casually (grey sweatshirt with her hood on, jeans, and a scarf).  T1: Groomed and well-dressed (grey long sleeve Henley and blue jeans).  T2: Abnormal appearance (wearing a lot of makeup, a bright multi-colored button- down shirt, a tie, and colorful plastic hair clips in her hair) |
| Moulage + location (e.g., none, bruises, scars, body piercing, tattoos) | None |
| Affect (e.g., pleasant, cooperative) | T0: Flat, with psychomotor retardation, but pleasant and cooperative  T1: Constricted range of affect and anhedonic, but pleasant and cooperative  T2: Labile with an expansive affect (irritable, euphoric, and sexually inappropriate). |
| Family group (e.g., who is family, who they live with) | Her partner (Bobby) recently left. She lives with her daughter Jamie (age 16). |
| Education | Law School Graduate |
| Level of health literacy | Adherent to her medication and doctor visits. Like many patients suffering from bipolar disorder, her insight into her condition varies. |
| Employment, if any - present and past, noting any current stresses | Works as an attorney and is generally successful, but missed a few days due to fatigue/anhedonia. |
| Home/homeless - type of dwelling, number of stories, owned or rented | Not specified |
| Financial situation- any current stresses | None |
| Insurance Status (e.g., un/under/insured, public/private, HMO/PPO) | Not specified |
| Habits (i.e., diet, exercise, caffeine, smoking, alcohol, drugs) | Not specified |
| Activities (i.e., hobbies, sports, clubs, friends) | Robbin has been too tired and depressed to attend social gatherings or to partake in her hobbies. She is very close with her daughter, but can rely on her too much at times. |
| Typical day - what is the usual daily routine | Robbin works as an attorney at a law firm. After work, she comes home and tries to spend time with her daughter. Lately she feels as though she is “just going through the motions.” She goes to work most of the time, but spends most of her free time in bed sleeping. |

| CASE INFORMATION | |
| --- | --- |
| Chief Concern: What the patient will say when greeted by the student. The patient’s primary reason for seeking medical care often stated in his/own words. | T0: “I’m doing okay, tired.”  (Her partner Bobby left her a few weeks ago and she has been feeling tired and sad since. She endorses sleeping up to 16 hours a day and having trouble going to work due to a lack of motivation).  T1: “I’m doing better. I’m going to work, but just feel like I am going through the motions”  (Robbin has enough energy to go to work, but still “feels tired all the time.” She is also worried about her daughter Jamie. “I am not doing what I am supposed to be doing as a parent and it is not fair to her.”)  T2: “I’m doing great! Let’s go, I gotta get out of here, let’s get out of here!”  (Robbin has no current complaints, stating that she feels great. She is in a rush to get back to work and wants the doctor to hurry up and finish the patient visit.) |
| Additional Concerns: Other, if any, concerns the patient has today (i.e., symptoms, requests, expectations, etc.) that will become part of set agenda. | T0: Robbin feels sad and tired and wants to find a way to feel better. She has trouble going to work, lacking motivation to get out of bed, and asks the doctor for help.  T1: Robbin has enough energy to go to work, but finds that most things are no longer enjoyable. “I am just going through the motions.” “I’m doing what I need to do, but it is just not fun anymore.” She is worried that her inability to be happy makes her a bad mother. Robbin also endorses passive suicidal ideation, stating that she would not go through with it because of her daughter, but that she has had “thoughts.”  T2: Robbin’s only current complaint is that she needs her current visit to wrap up because she needs to get back to work. She has a lot of things to do and cannot be bothered. |
| THE PATIENT STORY: The SP will be asked to tell their symptom story and the personal and emotion impact for each of their concerns. You will want to write this is the patient voice. The symptom story should be able to answer this question: “Tell me more about [chief concern/additional concern], starting at the beginning and bringing me up to now.”  The personal context should be able to answer questions concerning the broader personal/psychosocial context of symptoms, especially the patient beliefs/attributions.  The emotional context should be able to ask how are you doing with this, how does this make you feel, how has this affected you emotionally? IMPACT: How has this affected your life? How has this been for your family? | The conversation between Robbin and her doctor takes place at an outpatient clinic over three visits/appointments. During her first visit **(baseline, T0)**, Robbin presents mainly with **‘depressive symptoms’**; she is anhedonic, sad, and lacks motivation. She is prescribed an SSRI and beings treatment. During the second visit, a few weeks later **(T1)**, Robbin has a bit more energy, but continues feeling anhedonic and sad. She now endorses passive suicidal ideation. Her third visit takes place a few weeks after her second visit **(T2)**, with Robbin experiencing **‘manic symptoms’**, including racing thoughts, lack of need for sleep, and grandiosity.  First Visit (baseline, T0): Robbin arrives at her doctor’s office and is not engaging with her therapist; she is slightly **disheveled**, wearing sweatpants and a hood over her head, and is **lacking eye contact**). The therapist asks her how she is feeling a few times before she finally responds, stating that she is “constantly tired and sad,” sleeping 12 to 16 hours a day. When asked about what is making her sad, she states her partner, Bobby, left a few weeks ago (i.e.: they split up). Throughout the conversation she continues to state that she is tired, displaying **psychomotor retardation, anhedonia,** and a **slow/delayed speech**. She rarely expands on the therapist’s questions, answering with a few words each time. Robbin endorses missing work the last few days, due to a lack of motivation and “feeling tired.” The therapist then asks her if she has “any thoughts of harming herself.” She denies any suicidality, stating that she “just misses her [Bobby].” The therapist asks Robbin about her daughter: “She is at home. She cooks for me and cuddles me in bed.” The conversation abruptly ends here. Throughout the visit, Robbin displays a **constricted/flat affect**, rarely opening up. While it is not stated explicitly in the video, Robbin begins taking an SSRI (Zoloft) after this visit.  T1: Robbin arrives at her doctor’s office a few weeks after their initial visit, and states that “[she’s] better and going to work.” The therapist then asks, “you are going to work, but how do you feel?” Robbin responds, “I am going through the motions. I’m tired. I’m not sleeping as much as I was, but I’m still tired.” While Robbin seems a bit distressed, she is no longer disheveled, she is making good eye contact, and seems to be a bit more conversational (i.e.: no psychomotor retardation). Robbin then expands on what is bothering her: “I am worried about Jamie. I’m not doing what I am supposed to be doing as a parent. It is not fair to her. I’m just not right. I feel off. I miss having fun with my daughter.” Robbin once again displays **anhedonia** and a **flat/constricted affect**. Although she is going to work, she worries that she will be fired. “They know about my condition, but I am not sure how long they will put up with it.” Later in the conversation she brings up her daughter again: “I need to be mom and I don’t feel like mom.” The therapist follows up by asking if “she has had any thoughts of hurting herself.” “No, because of Jamie. But I have thought a few times of not being here because there is nothing here for me.” Robbin is endorsing **passive suicidal ideation**, but has her daughter as a protective factor. She goes on to say “It is not fun. I’m saying that a lot. I just feel flat.” It has been a few weeks since starting the SSRI, and while Robbin has more energy to go work, she also seems to have more energy to think more deeply about what has been bothering her. Unlike her first visit (before the SSRI), she feels guilty about being a bad mother and endorses suicidal ideation. While the SSRI is making her more functional, it is also putting her at greater risk of harming herself (i.e.: she has the physical energy to do so).    T2: Robbin arrives at her doctor’s office a few weeks after their last visit and states emphatically that she “is feeling great!” She is **abnormally dressed** (wearing a lot of makeup, a bright multi-colored button-down shirt, a tie, and colorful plastic hair clips in her hair). She also displays **abnormal eye contact**, staring intensely at the therapist throughout the conversation. After stating that she is doing great, she right away gets agitated. “Yeah, yeah, let’s go! I gotta get out of here. Let’s get out of here.” Throughout the conversation she displays **akathisia/psychomotor activation**, hoping to quickly finish the visit and head back to work. She is very **euphoric/manic** and displays **grandiose delusions**: “I’m a very important person. If you look up the word attorney in the dictionary it shows my face and says the most awesome attorney on the planet.” The therapist then asks her if “she is going to go to work looking like that.” She responds, “of course, I look fantastic. You know how great I look!” Immediately after this response, Robbin gets agitated and shouts “you’re aggravating me now because you asked me this before!” Robbin’s mood is very **labile** throughout the conversation, going from euphoric to agitated. She also displays **pressured speech, loosening of associations, and a tangential/circumstantial** thought process. During the conversation she randomly interjects with off topic comments: “Do you know GE has the best appliances. I use ammonia on the floor, not the store bought stuff. It makes the floors pristine.” Robbin also endorses a **decreased need for sleep** and **distractibility**. “Sleep? Who needs sleep when you are the #1 attorney in the world….oh yeah and the cabinets.” As the conversation continues, Robbin begins to be **sexually inappropriate** towards the therapist and gets too physically close to him **(abnormal physical proximity)**. “You’re very nosy! But you’re really cute too, more cute than nosy.” At this point, she is out of her chair and in his face. The therapist kindly asks her to go back to her seat, stating that “this is not appropriate.” Robbin listens and sits back down, but then continues with her mood lability, distractibility, and sexual inappropriateness: “You’re pissing me off, but you’re awfully cute in those cute littles shoes. What are you doing tonight? Take me out for a drink and I’ll forget all the things you are not doing right Doc!... I need to make steak tonight.” It seems as though the SSRI caused Robbin to switch into mania. The therapist needs to decide what he will do next (i.e.: stop the SSRI, possible hospitalization). |
| HISTORY OF PRESENT ILLNESS: Although some of the HPI will be given in the patient’s symptom story, the learners will expand the story during the direct question section. Below describe the detailed history, usually about the chief concern, which the student must develop in order to make a useful assessment of the problem: | |
| Onset (when; gradual or sudden) | Gradual |
| Setting (what was going on or where was patient when symptoms first noticed?) | T0: Robbin was feeling sad, tired, and depressed for a few weeks after her partner (Bobby) left. She has trouble going to work, lacking motivation to get out of bed, and asks the doctor for help.  T1: Robbin has enough energy to go to work, but finds that most things are no longer enjoyable. “I am just going through the motions.” “I’m doing what I need to do, but it is just not fun anymore.” She is worried that her inability to be happy makes her a bad mother. Robbin also endorses passive suicidal ideation, stating that she would not go through with it because of her daughter, but that she has had “thoughts.” While she was experiencing the depressive symptoms for at least a few weeks before the initial visit, her suicidality and guilt about ‘being a bad mother’ started after her last visit (cannot be longer than a few weeks).  T2: Robbin’s only current complaint is that she needs her current visit to wrap up because she needs to get back to work. She has a lot of things to do and cannot be bothered. From a clinician’s point of view, she has switched into mania (possibly due to the SSRIs), which began after her last visit (a few weeks ago). She does endorse racing thoughts and a decreased need for sleep, but thinks she is “doing great.” |
| Duration (how long) | The origin of her psychiatric illness is not specified |
| Time relationships (frequency, constant or intermittent) | Robbin’s symptoms seem to be intermittent. She experiences both depression and mania, but they come and go. |
| Location | N/A |
| Radiation | N/A |
| Quality | N/A |
| Amount | N/A |
| Aggravated by what | Mania from the SSRIs, depression from life stresses (such as splitting from her partner Bobby). |
| Relieved by what |  |
| Associated with what |  |
| Attitude (what does the patient think is the problem, and how does he/she feel about it) | T0: Robbin feels sad, depressed, and tired since her partner left. She seems to attribute some of the way she feels towards this external stressor, but she does not open up/express much during the visit.  T1: Unlike in her initial visit, Robbin seems a bit more curious and contemplative regarding the issues that are causing her depression. While she is still sad over her partner leaving, she also wonders if she is a good mom and how her depression is affecting her daughter. She is also more expressive about her anhedonia, stating that she is “just going through the motions.” She even endorses passive suicidal ideation during this visit. It seems as though Robbin is more aware of her depressed feelings and where they may be coming from.  T2: Robbin’s only current complaint is that she needs her current visit to wrap up because she needs to get back to work. She has a lot of things to do and cannot be bothered. From a clinician’s point of view, she has switched into mania (possibly due to the SSRIs), which began after her last visit (a few weeks ago). She does endorse racing thoughts and a decreased need for sleep, but thinks she is “doing great.” |
| Overall course |  |
| REVIEW OF SYSTEMS: Significant positives and negatives | |
| Past medical history |  |
| Medication allergies (Name and reaction) | NKDA |
| Environmental allergies (Name and reaction) | None |
| Illnesses | None |
| Vaccinations | None |
| Surgeries | None |
| Accidents/ injuries/ trauma | None |
| Hospitalization |  |
| Inclusive sexual and reproductive history | |
| Sexual practices  Sexual partners  Protection: Use of safer sex practices  Use of birth control if appropriate  Risk of intimate partner violence | Has sexual intercourse with women  Not Specified, but just separated from her partner Bobby  Not specified  N/A  Not specified |
| Ob/GYN HISTORY | Age of onset of menses: Not specified  Age of menopause: Not specified  Number of pregnancies: Not specified  Number of live births: Not specified  Number of miscarriages: Not specified  Number of abortions: Not specified |
| Medications | Prescription/dose/reason: Zoloft for her recent depression. Dose is not specified  Over the counter/dose/reason: None  Herbs/supplements/dose/reason: None  Other: |
| Immunizations | - Tetanus - Flu - Hepatitis - Pneumovax - HPV - Other |
| Tobacco products:   - Cigarettes - Cigar - Pipe - Chew - E-cigarettes | - Never - Past- year started/year quit - Current   - Quantity   - # of years |
| Alcohol   - Beer - Wine - Liquor - Other | - Never - Past- year started/year quit - Current   - Quantity   - # of years |
| Drugs   - Weed - Cocaine - Heroin - Meth - Other - IV - Inhalants - Other | - Never - Past- year started/year quit - Current   - Quantity - # of years |
| Diet (describe) | Not Specified |
| Exercise (describe) | Not Specified |
| List any other important social history or information important to this case | Her partner Bobby left her a few weeks ago and she is a single mother with a 16 year old daughter. |
| Family history |  |
| Mother, Father, Siblings, Grandparents, and other significant findings. | Not specified |
| Physical Exam- List exam maneuvers expected for this case and any abnormal findings that SP will simulate. (tenderness, hyper-hypo reflex, rebound, weakness etc. )  None | |
| PHYSICAL EXAM FINDINGS |  |
| 1) Written in layman’s terms |  |
| 2) General appearance- affect, appearance, position of patient at opening (i.e. sitting, laying down, holding abdomen etc.) | T0: Slightly disheveled and dressed very casually (grey sweatshirt with her hood on, jeans, and a scarf). The visit begins with her sitting on the couch with her hood on, looking down at the floor. She often crosses her arms, giving off a feeling of being closed off.  She is flat, exhibiting psychomotor retardation, but is pleasant and cooperative.  T1: Groomed and well-dressed (grey long sleeve Henley and blue jeans). She has improved eye contact without any psychomotor retardation. The visit begins with her sitting on the couch, being very cooperative and pleasant. Robbin continues to exhibits a constricted range of affect (although improves) and seems a bit tired.  T2: Abnormal appearance (wearing a lot of makeup, a bright multi-colored button-down shirt, a tie, and colorful plastic hair clips in her hair). The visit begins with her sitting on the couch in a very restless manner. She is shaking her legs up and down, screaming that she wants the therapist to hurry up. Robbin is very labile throughout the visit, constantly switching back and forth between euphoric and irritable. Later in the visit, she paces up and down the room, trying to flirt with the therapist. |
| 3) Vital signs | Not specified |
| 4) Specific findings and affect | T0: Robbin has a very flat affect. She is answering the therapist with very few words and never seems to be expanding on the question. She lacks good eye contact and exhibits psychomotor retardation.  T1: Robbin is more expressive than her initial visit, but still seems a bit constricted in her affect. She displays anhedonia and endorses passive suicidality. She is more contemplative and energetic than her first visit, but is still tired and depressed.  T2: Robbin is very labile throughout the visit, constantly switching back and forth between euphoric and irritable. She has pressured speech and racing thoughts, constantly going on tangents. |
| 5) Response to certain physical movements | Not specified |
| DIAGNOSIS AND DIFFERENTIAL |  |
| Diagnosis with support from positive and negative history and PE findings | Robbins seems to be suffering from a mood disorder, most likely bipolar disorder (BD). In her first visit, she is very depressed, exhibiting psychomotor retardation, a flat affect, hypersomnia, and extreme sadness. In her second visit, her physical energy is better, but she continues to display depressive symptoms (anhedonia, fatigue, guilt, passive SI, etc.). It ispossible that during the second visit (T1), Robbin is displaying some mixed symptoms, which is allowing her to have enough physical energy to contemplate what is making her so depressed and suicidal. During her third visit, she exhibits grandiose delusions, pressured speech, mood lability, and, inappropriate sexual behavior. She also endorses euphoria and a decreased need for sleep. The switch to mania that is seen during this visit (due to the SSRI) is a good indicator that Robbin suffers from BD. |
| Differential with support from positive and negative history and PE findings | The differential diagnosis in this case includes conditions that may have manic-like symptoms, including organic mood disorders, such as endocrine or metabolic conditions, drug intoxications, and tumors. Although these are all unlikely. |
| MANAGEMENT OR DIAGNOSITIC PLAN | Thinking that Robbin suffers from depression, the therapist did the right thing by prescribing her an SSRI. During her second visit (after taking the SSRI for a few weeks), she seems to be improving physically, but begins endorsing suicidal ideation. Being that it was passive SI, the therapist once again did the right thing by scheduling another visit shortly there-after. Once Robbin switched into mania, it is important for her SSRIs to be stopped immediately, an anti-psychotic to be started (for acute purposes), and a mood-stabilizer, such as lithium, should be prescribed for maintenance. Due to the severe nature of her mania, it is possible that Robbin may need to be hospitalized (her suicidality and risky behavior should be assessed further). |
| PROFESSIONALISM ISSUES OR CHALLENGES: |  |
