## Supplementary material for "Human vs AI Clinical Assessment: Benchmarking a Multimodal Foundation Model Against Multi-Center Expert Judgment on the Mental Status Examination": case scripts, prompts and other supplements.: C. Case Script Karthik Schizophrenia.docx

**Appendix C**
Karthik (Schizophrenia): Standardized Patient Case

Date: 2/2019

Primary Case Author: Doron Amsalem, MD, Andrés S. Martin, MD, MPH

Secondary Case Author: Asaf Jacobs, MS4, Robert Krause, DNP, APRN-BC

Standardized Patient Educator: Robert Krause, DNP, APRN-BC, Andrés S. Martin, MD, MPH

Name of Case: Karthik – Chronic Schizophrenia Scenario

Name of educational and or assessment activity: Mental status exam (MSE) video-based assessment tool

Patient Name: Karthik

Chief Complaint: **At his initial visit (T0):** Karthik suffers from chronic schizophrenia, but currently is experiencing minimal symptoms. He complains of a ‘noisy neighbor’ and asks the doctor to decrease his medication, complaining that it makes him too tired. But states that “he is doing good overall.” It is not readily apparent that there is anything substantially wrong with him – this could be a routine concern over a neighbor.

**T1 (3 months later):** Karthik states that “things are not as good as I would have hoped.” He has a lot of “racing thoughts and can’t focus on anything.” He states that about a month ago he thought someone was following him, but that he “is okay now.” Later in the conversation, he endorses suspicion that his noisy neighbor is following him. These various symptoms (racing thoughts, lack of concentration etc.) make the patient wonder if he should not have made medication changes after all.

**T2 (4 months after initial visit):** Karthik states that “everything has gotten worse since last time.” He fears that his neighbor is poisoning his water supply (by accessing the water pipes in the utility room of the building). The voices of his neighbor, and others, have gotten louder and are bothering him more often (“all day and all night’’). These voices are telling him to jump off the roof. He also complains of an inability to fall asleep and chronic shaking of his hand. Karthik regrets lowering his medication.

Most likely Diagnosis and Differential with rationale from history and/or physical exam: Karthik has suffered from chronic paranoid schizophrenia since his freshman year of college. He has been hospitalized in a psychiatric unit twice. His negative symptoms seem to be a bit more constant than his positive symptoms (i.e.: flat affect, social withdrawal, delayed speech etc.). His positive symptoms are more present during an exacerbation of his illness. By his last visit (T2), he displays auditory and visual hallucinations, ideas of reference, paranoid delusions, suicidal ideation, and response to internal stimuli. His thought process is tangential, with loosening of associations.

Challenge question: Can learners identify key elements in the mental status exam (MSE), and do their abilities vary according to level of clinical experience and to didactic interventions they are exposed to?

Domains: Check all that apply

- Professionalism
- Communication and Interpersonal skills
- Medical History
- Physical exam
- Shared Decision Making
- Patient Education

X	Clinical Reasoning

- Documentation
- Handoff
- Presentation

X	Other: psychiatric history and clinical exam

Type and level of learner: Medical, nursing and physician associate students; other learners for whom competence on the MSE could be relevant (p. ex. psychiatric technicians, social workers, supporting staff at psychiatric clinics).

Case Objectives: please list specific objectives for each of the domains you have checked above:

1. Identify key components of the MSE, specifically those pertaining to chronic schizophrenia

2. Differentiate negative and positive symptoms and their degrees of severity

3. Distinguish between different types of delusions and thought process abnormalities that are often seen in psychosis/schizophrenia.

| SETTING: outpatient, in patient, ED, home, nursing home, rehab, group etc. | Outpatient clinic |
| --- | --- |
| PATIENT PROFILE: Information about the “patient” that helps select an SP and helps the learner get an understanding of them as a person. SP will know more information about the patient than learner will ever ask but allows SP to portray a fully developed patient personality. If none of the items below are particulars for the case please write “all may be used.” | |
| Age range | ~ 22 years old |
| Religious/spiritual background | Not specified |
| Sex (e.g., male, female, intersex, transwoman, transman) | Male |
| Sexual Orientation (e.g., heterosexual, lesbian, gay, bisexual, pansexual, queer, asexual) | Heterosexual |
| Gender expression (e.g., man, woman, gender queer) | Man |
| Race/ethnicity: | Indian |
| Physical description (e.g., BMI, height range) | 4’8”, BMI: 18-22 |
| Physical limitations | None |
| Patient appearance (e.g., disheveled, hospital gown, business casual, casual) | T0: groomed and well-dressed (jeans, white v-neck undershirt, black collared jacket).  T1: Slightly disheveled (messy hair, worn out grey t-shirt, jeans)  T2: Disheveled (messy hair, unwashed clothes, wearing a hat and sunglasses, or headphones |
| Moulage + location (e.g., none, bruises, scars, body piercing, tattoos) | None |
| Affect (e.g., pleasant, cooperative) | T0: Flat with minimal facial movements, tense and tired but cooperative  T1: Tense and avoids eye contact, slightly suspicious  T2: Tense with poor eye contact and akathisia |
| Family group (e.g., who is family, who they live with) | Lives alone in a small apartment. He is the youngest of four (2 brothers, one sister). Attends twice-a-month dinners with his siblings and mother (who he is close with). |
| Education | Sophomore in college |
| Level of health literacy | Adherent to his medication and doctor visits. Like many patients suffering from psychosis, his insight into his condition varies. |
| Employment, if any - present and past, noting any current stresses | Been employed in a work-study program at a natural foods store for the last year. |
| Home/homeless - type of dwelling, number of stories, owned or rented | Lives in an apartment (rented) |
| Financial situation- any current stresses | Does not complain of any current financial stressors |
| Insurance Status (e.g., un/under/insured, public/private, HMO/PPO) | Not specified |
| Habits (i.e., diet, exercise, caffeine, smoking, alcohol, drugs) | Not specified |
| Activities (i.e., hobbies, sports, clubs, friends) | Karthik has acquaintances at work, where he is well liked, but has had no close friends or romantic attachments. He is content keeping up with his siblings and mother. |
| Typical day - what is the usual daily routine | Spends a lot of time at work and at his apartment. Does not have many friends, but is close with his family. Keeps his days pretty simple/routine in order to stay grounded in reality. Lately, has been increasingly confused, experiencing perceptual disturbances. |

| CASE INFORMATION | |
| --- | --- |
| Chief Concern: What the patient will say when greeted by the student. The patient’s primary reason for seeking medical care often stated in his/own words. | T0: “I am doing okay. Things are going pretty well.”  (He suffers from chronic schizophrenia, but currently is experiencing minimal symptoms. He complains of a ‘noisy neighbor’ and asks the doctor to decrease his medication, complaining that it makes him too tired. But states that “he is doing good overall.”)  T1: “Things are not as good as I would have hoped. Things are worse than last time. I wonder if I should have not changed my medication after all.”  (He has a lot of “racing thoughts and can’t focus on anything.” He states that about a month ago he thought someone was following him, but that he “is okay now.” Later in the conversation, he endorses suspicion that his noisy neighbor is following him and wants to evict him from the building. These various symptoms (racing thoughts, lack of concentration etc.) make the patient wonder if he should not have made medication changes after all.)  T2: “Everything has gotten worse since last time. I think I should come down on my medication.”  (Karthik worries that his neighbor is trying to poison him. The voices he hears are becoming more intense and frequent. They are telling him to jump off the roof and are disturbing him. He regrets lowering his medication.) |
| Additional Concerns: Other, if any, concerns the patient has today (i.e., symptoms, requests, expectations, etc.) that will become part of set agenda. | T0: His main request is to decrease the dosage of his medication, complaining that it makes him too tired. He also mentions having a noisy neighbor, but does not ask for any help regarding that problem. Other than those complaints, he is experiencing minimal symptoms and feels that he is doing well.  T1: He complains of racing thoughts and a lack of concentration. He is concerned that his noisy neighbor is following him and wants to evict him from the building. He wonders if he should not have made a medication change.  T2: Karthik worries that his neighbor is trying to poison him. The voices he hears are becoming more intense and frequent. They are telling him to jump off the roof and are disturbing him. He regrets lowering his medication. |
| THE PATIENT STORY: The SP will be asked to tell their symptom story and the personal and emotion impact for each of their concerns. You will want to write this is the patient voice. The symptom story should be able to answer this question: “Tell me more about [chief concern/additional concern], starting at the beginning and bringing me up to now.”  The personal context should be able to answer questions concerning the broader personal/psychosocial context of symptoms, especially the patient beliefs/attributions.  The emotional context should be able to ask how are you doing with this, how does this make you feel, how has this affected you emotionally? IMPACT: How has this affected your life? How has this been for your family? | The conversation between Karthik and his doctor takes place at an outpatient clinic over three visits/appointments. During his first visit **(baseline, T0)**, Karthik present with **‘minimal symptoms’**; he is cooperative and in a good mood. During the second visit, three months later **(T1)**, Karthik experiences an exacerbation of his disease, presenting as anxious and suspicious **(i.e.: ‘medium symptoms’)**. He has subtle paranoid thoughts, not immediately apparent, and no hallucinations. He is trying to hide his thoughts and feelings and shares only part of what he is going through. His third visit takes place a month later **(T2)**, with Karthik experiencing **‘severe symptoms’**, including overtly paranoid thoughts and hallucinations.  First Visit (baseline, T0): Karthik arrives at his doctor’s office and states that he is “doing pretty well.” He is looking forward to having his bi-monthly dinner with his family, but prefers “to avoid other people.” Karthik is happy with his current job at the health store: “I don’t have to interact with too many people. I can just make sure everything is on the shelf.” He does complain of a noisy neighbor, who he “hears at all hours of the night.” He endorses having “mood swings here and there,” but states that it is mainly due to his “annoying neighbor.” Karthik feels “good overall,” but does request to have the dosage of his medication lowered. “It makes me so tired. It slows me down during the day and I want to be at my best.” Overall, Karthik displays a bit of a **flat affect**, his facial movements are minimal and his blinking is reduced in frequency, making his gaze seem like he is staring. He also twirls his hair occasionally (**slight mannerism)**. Karthik is oriented to time and place, has insight into his situation and has fair judgement.  T1: Karthik arrives at his doctor’s office, 3 months after their initial visit, and states that “things are not as good as [he] would have hoped. They are worse than last time.” He is **disheveled** (messy hair and dirty shirt), displaying minimal eye contact, and **psychomotor retardation** (his movements are slower than the first visit). He complains of **racing thoughts** and an inability to concentrate, which started “about a month ago.” Karthik endorses suspicion that his neighbor is following him around and trying to evict him from the building: “My neighbor is still very annoying. I heard him through the wall and I think he is trying to get rid of me. I think he might be following me around.” The therapist asks him why he thinks his neighbor is following him and responds by saying that he saw his neighbor standing over the staircase of their apartment building, staring at him. He also endorses seeing his neighbor at the store he works at, “right down the aisle staring at me.” Moreover, Karthik thinks the neighbor is spreading rumors about him. “He was talking to my neighbor Marc and then the next day, Marc looked at me like I did something wrong to him.” The therapist than asks him again, “so you think he is following you?” He responds, “it just makes sense.” Throughout the conversation, Karthik has a **prolonged latency to response, decreased prosody** and a **general delayed/slowed speech**. During this visit, he expresses **paranoid delusions** (i.e.: neighbor following him) and **ideas of reference** (neighbor talking about him to other tenants). Seeing his neighbor at his store and hearing his neighbor through his walls can suggest **auditory and visual hallucinations**. He is oriented to time and place, but lacks insight into his situation.  T2: Karthik arrives at his doctor’s office one month after their last visit and states that “everything has gotten worse since last time.” He arrives **disheveled**, with messy hair and dirty clothing, wearing a wool hat and sunglasses. When he is asked to take his sunglasses off he responds, “are we alone here?” He then looks around the room, as if he is suspicious of his surroundings and is **internally preoccupied**. He displays **bradykinesia** (slowness of movements; latency to respond to questions) as well as **akathisia** (involuntary movements, restlessness, discomfort in his chair (i.e.: in being in ‘his own skin). Shortly after he endorses fear that his neighbor is trying to poison his water supply (by accessing the water pipes in the utility room of the building). Throughout the conversation he displays **loose associations** and **tangential speech**, as well as at least one **neologism**: “The utility room where the water pipes are is the ‘pupility’ room, not the popularity room.” Moreover, Karthik’s **auditory hallucinations** become more frequent and more disturbing. “Sometimes I can hear a lot of them talking, sometimes all the way down in the basement.” He begins to develop **suicidal ideation**, hearing voices telling him to kill himself. “I’m hearing voices all day. Sometimes they tell me to jump off the roof, but I don’t do that.” His **delusions of paranoia** and **delusions of reference** strengthen as well (he is now fully convinced that his neighbor wants to kill him). Karthik’s symptoms have become severe; he is no longer in touch with reality. |
| HISTORY OF PRESENT ILLNESS: Although some of the HPI will be given in the patient’s symptom story, the learners will expand the story during the direct question section. Below describe the detailed history, usually about the chief concern, which the student must develop in order to make a useful assessment of the problem: | |
| Onset (when; gradual or sudden) | gradual |
| Setting (what was going on or where was patient when symptoms first noticed?) | T0: Karthik was first diagnosed with schizophrenia as a college freshman and has been admitted to an inpatient unit a few times in his early 20’s. During his initial visit, he complains of medication side effects (being too tired) and a noisy neighbor, but overall is feeling good.  T1: Karthik noticed an exacerbation of his symptoms 1 month prior to his second visit. He began experiencing racing thoughts and an inability to concentrate. Around this same time, he began thinking that his neighbor was following him and trying to get him evicted from the building. All of these symptoms are making him question his decision to change his medication.  T2: Karthik’s symptoms have worsened since his last visit (a month ago). He is now fully convinced that his neighbor is trying to kill him. The voices he hears have become more frequent and more disturbing, telling him to kill himself. He is very internally preoccupied and paranoid. He is hoping to increase his medication. |
| Duration (how long) | His symptoms have been present on and off since his freshman year in college. He has not been hospitalized in the last 5 years. |
| Time relationships (frequency, constant or intermittent) | Karthik’s negative symptoms seem to be a bit more constant than his positive ones. He has intermittent positive symptoms (psychosis, delusions, etc.), but has been quite stable for the last 5 years. His symptoms worsen a few months after his initial visit. |
| Location | N/A |
| Radiation | N/A |
| Quality | N/A |
| Amount | N/A |
| Aggravated by what |  |
| Relieved by what |  |
| Associated with what |  |
| Attitude (what does the patient think is the problem, and how does he/she feel about it) | T0: Patient has insight into his chronic disease and is adherent to his medication and routine check-ups with his therapist. He does complain that his medication is making him tired and believes that he would be at his best with a lower dosage.  T1: Karthik partially understands that his racing thoughts and lack of concentration are due to the change in his medication, but also believes that a lot of his current stress is due to his neighbor. He is convinced that his neighbor is trying to evict him from his apartment and that he talks badly about him to other neighbors. He is becoming very suspicious and paranoid, with slight insight into his condition.  T2: Karthik believes that his neighbor is a big source of his current problems. He is convinced that his neighbor is poisoning him. He hears a lot of voices that tell him to kill himself, but he never questions if they are real. He continues to question his medication change, but does not seem to blame his perceptual disturbances on the change (i.e.: thinks they are real). He is very paranoid and seems uncomfortable/unsafe in the doctor’s office. |
| Overall course |  |
| REVIEW OF SYSTEMS: Significant positives and negatives | |
| Past medical history |  |
| Medication allergies (Name and reaction) | NKDA |
| Environmental allergies (Name and reaction) | None |
| Illnesses | None |
| Vaccinations | None |
| Surgeries | None |
| Accidents/ injuries/ trauma | None |
| Hospitalization | A few psychiatric inpatient hospitalizations in his 20’s (some were involuntary). |
| Inclusive sexual and reproductive history | |
| Sexual practices  Sexual partners  Protection: Use of safer sex practices  Use of birth control if appropriate  Risk of intimate partner violence | Not specified  Not specified  Not specified  N/A  Not specified |
| Ob/GYN HISTORY | Age of onset of menses N/A  Age of menopause N/A  Number of pregnancies N/A  Number of live births N/A  Number of miscarriages N/A  Number of abortions N/A |
| Medications | Prescription/dose/reason  Over the counter/dose/reason  Herbs/supplements/dose/reason  Other: |
| Immunizations | - Tetanus - Flu - Hepatitis - Pneumovax - HPV - Other |
| Tobacco products:   - Cigarettes - Cigar - Pipe - Chew - E-cigarettes | - Never - Past- year started/year quit - Current   - Quantity   - # of years |
| Alcohol   - Beer - Wine - Liquor - Other | - Never - Past- year started/year quit - Current   - Quantity   - # of years |
| Drugs   - Weed - Cocaine - Heroin - Meth - Other - IV - Inhalants - Other | - Never - Past- year started/year quit - Current   - Quantity - # of years |
| Diet (describe) | Not specified |
| Exercise (describe) | Not specified |
| List any other important social history or information important to this case | Patient has bi-monthly dinners with his family, but has had no close friends or romantic attachments; he prefers to avoid people and to have minimal interaction with them. He as a small apartment where he lives on his own. |
| Family history |  |
| Mother, Father, Siblings, Grandparents, and other significant findings. | Not specified |
| Physical Exam- List exam maneuvers expected for this case and any abnormal findings that SP will simulate. (tenderness, hyper-hypo reflex, rebound, weakness etc. )  None | |
| PHYSICAL EXAM FINDINGS |  |
| 1) Written in layman’s terms |  |
| 2) General appearance- affect, appearance, position of patient at opening (i.e. sitting, laying down, holding abdomen etc.) | T0: Groomed and well-dressed (jeans, white v-neck undershirt, black collared jacket), sitting in a chair at the doctor’s office. Karthik displays a bit of a flat affect, his facial movements are minimal and his blinking is reduced in frequency, making his gaze seem like he is staring. He also twirls his hair occasionally (slight mannerism).  T1: Slightly disheveled (messy hair, worn out grey t-shirt, jeans), sitting in a chair at the doctor’s office. Karthik displays a flat affect, with minimal eye contact and delayed/slowed speech. In addition, he has decreased prosody with a prolonged latency to response.  T2: He is disheveled, with messy hair and dirty clothing, wearing a wool hat and sunglasses. He looks around the room, as if he is suspicious of his surroundings and is internally preoccupied. He displays bradykinesia (slowness of movements; latency to respond to questions) as well as akathisia (involuntary movements, restlessness, and discomfort in his chair). |
| 3) Vital signs | Not specified |
| 4) Specific findings and affect | T0: Karthik has minimal facial movements with reduced blinking. He displays a flat affect and a slight mannerism (twirling his hair).  T1: Karthik displays a flat affect with minimal facial movements and eye-contact. Throughout the conversation he exhibits delayed speech with prolonged latency to response.  T2: Karthik continues to display a flat affect and avoids eye contact. He then looks around the room, as if he is suspicious of his surroundings and is internally preoccupied. He displays bradykinesia (slowness of movements; latency to respond to questions) as well as akathisia (involuntary movements, restlessness, and discomfort in his chair. Throughout the conversation he uses neologisms, loose associations, and tangential speech: |
| 5) Response to certain physical movements | N/A |
| DIAGNOSIS AND DIFFERENTIAL |  |
| Diagnosis with support from positive and negative history and PE findings | Karthik suffers from chronic schizophrenia since his freshman year of college. He has been hospitalized in a psychiatric unit a few times in his 20’s. His negative symptoms seem to be a bit more constant than his positive symptoms (i.e.: flat affect, social withdrawal, delayed speech etc.). His positive symptoms are more present during an exacerbation of his illness. By his last visit (T2), he displays auditory and visual hallucinations, ideas of reference, paranoid delusions, suicidal ideation, and response to internal stimuli. His thought process is tangential with loosening of associations. |
| Differential with support from positive and negative history and PE findings | Karthik may suffer from a psychotic disorder of unspecified origin. As listed above, he suffers from many positive and negative symptoms that would suggest a psychotic disorder. His lack of sleep and racing thoughts make bipolar disorder another possible diagnosis. |
| MANAGEMENT OR DIAGNOSITIC PLAN | T0: Consider lowering his medication and closely monitor for an exacerbation of symptoms (i.e.: schedule a follow-up visit).  T1: Consider increasing his medications back to the original dose and schedule a follow-up visit to assess his condition.  T2: Karthik is exhibiting suicidal ideation and lacks good reality testing. His medication should be increased and an involuntary psychiatric hospitalization may be necessary. |
| PROFESSIONALISM ISSUES OR CHALLENGES: |  |
