## Supplementary material for "Human vs AI Clinical Assessment: Benchmarking a Multimodal Foundation Model Against Multi-Center Expert Judgment on the Mental Status Examination": case scripts, prompts and other supplements.: M. Scoring Sheet.docx

| **APPENDIX M** |  |  |  |  |  |  |  |  |  |  |
| --- | --- | --- | --- | --- | --- | --- | --- | --- | --- | --- |
| **Blank scoring sheet for MSE components** | | | | | | | |  |  |  |
|  | **Diagnostic category and time point** | | | | | | | | | **Total items** |
|  | **Schizophrenia (Karthik)** | | | **Bipolar disorder (Robbin)** | | | **OCD (Ben)** | | |  |
| **MSE component** | **1** | **2** | **3** | **1** | **2** | **3** | **1** | **2** | **3** |  |
| Sign or symptom |  |  |  |  |  |  |  |  |  |  |
| **Appearance, Behavior and Cooperation** |  |  |  |  |  |  |  |  |  |  |
| Grooming and hygiene (abnormal) |  |  |  |  |  |  |  |  |  |  |
| Eye contact (abnormal) |  |  |  |  |  |  |  |  |  |  |
| Psychomotor retardation / bradykinesia |  |  |  |  |  |  |  |  |  |  |
| Psychomotor activation / akathisia |  |  |  |  |  |  |  |  |  |  |
| Physical proximity / distance (abnormal) |  |  |  |  |  |  |  |  |  |  |
| Stereotypies or mannerisms |  |  |  |  |  |  |  |  |  |  |
| Tics |  |  |  |  |  |  |  |  |  |  |
| Tremor |  |  |  |  |  |  |  |  |  |  |
| **Speech** |  |  |  |  |  |  |  |  |  |  |
| Slowed/delayed |  |  |  |  |  |  |  |  |  |  |
| Prolonged latency to response |  |  |  |  |  |  |  |  |  |  |
| Prosody (decreased) |  |  |  |  |  |  |  |  |  |  |
| Pressured |  |  |  |  |  |  |  |  |  |  |
| **Thought Process A: Coherence** |  |  |  |  |  |  |  |  |  |  |
| Circumstantial / tangential |  |  |  |  |  |  |  |  |  |  |
| Loosening of associations |  |  |  |  |  |  |  |  |  |  |
| Neologism |  |  |  |  |  |  |  |  |  |  |
| Word salad |  |  |  |  |  |  |  |  |  |  |
| **Thought Process B: Speed** |  |  |  |  |  |  |  |  |  |  |
| Mutism |  |  |  |  |  |  |  |  |  |  |
| Thought blocking |  |  |  |  |  |  |  |  |  |  |
| Racing thoughts |  |  |  |  |  |  |  |  |  |  |
| Flight of ideas |  |  |  |  |  |  |  |  |  |  |
| **Thought Content A: Delusions** |  |  |  |  |  |  |  |  |  |  |
| Paranoid |  |  |  |  |  |  |  |  |  |  |
| Of reference |  |  |  |  |  |  |  |  |  |  |
| Somatic |  |  |  |  |  |  |  |  |  |  |
| Grandiose |  |  |  |  |  |  |  |  |  |  |
| Of control |  |  |  |  |  |  |  |  |  |  |
| **Thought Content B: Obsessions** |  |  |  |  |  |  |  |  |  |  |
| Contamination / environmental concerns |  |  |  |  |  |  |  |  |  |  |
| Somatic / illness |  |  |  |  |  |  |  |  |  |  |
| Aggressive / sexual / forbidden thoughts / loss of control |  |  |  |  |  |  |  |  |  |  |
| Pathological doubting |  |  |  |  |  |  |  |  |  |  |
| **Thought Content C: Compulsions** |  |  |  |  |  |  |  |  |  |  |
| Cleaning / washing |  |  |  |  |  |  |  |  |  |  |
| Checking / seeking reassurance |  |  |  |  |  |  |  |  |  |  |
| Ordering / organizing |  |  |  |  |  |  |  |  |  |  |
| Saving / Hoarding |  |  |  |  |  |  |  |  |  |  |
| **Affect and Mood** |  |  |  |  |  |  |  |  |  |  |
| Constricted range of affect, flat |  |  |  |  |  |  |  |  |  |  |
| Anhedonic |  |  |  |  |  |  |  |  |  |  |
| Expansive range of affect, wide |  |  |  |  |  |  |  |  |  |  |
| Inappropriate and/or labile affect |  |  |  |  |  |  |  |  |  |  |
| Euphoric / hypomanic / manic |  |  |  |  |  |  |  |  |  |  |
| **Perceptions** |  |  |  |  |  |  |  |  |  |  |
| Auditory hallucination |  |  |  |  |  |  |  |  |  |  |
| Visual hallucination / illusion / misperception |  |  |  |  |  |  |  |  |  |  |
| Tactile hallucination |  |  |  |  |  |  |  |  |  |  |
| Responding to internal stimuli |  |  |  |  |  |  |  |  |  |  |
| **Suicidality** |  |  |  |  |  |  |  |  |  |  |
| Suicidal ideation (active or passive) |  |  |  |  |  |  |  |  |  |  |
| Homicidal ideation |  |  |  |  |  |  |  |  |  |  |
| **Total** |  |  |  |  |  |  |  |  |  |  |
| OCD = obsessive-compulsive disorder | | | | | | | | | |  |
