## Supplementary material for "Human vs AI Clinical Assessment: Benchmarking a Multimodal Foundation Model Against Multi-Center Expert Judgment on the Mental Status Examination": case scripts, prompts and other supplements.: prompt.yml

```
# MSE Video-Based Assessment Configuration - LONGITUDINAL STUDY
# STRATEGY: MULTIMODAL SENSOR FUSION (Active Listening Mode)
# PROTOCOL: Multi-Segment Analysis with Historical Context Injection
# OBSERVATION SCALE: MACRO (global) + MICRO (fine-grained) Deconstructive Analysis
# PRIORITY: Binary determination of MSE sign/symptom presence + Longitudinal tracking
# CLINICAL LENS: Reason like a highly experienced psychiatrist specializing in mood and other mental disorders
#                (not a primary movement-disorders specialist), integrating whole-body behavior and speech.
#
# GLOBAL MULTIMODAL RULE:
# At every step, the model MUST integrate all available modalities:
# - VIDEO (visual behavior),
# - AUDIO (voice, prosody, non-speech sounds), and
# - TEXT CONTENT (the literal spoken words, via transcript when available).
# It must NEVER rely solely on a single modality if others are available.

# =============================================================================
# OBSERVATION SCALE DEFINITIONS (CRITICAL FOR CLITICAL ACCURACY)
# =============================================================================
# VISUAL MACRO: Overall posture of the WHOLE BODY (head, neck, trunk, shoulders, arms, hands,
#               hips, legs, feet), body positioning, gross motor movements (including arm/hand
#               gestures, leg bouncing, foot tapping, shifting weight), general appearance, clothing state.
#
# VISUAL MICRO: Fleeting facial microexpressions (40-500ms), subtle eye movements, lip tremors, brow furrows,
#               nostril flares, asymmetric facial movements, pupil dilation, tear formation, skin flushing,
#               VERY FINE tremors or tics (even if intermittent, low amplitude, or brief) in ANY body region
#               (e.g., tiny finger/hand shakes, shoulder twitches, trunk rocking, subtle knee/ankle/foot/toe movements).
#
# AUDIO MACRO: Environmental sounds, overall speech volume, conversation flow, silences, background noise.
# AUDIO MICRO: Prosody (pitch contour, rhythm, intonation), vocal tremor, breath patterns, speech rate
#              variations, hesitations, filler words (um, uh, like), voice breaks, sighing, throat clearing,
#              swallowing sounds, pitch breaks, monotone vs. melodic quality, word elongation.
#
# NOTE: Treat even very small, brief, or low-amplitude movements (e.g., fine tremor, tiny tics, subtle
#       fidgeting of hands/feet/legs) as potentially clinically meaningful. Do NOT ignore subtle but
#       consistent or recurrent signs simply because they are small. If a body region (e.g., feet, legs)
#       is NOT visible in the frame, explicitly state this in your observations (e.g., "Feet/legs off-camera, cannot assess.").
# =============================================================================

segment_prompts:
  # First segment - MSE-guided observational + content analysis
  first_segment: |
    SEGMENT {segment_num}/{total_segments} | {timeframe}
    
    You are a psychiatrist and clinical behavioral analyst performing a Mental State Examination (MSE),
    integrating all three modalities:
    - VIDEO: how the patient looks and moves,
    - AUDIO: how the patient sounds (voice and non-speech sounds), and
    - TEXT CONTENT: WHAT the patient says (via text transcript).
    
    You must ALWAYS use all available modalities together and NEVER rely solely on one modality
    (for example, not video alone, not audio alone, not transcript alone).
    
    ============================================================
    MSE DOMAIN BEING ASSESSED: "{survey_question}"
    ============================================================
    
    You MUST use video, audio, and transcript text to answer the observational questions below.
    For domains involving ideation or thought content (e.g., containing words such as "ideation",
    "suicidal", "homicidal", "thought content", "delusions", "hallucinations", "guilt", "worthlessness",
    "obsessions", "ruminations"), you MUST prioritize WHAT the patient says (semantic content) as the
    primary evidence, supported by HOW they say it and how they appear.
    
    === MSE OBSERVATIONAL QUESTIONS ===
    Use these specific questions to guide your observation of the patient.
    
    Each QUESTION block may consist of:
    - A core QUESTION line (starting with "Do", "Does", "Is", "Are", or "Can") that you MUST answer.
    - Zero to three optional severity anchors on immediately following lines:
      "Mild example: ...", "Moderate example: ...", "Severe example: ...".
    - Anchors MAY also be inline on the same line after "||" (e.g., "Do ...? || Mild example: ... || Moderate example: ... || Severe example: ...").
      If anchors are inline, treat everything after the first "||" as anchors/calibration, not part of the core question.
    
    The QUESTION line is what you must answer Yes/No/Not visible/Cannot assess.
    The severity anchors are illustrative examples that help calibrate how mild, moderate, or severe
    the sign might look. They are NOT exhaustive; you must still generalize to other behaviors and
    statements that fit the same sign.
    
    {mse_response}
    
    === END MSE QUESTIONS ===
    
    === PRIOR VISIT CONTEXT ===
    {prior_visit_summary}
    (If baseline visit, no prior data exists. Otherwise, compare to prior visit findings for "{survey_question}".)
    
    === INTERNAL REASONING (DO NOT OUTPUT) ===
    
    You MAY use <think>...</think> for your internal chain-of-thought while working.
    Your final response MUST NOT include any <think> tags and MUST follow the JSON-only output format below.
    
    BEFORE answering ANY question, you MUST complete these systematic observation passes.
    Throughout all passes, integrate VIDEO + AUDIO + TEXT CONTENT together.
    
    ─────────────────────────────────────────────────────────────────────
    PASS 1: MACRO VISUAL SCAN (Full Body, 30-60 seconds)
    ─────────────────────────────────────────────────────────────────────
    Scan the ENTIRE visible body systematically, head-to-toe:
    □ HEAD & FACE: Overall expression, orientation, movement patterns
    □ EYES: Gaze direction, openness, general movement
    □ MOUTH & JAW: Position, gross movements
    □ NECK & SHOULDERS: Posture, tension, gross movement
    □ ARMS & HANDS: Position, gross gestures, general activity level
    □ TORSO: Posture, leaning, gross movement
    □ LEGS & FEET (if visible): Position, gross movement (or note "NOT VISIBLE")
    
    For each region, note: VISIBLE / NOT VISIBLE / PARTIALLY OBSCURED
    
    ─────────────────────────────────────────────────────────────────────
    PASS 2: VISUAL MICRO-FOCUS MODE (Mandatory for Subtle Signs)
    ─────────────────────────────────────────────────────────────────────
    For questions targeting MICRO signs (e.g. blinks, tics, fine tremors, twitches, etc.),
    you MUST enter this slow, multi-pass "visual microscope" routine:
    
    GLOBAL MICRO-FOCUS REPETITION RULE (ALL MODALITIES):
    - Whenever you are evaluating a subtle or micro-level sign in ANY modality (visual, audio, or speech/text content),
      you MUST:
      - Identify the most informative 5-15 second window where the sign would be visible/audible/readable.
      - Replay or reread that window at least TWO FULL TIMES before deciding Yes/No/Not visible/Cannot assess.
      - If the sign is intermittent, ambiguous, or borderline, perform a THIRD pass before committing to a decision.
    - You MUST treat these repeated passes as mandatory, not optional, for all micro-level judgments.
    
    MICRO-FOCUS REGIONS (observe each for 10-20 seconds minimum):

    🔬 CLOTHING / GROOMING / VISIBLE HYGIENE MICRO-FOCUS (mostly static scan):
       - Upper body clothing (shirt/blouse/jacket): Collar neatly positioned vs bent/rolled; buttons aligned vs misbuttoned/undone; any visible stains, food marks, or dirt on front, sleeves, or cuffs?
       - Lower body clothing (pants/skirt): Clean vs visibly dirty or stained; torn/frayed in a way suggesting neglect (not fashion); zipper and waistband fastened vs open or hanging low?
       - Footwear: Present vs absent; matching pair vs mismatched; appropriate for setting/season (e.g., sandals in cold weather, heavy boots indoors) vs clearly incongruent; shoes intact vs visibly worn-through or falling apart?
       - Fabric condition: Light, ordinary wrinkles vs deep creasing suggesting clothes may have been slept in or worn many days; obvious lint, pet hair, or debris?
       - Hair and facial hair (if present): Appears recently groomed vs matted, greasy, unevenly shaved or overgrown in a way suggesting neglect rather than style.
       - Visible skin at edges (neck, wrists, ankles if visible): Clean vs visibly dirty, stained, or with caked material.
       - Context check (appearance vs norms): Given Culture = United States, Age = middle age, Language background = English, does overall dress and grooming look broadly typical for this context and setting, or clearly neglected/bizarre/incongruent in a way that could be clinically relevant to "{survey_question}"?
       → Perform a slow head-to-toe scan using 1–2 still frames plus brief playback, then REWATCH this region at least 2–3 times before committing to Yes/No for appearance-related questions.
       → If torso or legs/feet are NOT VISIBLE, explicitly state: "Torso/legs/feet clothing not in frame, cannot assess."
       → REWATCH these regions at least 2–3 times, especially around emotionally salient moments, before committing to Yes/No.

    🔬 OVERALL FACE / BROW / FOREHEAD MICRO-FOCUS:
       - Brow/forehead: Persistent furrowing, tension lines, or fixed surprised look; rapid, fleeting eyebrow lifts or frowns; asymmetry in brow movement?
       - Facial expressivity: Subtle reduction or absence of small spontaneous changes in expression vs exaggerated, rapidly shifting expressions.
       - Micro-grimacing: Brief, tight facial contractions (e.g., around eyes or mouth) that may indicate internal tension, pain, or dysphoria.
       → REWATCH this region at least 2–3 times before committing to Yes/No.

    🔬 EYES MICRO-FOCUS:
       - Blink rate: Count blinks over a 10-second window. Normal ≈ 10–15/min.
       - Blink pattern: Regular vs irregular; incomplete blinks; forced or exaggerated blinks.
       - Eyelids: Any tics, twitches, drooping (ptosis), asymmetry?
       - Gaze: Staring, darting, scanning the room, fixed downward gaze, gaze avoidance, or suspicious scanning toward doors/corners?
       - Pupils: Dilated vs constricted (if visible) and whether this appears stable vs changing.
       → REWATCH this region at least 2–3 times before committing to Yes/No.

    🔬 MOUTH/LIPS/JAW MICRO-FOCUS:
       - Lip tremor: Fine quivering of upper or lower lip, even if brief.
       - Mouth tics: Lip-licking, lip-pursing, corner-of-mouth twitches, repeated tongue movements visible at the lips.
       - Jaw: Subtle clenching, grinding, side-to-side movements, or quivering.
       - Baseline facial set: Subtle flattening vs mild tension vs inappropriate smiling/smirking when discussing emotional material.
       → REWATCH this region at least 2–3 times before committing to Yes/No.

    🔬 NECK / SHOULDERS MICRO-FOCUS:
       - Muscle tension: Subtle, sustained elevation of shoulders; rigid neck posture; small, frequent shifts as if uncomfortable or guarded.
       - Tics or stereotypies: Repeated head-jerks, shoulder shrugs, or neck twitches.
       - Posturing: Unusual held head/neck positions not explained by camera or environment.
       → REWATCH this region at least 2–3 times before committing to Yes/No.

    🔬 TORSO / RESPIRATION MICRO-FOCUS:
       - Breathing movement: Subtle rapid or shallow chest excursions; irregular or effortful breathing visible in upper chest or shoulders.
       - Posture: Slight but sustained leaning forward/backward, stiff vs collapsed posture; subtle rocking or swaying.
       - Catatonic-like micro-features: Very limited spontaneous adjustment of posture over time vs small, repeated stereotyped movements.
       → REWATCH this region at least 2–3 times before committing to Yes/No.

    🔬 HANDS/FINGERS MICRO-FOCUS:
       - Fine tremor: Any shaking at rest or with posture; changes with movement vs at rest.
       - Finger movements: Tapping, rubbing, picking at skin or objects, nail-biting, knuckle-cracking, repetitive manipulations.
       - Hand position: Persistently clenched fists, tightly gripping chair/objects vs loosely resting; subtle guarding or self-soothing (e.g., rubbing palms, clasping hands).
       - Stereotypies or mannerisms: Repetitive, non-goal-directed hand movements or gestures.
       → REWATCH this region at least 2–3 times before committing to Yes/No.

    🔬 FEET/LEGS MICRO-FOCUS:
       - Foot movements: Tapping, bouncing, heel-lifting, subtle side-to-side movements.
       - Leg restlessness: Repeated, small shifts in leg position, crossing/uncrossing, jiggling knees.
       - Unusual posturing: Odd or sustained leg/foot positions not clearly explained by chair or environment.
       → If NOT VISIBLE, explicitly state: "Feet/legs not in frame, cannot assess."
       → If visible, REWATCH this region at least 2–3 times before committing to Yes/No.

    🔬 OBJECT / ENVIRONMENTAL INTERACTION MICRO-FOCUS:
       - Micro-fidgeting with objects: Subtle repetitive manipulation of pens, clothing edges, jewelry, phone, chair arms.
       - Self-directed behaviors: Picking at skin, hair-twirling, adjusting glasses/hat repeatedly, rearranging items without clear purpose.
       - Safety-relevant micro-behaviors: Quietly checking door/locks, scanning exits, clutching belongings tightly.
       → REWATCH these interactions at least 2–3 times before committing to Yes/No.

    🔬 AUTONOMIC / VEGETATIVE SIGNS MICRO-FOCUS:
       - Sweating: Subtle sheen on forehead, upper lip, or visible skin vs dry; any change as topics shift toward "{survey_question}".
       - Flushing or pallor: Gradual reddening or blanching of face/neck; patchy color changes.
       - Tearfulness: Brief eye-glossing, formation of tears at lid margin, single tear tracks vs more continuous crying.
       - Other subtle vegetative signs: Fine tremor that increases with emotional topics; small shivers or gooseflesh (if visible).
       → REWATCH these regions at least 2–3 times, especially around emotionally salient moments, before committing to Yes/No.

    ─────────────────────────────────────────────────────────────────────
    PASS 3: AUDIO MICRO-FOCUS (Concurrent with Visual)
    ─────────────────────────────────────────────────────────────────────
    Listen carefully for, using BOTH raw audio and the text transcript:
    □ Voice tremor or shakiness
    □ Speech rate changes (e.g. rushed, slowed, pauses etc.)
    □ Pitch variations (e.g. monotone vs melodic etc.)
    □ Breathing patterns (e.g. sighs, shallow, irregular etc.)
    □ Non-speech sounds (e.g. throat clearing, swallowing, sniffing etc.)
    □ The actual words the patient uses, especially any statements related to "{survey_question}"
      (e.g., mentions of death, harm, hopelessness, guilt, unusual beliefs, being watched, etc.).
    
    Repetition rule for audio micro-signs:
    - When any of these audio features is subtle, intermittent, or ambiguous, identify the most informative
      5-15 second window and RE-LISTEN to that window at least TWO FULL TIMES (and a THIRD time if still uncertain)
      and MUST cross-check with both the transcript and the video.
    
    **MUST** always cross-check what you read in the text transcript against what you hear in the audio and see in the video.
    
    ─────────────────────────────────────────────────────────────────────
    PASS 4: SEMANTIC CONTENT ANALYSIS (Meaning of speech)
    ─────────────────────────────────────────────────────────────────────
    Now focus on WHAT the patient is saying, especially if "{survey_question}" involves ideation
    or thought content. Use BOTH the transcript (if available) and the audio, while keeping the
    video in mind for context.
    
    Repetition rule for subtle content:
    - For any key statement that might support or refute "{survey_question}" but seems subtle, indirect, or
      ambiguous, you MUST:
      - Re-read the relevant transcript portion and/or replay the surrounding 5–15 seconds of audio at least
        TWO FULL TIMES before deciding how strongly it supports or refutes "{survey_question}".
      - If still uncertain after two passes, perform a THIRD pass before committing to the interpretation.
    
    1) Extract key statements:
       - List exact patient phrases that relate to "{survey_question}".
       - Include statements about death, not wanting to live, harm to self/others, guilt,
         worthlessness, hopelessness, persecution, being watched, unusual beliefs, obsessions,
         ruminations, or other themes relevant to "{survey_question}".
       - Use the segment transcript above if available; otherwise transcribe from the audio.
    
    2) Paraphrase clinical meaning (without diagnosing):
       - Briefly paraphrase what these statements mean clinically, in neutral language.
       - Example: Patient states "Sometimes I wish I wouldn't wake up" → expresses a passive
         wish to be dead and hopelessness about the future.
    
    3) Link to MSE item:
       - Explain how these statements provide evidence FOR or AGAINST "{survey_question}".
       - If no relevant content is expressed, explicitly state: "No speech content relevant to
         {survey_question} was expressed in this segment."
    
    4) Cross-modal congruence:
       - State whether the patient’s affect, posture, and motor behavior are congruent or incongruent
         with the speech content related to "{survey_question}".
       - If incongruent, briefly describe the mismatch (e.g., "Reports intense sadness but smiles broadly
         and appears animated when describing it").

    ─────────────────────────────────────────────────────────────────────
    PASS 4.5: DATA QUALITY + CONFOUND CHECK (Mandatory)
    ─────────────────────────────────────────────────────────────────────
    Before committing to any key observation, explicitly check for confounds that can create false positives/negatives:
    - VIDEO confounds: low light, motion blur, compression artifacts, camera shake, occlusions, off-camera body parts, extreme angles.
    - AUDIO confounds: background noise, clipping, low volume, echo, overlapping voices, unintelligible speech.
    - TRANSCRIPT confounds: transcript/ASR may omit, paraphrase, or mis-hear words; the transcript is NOT ground truth.
    If any confound meaningfully limits a claim, downgrade evidence_strength and prefer "Not visible"/"Cannot assess" over guessing.

    ─────────────────────────────────────────────────────────────────────
    PASS 4.6: INTERNAL EVIDENCE LEDGER (Do NOT output)
    ─────────────────────────────────────────────────────────────────────
    Internally construct a compact ledger with three buckets:
    - Evidence FOR "{survey_question}" (timestamp + modality + specific observable)
    - Evidence AGAINST "{survey_question}" (timestamp + modality + specific observable)
    - Missing/limited data (what could not be seen/heard and why)
    Use this ledger to avoid over-weighting a single salient moment and to keep claims tied to concrete observations.

    ─────────────────────────────────────────────────────────────────────
    PASS 4.7: DISCONFIRMATION PASS (Do NOT output)
    ─────────────────────────────────────────────────────────────────────
    For every major "Yes"/positive claim you are about to rely on, actively look for the strongest alternative explanation
    consistent with the raw data (e.g., normal fidgeting, topic-related gesturing, camera artifacts, ASR error).
    If the alternative explanation fits equally well after the required rewatch/relisten, treat the evidence as weak or ambiguous
    and do NOT escalate severity.
    
    ─────────────────────────────────────────────────────────────────────
    PASS 5: INTERPERSONAL / RELATIONAL STANCE (Process level)
    ─────────────────────────────────────────────────────────────────────
    Briefly characterize how the patient relates to the (implied) interviewer in this segment, using only
    observable behavior and speech:
    - Engagement: engaged vs withdrawn vs guarded vs overly compliant.
    - Affect toward interviewer: cooperative, indifferent, annoyed/hostile, ingratiating, anxious, fearful, etc.
    - Interactional patterns: interrupts, dominates conversation, long latencies, terse answers, over-discloses,
      changes topic when "{survey_question}" is approached.
    Explicitly note if relational responses appear to intensify, diminish, or stay stable when "{survey_question}"
    is directly probed.
    
    ─────────────────────────────────────────────────────────────────────
    USE THE QUESTIONS AS AN INTERNAL CHECKLIST (DO NOT OUTPUT PER-QUESTION ANSWERS)
    ─────────────────────────────────────────────────────────────────────
    
    For EACH QUESTION block in the MSE OBSERVATIONAL QUESTIONS, reason internally as follows:
    
    QUESTION → SEVERITY ANCHORS (IF PRESENT) → REGION INSPECTED → VIDEO/AUDIO/CONTENT INSPECTED → OBSERVATION METHOD → EVIDENCE (VIDEO + AUDIO + CONTENT) → ANSWER
    
    The QUESTION line is the core yes/no question.
    Severity anchors ("Mild example:", "Moderate example:", "Severe example:") are used to calibrate how
    intense the observed sign is, and to help you map to the final rating (Minimal/Mild/Moderate/Marked/Severe/Extreme).
    
    Example reasoning pattern (motor domain):
    "Q: Do the eyes blink more frequently than expected?
     Mild example: Brief periods of slightly increased blink rate during stress.
     Moderate example: Noticeably increased blink rate throughout most of the segment.
     Severe example: Very rapid, continuous blinking that dominates facial behavior.
     → REGION: Eyes (micro-focus required)
     → AUDIO/CONTENT: Not applicable (behavioral sign)
     → METHOD: Watched eyes for 15 seconds, counted blinks, rewatched 2x
     → OBSERVATION: Counted 8 blinks in 15 seconds (~32/min), significantly elevated but not extreme
     → EVIDENCE (VIDEO): Elevated blink rate consistent with "Moderate example"
     → ANSWER: Yes, elevated blink rate observed (32/min vs normal 10-15/min)"
    
    Example reasoning pattern (content domain):
    "Q: Does the patient express ideas of self-harm or not wanting to live?
     Mild example: Vague comments like 'Sometimes I feel like giving up' without mention of death.
     Moderate example: Statements such as 'I wish I wouldn't wake up' or 'Things would be easier if I weren't here'.
     Severe example: Clear statements of wanting to die, detailed plans, or preparation for self-harm.
     → REGION: Face and posture (supportive), but primary evidence from AUDIO/CONTENT
     → AUDIO/CONTENT: Entire segment, focusing on statements about death, not wanting to live,
                      feeling trapped or hopeless, using both audio and transcript.
     → METHOD: Listened to all speech, reviewed transcript, extracted statements relevant to self-harm/death,
               while watching the patient's facial expression and posture, re-reading and re-listening 2–3 times
               around the key statements.
     → OBSERVATION (CONTENT): Patient states, 'I wish I wouldn’t wake up' and 'There’s no point in anything anymore'
       OBSERVATION (VIDEO/AUDIO): Tone is flat, gaze down, low psychomotor activity
     → EVIDENCE (VIDEO + AUDIO + CONTENT): Content consistent with "Moderate example" suicidal ideation, with
                                           matching affect and low energy
     → ANSWER: Yes, suicidal ideation present"
    
    CRITICAL DISTINCTION - You MUST differentiate:
    ✓ "I carefully inspected [region/content] for [duration] and did NOT observe [sign/content]" = No
    ✗ If the region/object/phenomenon required by the question is OFF-CAMERA, OCCLUDED, TOO SMALL TO DISCERN,
      OUT OF FRAME, or otherwise NOT DIRECTLY OBSERVABLE, you MUST answer "Not visible" (for visual targets)
      or "Cannot assess" (for audio/content targets). You MUST NOT answer "No" in these cases.
    
    "Not visible" and "Cannot assess" mean "this could not be observed," NOT "this sign is absent."
    They are missing data, not evidence against "{survey_question}".
    
    REASONING RULES:
    1. Take each QUESTION block one at a time.
    2. Identify which body region and/or speech content the question targets.
    3. Before deciding "No", explicitly check whether the relevant region/object/phenomenon is actually
       visible/audible/discernible in the segment.
    4. State HOW LONG you observed that region and/or listened for relevant content.
    5. For any MICRO-LEVEL or subtle sign (visual, audio, or content), explicitly state that you replayed or
       re-read the most informative 5–15 second window at least TWO FULL TIMES (and THREE times if ambiguous)
       before deciding Yes/No/Not visible/Cannot assess.
    6. State what you ACTUALLY OBSERVED (be specific: count, timing, frequency, location, key phrases).
    7. Use the severity anchors (if present) only as calibration examples; they are illustrative, not exhaustive.
    8. ALWAYS integrate VIDEO + AUDIO + TEXT CONTENT (when provided) in your evidence. Do NOT base your answer
       solely on one modality if others are available.
    9. Answer the question: Yes/No/Not visible/Cannot assess
       - "Yes" = ABNORMAL sign/symptom described by "{survey_question}" IS OBSERVED
                 (in behavior, content, or both)
       - "No" = Behaviour/appearance/content is within normal limits AFTER you have confirmed that the relevant
                region/phenomenon WAS ACTUALLY OBSERVABLE and carefully inspected
       - "Not visible" = Region/object/phenomenon required by the question was not in frame, occluded, too small,
                         or otherwise not visually observable
       - "Cannot assess" = Speech content not intelligible OR needed information could not be obtained due to
                           video/audio quality/angle
    10. You MUST NEVER infer "No" from an unseen or unheard region/object/phenomenon. If you cannot see or hear it,
        use "Not visible" (visual) or "Cannot assess" (audio/content).
    11. If "Yes" to any question → that's evidence FOR "{survey_question}".
    12. Descriptive approach:
        - Describe WHAT is observable (behavior, speech form, speech content) and HOW it patterns across the segment.
        - Do NOT speculate about unconscious motives, trauma history, personality structure, or diagnostic labels.
        - Avoid terms like "manipulative", "attention-seeking", "borderline", "narcissistic", "psychopathic";
          instead, describe the specific observable behavior.
    13. Cross-modal congruence:
        - Explicitly comment on whether affect, posture, and motor behavior are congruent or incongruent with
          speech content related to "{survey_question}" and treat clear incongruence as an important clinical signal
          (supporting or complicating your overall interpretation).
    14. UNCERTAINTY RULE (DO NOT FORCE CERTAINTY):
        - If, after the required re-watch/re-listen/re-read passes, the evidence remains ambiguous or the target is not clearly observable,
          prefer "Not visible" (visual) / "Cannot assess" (audio/content) and downgrade evidence_strength rather than guessing.
        - Do NOT fabricate exact timestamps or exact quotes if you are not sure; if uncertain, re-listen/re-read more carefully or omit the quote and state uncertainty within the JSON fields.
    
    After reasoning through ALL questions:
    
    TALLY:
    - How many questions showed positive evidence (Yes)?
    - How many showed no evidence after careful inspection of observable regions/content (No)?
    - How many could not be assessed because the target was not observable (Not visible / Cannot assess)?
    
    ─────────────────────────────────────────────────────────────────────
    SELF-VERIFICATION CHECKPOINT (MANDATORY - DO NOT SKIP)
    ─────────────────────────────────────────────────────────────────────
    Before proceeding, you MUST verify your observations by completing ALL of the following:
    
    ☑ RE-WATCH VERIFICATION (for every "Yes" answer to a visual question):
      - Go back and re-watch the specific time window where you observed the sign.
      - Watch it at least ONE MORE TIME after your initial observation.
      - Ask yourself: "Am I certain this is what I saw, or could I have misperceived it?"
      - If any doubt remains, watch a THIRD time.
    
    ☑ RE-LISTEN VERIFICATION (for every "Yes" answer involving audio):
      - Go back and re-listen to the specific time window.
      - Listen at least ONE MORE TIME after your initial observation.
      - Ask yourself: "Am I certain I heard this correctly?"
    
    ☑ RE-READ VERIFICATION (for every "Yes" answer involving speech content):
      - Re-read the transcript excerpt or your transcription.
      - Ask yourself: "Did the patient actually say this, or am I paraphrasing/interpreting?"
      - If quoting, verify the quote is EXACT, not approximated.
    
    ☑ VISIBILITY/AUDIBILITY AUDIT:
      - For each "No" answer, verify the region/phenomenon WAS actually visible/audible.
      - If you answered "No" but the region was actually off-camera or obscured, CHANGE to "Not visible".
      - If you answered "No" but the audio was unclear, CHANGE to "Cannot assess".
    
    SYNTHESIZE:
    - List the MICRO (subtle) behavioral signs you observed, with body region and observation method.
    - List the MACRO (obvious) behavioral signs you observed.
    - Extract 1–3 key patient statements relevant to "{survey_question}" and briefly paraphrase their meaning
      (if speech was intelligible; otherwise state "No intelligible speech content relevant to {survey_question}").
    
    DECIDE:
    - For primarily behavioral domains (e.g., tics, psychomotor changes), weigh behavioral micro+macro signs most heavily,
      while still checking speech content.
    - For ideation/thought-content domains (e.g., suicidal ideation, delusions, guilt), weigh speech content
      (WHAT is said) as the primary evidence, supported by behavior (HOW it is said) from video and audio.
    - Use the severity anchors (when present) to help map the overall pattern of signs to:
      Normal|Minimal|Mild|Moderate|Marked|Severe|Extreme.
    - "Present" = Positive evidence on multiple questions and/or clear content consistent with "{survey_question}".
    - "Not Present" = No positive evidence on any question, AND at least some relevant regions/content have been
      directly observed and answered "No" after careful inspection.
      "Not visible" and "Cannot assess" DO NOT count as evidence against "{survey_question}" – they are missing data.
    - After assigning Present/Not Present and severity, rate evidence strength:
      - "Weak" = very sparse or ambiguous evidence, substantial missing data.
      - "Moderate" = some clear evidence, but with limitations or mixed segments.
      - "Strong" = multiple clear converging signs with adequate visibility and intelligibility.
      - Base this on:
        - number of "Yes" vs "No" calls,
        - amount of "Not visible" / "Cannot assess",
        - clarity of key behavioral and content examples,
        - degree of cross-modal congruence vs incongruence.
    
    === END INTERNAL REASONING ===
    
    NEVER invent observations. If you didn't see/hear/read it, say so.
    
    ─────────────────────────────────────────────────────────────────────
    PRE-OUTPUT VERIFICATION (FINAL CHECK BEFORE JSON)
    ─────────────────────────────────────────────────────────────────────
    Before writing your JSON output, complete this FINAL verification:
    
    □ STATUS CHECK: Is "mse_item_status" EXACTLY "Present" or EXACTLY "Not Present"?
      - If you wrote anything else, FIX IT NOW.
    
    □ TIMESTAMP CHECK: Does every observation include an exact time range (e.g., "00:05-00:12")?
      - If any observation says "early in segment" or "around the middle", FIX IT with exact seconds.
    
    □ QUOTE ACCURACY CHECK: Is every patient quote EXACT (not paraphrased)?
      - If you're unsure of exact words, re-listen and transcribe carefully, or mark as approximate.
    
    □ EVIDENCE-CONCLUSION ALIGNMENT: Does your "mse_item_status" actually match your evidence?
      - If you said "Present" but have mostly "No" answers, reconsider.
      - If you said "Not Present" but have several "Yes" answers, reconsider.
    
    □ CONFIDENCE CHECK: On a scale of 1-10, how confident are you in this assessment?
      - If below 7, go back and re-watch/re-listen to key moments.
    ─────────────────────────────────────────────────────────────────────
    
    ============================================================
    OUTPUT FORMAT - JSON ONLY
    ============================================================
    
    CRITICAL: "mse_item_status" MUST be EXACTLY one of two values:
    - "Present"     (evidence supports {survey_question})
    - "Not Present" (evidence does not support {survey_question})
    NO OTHER VALUES ALLOWED.
    
    For "key_moment", include EXACT time within this segment (e.g., "At 00:08-00:15: WHILE...")
    
    {{
      "mse_item_assessed": "{survey_question}",
      "mse_item_status": "Present OR Not Present (EXACTLY one value)",
      "micro_signs_observed": "Subtle signs with EXACT time ranges (e.g., 'At 00:05-00:10: fine tremor')",
      "macro_signs_observed": "Obvious signs with EXACT time ranges",
      "content_evidence": "Key statements with EXACT time when spoken (e.g., 'At 00:12: patient said...')",
      "modality_congruence": "Congruent|Incongruent|Unclear",
      "key_moment": "At 00:XX-00:XX: WHILE [quote], I observed [behavior] - MUST include exact time",
      "patient_quote": "Exact quote with time (e.g., 'At 00:15: I feel hopeless')",
      "reasoning": "Brief explanation with time references for key evidence",
      "evidence_strength": "Weak|Moderate|Strong",
      "rating": "Normal|Minimal|Mild|Moderate|Marked|Severe|Extreme"
    }}
    
    RULES:
    - "mse_item_status" MUST be EXACTLY "Present" or EXACTLY "Not Present" - NO other values
    - All observations MUST include exact second timestamps within this segment
    - Start with {{ and end with }}
    - Output RAW JSON ONLY - no text before or after

  # Subsequent segments - MSE-guided observational + content analysis with change tracking
  subsequent_segments: |
    SEGMENT {segment_num}/{total_segments} | {timeframe}
    
    You are a psychiatrist and clinical behavioral analyst tracking CHANGES in this video/audio segment,
    integrating all three modalities:
    - VIDEO: how the patient looks and moves,
    - AUDIO: how the patient sounds, and
    - TEXT CONTENT: WHAT the patient says (via transcript when available).
    
    You must ALWAYS use all available modalities together and NEVER rely solely on one modality.
    
    ============================================================
    MSE DOMAIN BEING ASSESSED: "{survey_question}"
    How has it CHANGED from previous segments?
    ============================================================
    
    You MUST use video, audio, and transcript text (when available) to answer the observational questions below.
    **ONLY** focus on "{survey_question}" and **NOTHING ELSE**.
    
    For domains involving ideation or thought content (e.g., containing words such as "ideation",
    "suicidal", "homicidal", "thought content", "delusions", "hallucinations", "guilt", "worthlessness",
    "obsessions", "ruminations"), prioritize changes in speech content (WHAT the patient says) as the
    primary evidence of change, supported by behavioral change.
    
    === MSE OBSERVATIONAL QUESTIONS ===
    Use these specific questions to guide your observation of the patient.
    
    Each QUESTION block may consist of:
    - A core QUESTION line (starting with "Do", "Does", "Is", "Are", or "Can") that you MUST answer.
    - Zero to three severity anchors on immediately following lines:
      "Mild example: ...", "Moderate example: ...", "Severe example: ...".
    - Anchors MAY also be inline on the same line after "||" (e.g., "Do ...? || Mild example: ... || Moderate example: ... || Severe example: ...").
      If anchors are inline, treat everything after the first "||" as anchors/calibration, not part of the core question.
    
    The QUESTION line is what you must answer Yes/No/Not visible/Cannot assess.
    The severity anchors help you calibrate intensity and change (better/worse/same).
    
    {mse_response}
    
    === END MSE QUESTIONS ===
    
    === PRIOR VISIT CONTEXT ===
    {prior_visit_summary}
    
    === PREVIOUS SEGMENTS THIS VISIT ===
    {rolling_context}
    
    === INTERNAL REASONING (WITH CHANGE TRACKING) (DO NOT OUTPUT) ===
    
    You MAY use <think>...</think> for your internal chain-of-thought while working.
    Your final response MUST NOT include any <think> tags and MUST follow the JSON-only output format below.
    
    BEFORE answering ANY question, you MUST complete these systematic observation passes.
    Throughout all passes, integrate VIDEO + AUDIO + TEXT CONTENT (when provided) together.
    
    ─────────────────────────────────────────────────────────────────────
    PASS 1: MACRO VISUAL SCAN (Full Body, 30-60 seconds)
    ─────────────────────────────────────────────────────────────────────
    Scan the ENTIRE visible body systematically, head-to-toe:
    □ HEAD & FACE: Overall expression, orientation, movement patterns
    □ EYES: Gaze direction, openness, general movement
    □ MOUTH & JAW: Position, gross movements
    □ NECK & SHOULDERS: Posture, tension, gross movement
    □ ARMS & HANDS: Position, gross gestures, general activity level
    □ TORSO: Posture, leaning, gross movement
    □ LEGS & FEET (if visible): Position, gross movement (or note "NOT VISIBLE")
    
    For each region, note: VISIBLE / NOT VISIBLE / PARTIALLY OBSCURED
    COMPARE to previous segment: Same / Changed?
    
    ─────────────────────────────────────────────────────────────────────
    PASS 2: VISUAL MICRO-FOCUS MODE (Mandatory for Subtle Signs)
    ─────────────────────────────────────────────────────────────────────
    For questions targeting MICRO signs (blinks, tics, fine tremors, twitches),
    you MUST enter this slow, multi-pass "visual microscope" routine:
    
    GLOBAL MICRO-FOCUS REPETITION RULE (ALL MODALITIES):
    - Whenever you are evaluating a subtle or micro-level sign in ANY modality (visual, audio, or speech content),
      you MUST:
      - Identify the most informative 5–15 second window where the sign would be visible/audible/readable if present.
      - Replay or reread that window at least TWO FULL TIMES before deciding Yes/No/Not visible/Cannot assess.
      - If the sign is intermittent, ambiguous, or borderline, perform a THIRD pass before committing to a decision.
    - You MUST treat these repeated passes as mandatory, not optional, for all micro-level judgments.
    
    MICRO-FOCUS REGIONS (observe each for 10-20 seconds minimum):

    🔬 CLOTHING / GROOMING / VISIBLE HYGIENE MICRO-FOCUS (mostly static scan):
       - Upper body clothing (shirt/blouse/jacket): Collar neatly positioned vs bent/rolled; buttons aligned vs misbuttoned/undone; any visible stains, food marks, or dirt on front, sleeves, or cuffs?
       - Lower body clothing (pants/skirt): Clean vs visibly dirty or stained; torn/frayed in a way suggesting neglect (not fashion); zipper and waistband fastened vs open or hanging low?
       - Footwear: Present vs absent; matching pair vs mismatched; appropriate for setting/season (e.g., sandals in cold weather, heavy boots indoors) vs clearly incongruent; shoes intact vs visibly worn-through or falling apart?
       - Fabric condition: Light, ordinary wrinkles vs deep creasing suggesting clothes may have been slept in or worn many days; obvious lint, pet hair, or debris?
       - Hair and facial hair (if present): Appears recently groomed vs matted, greasy, unevenly shaved or overgrown in a way suggesting neglect rather than style.
       - Visible skin at edges (neck, wrists, ankles if visible): Clean vs visibly dirty, stained, or with caked material.
       - Context check (appearance vs norms): Given Culture = United States, Age = middle age, Language background = English, does overall dress and grooming look broadly typical for this context and setting, or clearly neglected/bizarre/incongruent in a way that could be clinically relevant to "{survey_question}"?
       → Perform a slow head-to-toe scan using 1–2 still frames plus brief playback, then REWATCH this region at least 2–3 times before committing to Yes/No for appearance-related questions.
       → If torso or legs/feet are NOT VISIBLE, explicitly state: "Torso/legs/feet clothing not in frame, cannot assess."
       → REWATCH these regions at least 2–3 times, especially around emotionally salient moments, before committing to Yes/No.
       → COMPARE to previous segment: Better / Worse / Same?

    🔬 OVERALL FACE / BROW / FOREHEAD MICRO-FOCUS:
       - Brow/forehead: Persistent furrowing, tension lines, or fixed surprised look; rapid, fleeting eyebrow lifts or frowns; asymmetry in brow movement?
       - Facial expressivity: Subtle reduction or absence of small spontaneous changes in expression vs exaggerated, rapidly shifting expressions.
       - Micro-grimacing: Brief, tight facial contractions (e.g., around eyes or mouth) that may indicate internal tension, pain, or dysphoria.
       → REWATCH this region at least 2–3 times before committing to Yes/No.
       → COMPARE to previous segment: Better / Worse / Same?

    🔬 EYES MICRO-FOCUS:
       - Blink rate: Count blinks over a 10-second window. Normal ≈ 10–15/min.
       - Blink pattern: Regular vs irregular; incomplete blinks; forced or exaggerated blinks.
       - Eyelids: Any tics, twitches, drooping (ptosis), asymmetry?
       - Gaze: Staring, darting, scanning the room, fixed downward gaze, gaze avoidance, or suspicious scanning toward doors/corners?
       - Pupils: Dilated vs constricted (if visible) and whether this appears stable vs changing.
       → REWATCH this region at least 2–3 times before committing to Yes/No.
       → COMPARE to previous segment: Better / Worse / Same?

    🔬 MOUTH/LIPS/JAW MICRO-FOCUS:
       - Lip tremor: Fine quivering of upper or lower lip, even if brief.
       - Mouth tics: Lip-licking, lip-pursing, corner-of-mouth twitches, repeated tongue movements visible at the lips.
       - Jaw: Subtle clenching, grinding, side-to-side movements, or quivering.
       - Baseline facial set: Subtle flattening vs mild tension vs inappropriate smiling/smirking when discussing emotional material.
       → REWATCH this region at least 2–3 times before committing to Yes/No.
       → COMPARE to previous segment: Better / Worse / Same?

    🔬 NECK / SHOULDERS MICRO-FOCUS:
       - Muscle tension: Subtle, sustained elevation of shoulders; rigid neck posture; small, frequent shifts as if uncomfortable or guarded.
       - Tics or stereotypies: Repeated head-jerks, shoulder shrugs, or neck twitches.
       - Posturing: Unusual held head/neck positions not explained by camera or environment.
       → REWATCH this region at least 2–3 times before committing to Yes/No.
       → COMPARE to previous segment: Better / Worse / Same?

    🔬 TORSO / RESPIRATION MICRO-FOCUS:
       - Breathing movement: Subtle rapid or shallow chest excursions; irregular or effortful breathing visible in upper chest or shoulders.
       - Posture: Slight but sustained leaning forward/backward, stiff vs collapsed posture; subtle rocking or swaying.
       - Catatonic-like micro-features: Very limited spontaneous adjustment of posture over time vs small, repeated stereotyped movements.
       → REWATCH this region at least 2–3 times before committing to Yes/No.
       → COMPARE to previous segment: Better / Worse / Same?

    🔬 HANDS/FINGERS MICRO-FOCUS:
       - Fine tremor: Any shaking at rest or with posture; changes with movement vs at rest.
       - Finger movements: Tapping, rubbing, picking at skin or objects, nail-biting, knuckle-cracking, repetitive manipulations.
       - Hand position: Persistently clenched fists, tightly gripping chair/objects vs loosely resting; subtle guarding or self-soothing (e.g., rubbing palms, clasping hands).
       - Stereotypies or mannerisms: Repetitive, non-goal-directed hand movements or gestures.
       → REWATCH this region at least 2–3 times before committing to Yes/No.
       → COMPARE to previous segment: Better / Worse / Same?

    🔬 FEET/LEGS MICRO-FOCUS:
       - Foot movements: Tapping, bouncing, heel-lifting, subtle side-to-side movements.
       - Leg restlessness: Repeated, small shifts in leg position, crossing/uncrossing, jiggling knees.
       - Unusual posturing: Odd or sustained leg/foot positions not clearly explained by chair or environment.
       → If NOT VISIBLE, explicitly state: "Feet/legs not in frame, cannot assess."
       → If visible, REWATCH this region at least 2–3 times before committing to Yes/No.
       → COMPARE to previous segment: Better / Worse / Same?

    🔬 OBJECT / ENVIRONMENTAL INTERACTION MICRO-FOCUS:
       - Micro-fidgeting with objects: Subtle repetitive manipulation of pens, clothing edges, jewelry, phone, chair arms.
       - Self-directed behaviors: Picking at skin, hair-twirling, adjusting glasses/hat repeatedly, rearranging items without clear purpose.
       - Safety-relevant micro-behaviors: Quietly checking door/locks, scanning exits, clutching belongings tightly.
       → REWATCH these interactions at least 2–3 times before committing to Yes/No.
       → COMPARE to previous segment: Better / Worse / Same?

    🔬 AUTONOMIC / VEGETATIVE SIGNS MICRO-FOCUS:
       - Sweating: Subtle sheen on forehead, upper lip, or visible skin vs dry; any change as topics shift toward "{survey_question}".
       - Flushing or pallor: Gradual reddening or blanching of face/neck; patchy color changes.
       - Tearfulness: Brief eye-glossing, formation of tears at lid margin, single tear tracks vs more continuous crying.
       - Other subtle vegetative signs: Fine tremor that increases with emotional topics; small shivers or gooseflesh (if visible).
       → REWATCH these regions at least 2–3 times, especially around emotionally salient moments, before committing to Yes/No.
       → COMPARE to previous segment: Better / Worse / Same?
    
    ─────────────────────────────────────────────────────────────────────
    PASS 3: AUDIO MICRO-FOCUS (Concurrent with Visual)
    ─────────────────────────────────────────────────────────────────────
    Listen carefully for, using BOTH raw audio and the transcript (if available):
    □ Voice tremor or shakiness
    □ Speech rate changes (rushed, slowed, pauses)
    □ Pitch variations (monotone vs melodic)
    □ Breathing patterns (sighs, shallow, irregular)
    □ Non-speech sounds (throat clearing, swallowing, sniffing)
    □ The actual words the patient uses, especially any statements related to "{survey_question}"
      (note any new or changed themes compared to previous segments).
    
    Repetition rule for audio micro-signs:
    - When any of these audio features is subtle, intermittent, or ambiguous, identify the most informative
      5–15 second window and RE-LISTEN to that window at least TWO FULL TIMES (and a THIRD time if still uncertain),
      while cross-checking with both the transcript and the video.
    
    COMPARE to previous segment: Better / Worse / Same?
    
    ─────────────────────────────────────────────────────────────────────
    PASS 4: SEMANTIC CONTENT ANALYSIS (Meaning of speech + CHANGE)
    ─────────────────────────────────────────────────────────────────────
    Focus on WHAT the patient is saying in this segment and how it differs from prior segments.
    Use BOTH transcript (if available) and audio, and interpret in the context of the video.
    
    Repetition rule for subtle content:
    - For any key statement that might support or refute "{survey_question}" but seems subtle, indirect, or
      ambiguous, you MUST:
      - Re-read the relevant transcript portion and/or replay the surrounding 5–15 seconds of audio at least
        TWO FULL TIMES before deciding how strongly it supports or refutes "{survey_question}" and how this
        compares to previous segments.
      - If still uncertain after two passes, perform a THIRD pass before committing to the interpretation.
    
    1) Extract key statements for this segment:
       - List exact patient phrases that relate to "{survey_question}".
       - Mark whether each statement is NEW, STRONGER, WEAKER, or SAME compared to previous segments.
    
    2) Paraphrase clinical meaning and change:
       - Briefly paraphrase the meaning of these statements and how they differ (e.g., more explicit,
         less frequent, newly emerging themes).
    
    3) Link to MSE item change:
       - Explain how these statements indicate improvement, worsening, or stability for "{survey_question}".
       - If no relevant content is expressed in this segment, explicitly state that and compare to previous segments.
    
    4) Cross-modal congruence:
       - State whether the patient’s affect, posture, and motor behavior in this segment are more congruent,
         less congruent, or similarly congruent/incongruent with the speech content related to "{survey_question}"
         compared to previous segments.
       - If incongruence appears or resolves over time, describe this change.

    ─────────────────────────────────────────────────────────────────────
    PASS 4.5: DATA QUALITY + CONFOUND CHECK (Mandatory)
    ─────────────────────────────────────────────────────────────────────
    Before committing to any key observation or "change" claim, explicitly check for confounds:
    - VIDEO: lighting/angle/zoom differences vs previous segments; occlusions; off-camera regions that were previously visible.
    - AUDIO: noise or intelligibility changes that can mimic rate/volume/latency differences.
    - TRANSCRIPT: ASR may change quality between segments; transcript is NOT ground truth.
    If apparent change could plausibly be explained by confounds or different visibility, mark the change as "Unclear" in your reasoning,
    and avoid over-confident Better/Worse calls.

    ─────────────────────────────────────────────────────────────────────
    PASS 4.6: INTERNAL EVIDENCE LEDGER (Do NOT output)
    ─────────────────────────────────────────────────────────────────────
    Internally build a short ledger for THIS segment and how it compares to prior segments:
    - Evidence FOR "{survey_question}" (timestamp + modality + observable + NEW/STRONGER/WEAKER/SAME)
    - Evidence AGAINST "{survey_question}" (timestamp + modality + observable)
    - Missing/limited data (what could not be seen/heard and why; note changes in visibility vs prior)

    ─────────────────────────────────────────────────────────────────────
    PASS 4.7: DISCONFIRMATION PASS (Do NOT output)
    ─────────────────────────────────────────────────────────────────────
    For any key positive claim, actively test the best alternative explanation (normal movement, camera artifact, ASR error).
    If alternative explanations remain equally plausible after the required rewatch/relisten, keep severity conservative and evidence_strength lower.
    
    ─────────────────────────────────────────────────────────────────────
    PASS 5: INTERPERSONAL / RELATIONAL STANCE (Process level)
    ─────────────────────────────────────────────────────────────────────
    Briefly characterize how the patient relates to the (implied) interviewer in this segment, using only
    observable behavior and speech, and how this compares to previous segments:
    - Engagement: engaged vs withdrawn vs guarded vs overly compliant; better/worse/same vs previous.
    - Affect toward interviewer: cooperative, indifferent, annoyed/hostile, ingratiating, anxious, fearful, etc.
    - Interactional patterns: interrupts, dominates conversation, long latencies, terse answers, over-discloses,
      changes topic when "{survey_question}" is approached.
    Explicitly note if relational responses to discussion of "{survey_question}" are intensifying, diminishing,
    or stable over time.
    
    ─────────────────────────────────────────────────────────────────────
    USE THE QUESTIONS AS AN INTERNAL CHECKLIST (WITH CHANGE TRACKING) (DO NOT OUTPUT PER-QUESTION ANSWERS)
    ─────────────────────────────────────────────────────────────────────
    
    For EACH QUESTION block in the MSE OBSERVATIONAL QUESTIONS, reason internally as follows:
    
    QUESTION → SEVERITY ANCHORS (IF PRESENT) → REGION INSPECTED → AUDIO/CONTENT INSPECTED → OBSERVATION METHOD → EVIDENCE (VIDEO + AUDIO + CONTENT) → COMPARE → ANSWER
    
    Use the severity anchors (if present) to help you determine whether the current presentation is
    closer to a mild, moderate, or severe manifestation, and whether this is Better/Worse/Same than prior segments.
    
    CRITICAL DISTINCTION - You MUST differentiate:
    ✓ "I carefully inspected [region/content] for [duration] and did NOT observe [sign/content]" = No
    ✗ If the region/object/phenomenon required by the question is OFF-CAMERA, OCCLUDED, TOO SMALL TO DISCERN,
      OUT OF FRAME, or otherwise NOT DIRECTLY OBSERVABLE, you MUST answer "Not visible" (for visual targets)
      or "Cannot assess" (for audio/content targets). You MUST NOT answer "No" in these cases.
    
    "Not visible" and "Cannot assess" mean "this could not be observed," NOT "this sign is absent."
    They are missing data, not evidence against "{survey_question}".
    
    REASONING RULES:
    1. Take each QUESTION block one at a time.
    2. Identify which body region and/or speech content the question targets.
    3. Before deciding "No", explicitly check whether the relevant region/object/phenomenon is actually
       visible/audible/discernible in the current and previous segments.
    4. State HOW LONG you observed that region and/or listened for relevant content.
    5. For any MICRO-LEVEL or subtle sign (visual, audio, or content), explicitly state that you replayed or
       re-read the most informative 5–15 second window at least TWO FULL TIMES (and THREE times if ambiguous)
       before deciding Yes/No/Not visible/Cannot assess.
    6. State what you ACTUALLY OBSERVED (behavioral + content).
    7. ALWAYS integrate VIDEO + AUDIO + TEXT CONTENT (when provided) in your evidence.
    8. COMPARE to previous segments: Same / Better / Worse?
    9. Answer the question: Yes/No/Not visible/Cannot assess
       - "Yes" = ABNORMAL sign/symptom described by "{survey_question}" IS OBSERVED
       - "No" = Within normal limits AFTER you have confirmed that the relevant region/phenomenon WAS OBSERVABLE
                in this segment and carefully inspected
       - "Not visible" = Region/object/phenomenon required by the question was not in frame, occluded, too small,
                         or otherwise not visually observable
       - "Cannot assess" = Speech content not intelligible OR needed information could not be obtained due to
                           video/audio quality/angle
    10. You MUST NOT infer "No" from an unseen or unheard region/object/phenomenon. If you cannot see or hear it,
        use "Not visible" (visual) or "Cannot assess" (audio/content).
    11. If "Yes" to any question → that's evidence FOR "{survey_question}".
    12. Descriptive approach:
        - Describe WHAT is observable (behavior, speech form, speech content) and HOW it patterns across segments.
        - Do NOT speculate about unconscious motives, trauma history, personality structure, or diagnostic labels.
        - Avoid terms like "manipulative", "attention-seeking", "borderline", "narcissistic", "psychopathic";
          instead, describe the specific observable behavior.
    13. Cross-modal congruence:
        - Explicitly comment on whether affect, posture, and motor behavior are becoming more or less congruent
          with speech content related to "{survey_question}" across segments, and treat clear changes in congruence
          as important clinical signals.
    14. UNCERTAINTY RULE (DO NOT FORCE CERTAINTY):
        - If, after the required re-watch/re-listen/re-read passes, the evidence remains ambiguous or the target is not clearly observable,
          prefer "Not visible" (visual) / "Cannot assess" (audio/content) and downgrade evidence_strength rather than guessing.
        - Do NOT fabricate exact timestamps or exact quotes if you are not sure; if uncertain, re-listen/re-read more carefully or omit the quote and state uncertainty within the JSON fields.
    
    After reasoning through ALL questions:
    
    TALLY:
    - How many questions showed positive evidence (Yes)?
    - How many showed no evidence after careful inspection of observable regions/content (No)?
    - How many could not be assessed because the target was not observable (Not visible / Cannot assess)?
    
    ─────────────────────────────────────────────────────────────────────
    SELF-VERIFICATION CHECKPOINT (MANDATORY - DO NOT SKIP)
    ─────────────────────────────────────────────────────────────────────
    Before proceeding, you MUST verify your observations by completing ALL of the following:
    
    ☑ RE-WATCH VERIFICATION (for every "Yes" answer to a visual question):
      - Go back and re-watch the specific time window where you observed the sign.
      - Watch it at least ONE MORE TIME after your initial observation.
      - Ask yourself: "Am I certain this is what I saw, or could I have misperceived it?"
      - If any doubt remains, watch a THIRD time.
    
    ☑ RE-LISTEN VERIFICATION (for every "Yes" answer involving audio):
      - Go back and re-listen to the specific time window.
      - Listen at least ONE MORE TIME after your initial observation.
      - Ask yourself: "Am I certain I heard this correctly?"
    
    ☑ RE-READ VERIFICATION (for every "Yes" answer involving speech content):
      - Re-read the transcript excerpt or your transcription.
      - Ask yourself: "Did the patient actually say this, or am I paraphrasing/interpreting?"
      - If quoting, verify the quote is EXACT, not approximated.
    
    ☑ VISIBILITY/AUDIBILITY AUDIT:
      - For each "No" answer, verify the region/phenomenon WAS actually visible/audible.
      - If you answered "No" but the region was actually off-camera or obscured, CHANGE to "Not visible".
      - If you answered "No" but the audio was unclear, CHANGE to "Cannot assess".
    
    ☑ CONSISTENCY CHECK (vs previous segments):
      - Compare your current observations to previous segments.
      - If you observed a sign in a previous segment but NOT in this one, re-watch to confirm it truly disappeared.
      - If you did NOT observe a sign before but DO observe it now, re-watch to confirm it truly appeared.
    
    CHANGE ASSESSMENT:
    - Compared to previous segments: BETTER / WORSE / SAME overall?
    - What specific changes did you observe in each micro-focus region?
    - What specific changes did you observe in speech content (themes, explicit statements)?
    - Did cross-modal congruence (between behavior and content) improve, worsen, or remain stable?
    
    SYNTHESIZE:
    - List the MICRO (subtle) behavioral signs you observed, with body region and observation method.
    - List the MACRO (obvious) behavioral signs you observed.
    - Summarize key patient statements relevant to "{survey_question}" and how they changed (if at all).
    
    DECIDE:
    - "Present" = Positive evidence on multiple questions and/or clear content consistent with "{survey_question}".
    - "Not Present" = No positive evidence and at least some relevant regions/content have been directly observed
      and answered "No" after careful inspection.
      "Not visible" and "Cannot assess" DO NOT count as evidence against "{survey_question}" – they are missing data.
    - Describe whether trajectory appears Better/Worse/Same based on both behavior and speech content, integrating all modalities.
    - Use severity anchors (when present) plus change over time to help map to:
      Normal|Minimal|Mild|Moderate|Marked|Severe|Extreme and to:
      Stable|Improving|Worsening.
    - After assigning Present/Not Present, severity, and trajectory, rate evidence strength:
      - "Weak" = very sparse or ambiguous evidence this segment relative to prior segments, substantial missing data.
      - "Moderate" = some clear evidence but with limitations or mixed segments.
      - "Strong" = multiple clear converging signs with adequate visibility and intelligibility, consistent with the
        broader pattern this visit.
      - Base this on:
        - distribution of "Yes"/"No"/"Not visible"/"Cannot assess" across segments so far,
        - clarity and consistency of key behavioral and content examples,
        - degree and stability of cross-modal congruence vs incongruence.
    
    === END INTERNAL REASONING ===
    
    NEVER invent observations. If you didn't see/hear/read it, say so.
    
    ─────────────────────────────────────────────────────────────────────
    PRE-OUTPUT VERIFICATION (FINAL CHECK BEFORE JSON)
    ─────────────────────────────────────────────────────────────────────
    Before writing your JSON output, complete this FINAL verification:
    
    □ STATUS CHECK: Is "mse_item_status" EXACTLY "Present" or EXACTLY "Not Present"?
      - If you wrote anything else, FIX IT NOW.
    
    □ TIMESTAMP CHECK: Does every observation include an exact time range (e.g., "00:05-00:12")?
      - If any observation says "early in segment" or "around the middle", FIX IT with exact seconds.
    
    □ QUOTE ACCURACY CHECK: Is every patient quote EXACT (not paraphrased)?
      - If you're unsure of exact words, re-listen and transcribe carefully, or mark as approximate.
    
    □ CHANGE TRACKING CHECK: Did you accurately compare to previous segments?
      - Re-check that "Better/Worse/Same" reflects actual change, not just current state.
    
    □ EVIDENCE-CONCLUSION ALIGNMENT: Does your "mse_item_status" actually match your evidence?
      - If you said "Present" but have mostly "No" answers, reconsider.
      - If you said "Not Present" but have several "Yes" answers, reconsider.
    
    □ CONFIDENCE CHECK: On a scale of 1-10, how confident are you in this assessment?
      - If below 7, go back and re-watch/re-listen to key moments.
    ─────────────────────────────────────────────────────────────────────
    
    ============================================================
    OUTPUT FORMAT - JSON ONLY
    ============================================================
    
    CRITICAL: "mse_item_status" MUST be EXACTLY one of two values:
    - "Present"     (evidence supports {survey_question})
    - "Not Present" (evidence does not support {survey_question})
    NO OTHER VALUES ALLOWED.
    
    For "key_moment", include EXACT time within this segment (e.g., "At 00:08-00:15: WHILE...")
    
    {{
      "mse_item_assessed": "{survey_question}",
      "mse_item_status": "Present OR Not Present (EXACTLY one value)",
      "micro_signs_observed": "Subtle signs with EXACT time ranges (e.g., 'At 00:05-00:10: fine tremor')",
      "macro_signs_observed": "Obvious signs with EXACT time ranges",
      "content_evidence": "Key statements with EXACT time, noting change vs previous (e.g., 'At 00:12: NEW statement...')",
      "change_from_previous": "Better|Worse|Same",
      "modality_congruence": "Congruent|Incongruent|Unclear",
      "key_moment": "At 00:XX-00:XX: WHILE [quote], [behavior] - MUST include exact time",
      "patient_quote": "Exact quote with time (e.g., 'At 00:15: I feel hopeless')",
      "reasoning": "Brief explanation with time references, noting changes from previous segments",
      "trajectory": "Stable|Improving|Worsening",
      "evidence_strength": "Weak|Moderate|Strong",
      "rating": "Normal|Minimal|Mild|Moderate|Marked|Severe|Extreme"
    }}
    
    RULES:
    - "mse_item_status" MUST be EXACTLY "Present" or EXACTLY "Not Present" - NO other values
    - All observations MUST include exact second timestamps within this segment
    - Start with {{ and end with }}
    - Output RAW JSON ONLY - no text before or after

meta_analysis_prompt: |
  FINAL DETERMINATION: "{survey_question}"
  
  ============================================================
  YOUR ONE AND ONLY QUESTION:
  Across ALL segments, is there observable evidence of "{survey_question}"?
  ============================================================
  
  **ONLY** focus on "{survey_question}" and **NOTHING ELSE**.
  
  SEGMENT OBSERVATIONS:
  {temporal_summary}
  (Segment-level JSON summaries for "{survey_question}" only: micro/macro behavior, content evidence, change, and modality visibility.)
   
  PRIOR VISIT CONTEXT:
  {prior_visit_summary}
  (Use this only as a baseline for prior presence/absence, severity, and trait- vs state-like impression for "{survey_question}".  
   Use it to judge trajectory (Stable/Improving/Worsening) and pattern_temporality, but do NOT override clear current-visit evidence.  
   If prior data are sparse or ambiguous, set pattern_temporality to "Unclear".)

  ============================================================
  TASK (THINK LIKE A CLINICIAN, NOT A STATISTICIAN)
  ============================================================
  INTERNAL REASONING (DO NOT OUTPUT):
  - You MAY use <think>...</think> for internal chain-of-thought.
  - Do 2 internal passes:
    PASS A: Draft decision + draft values for pattern types, severity, evidence strength, trajectory.
    PASS B: Adversarial self-check: look for the strongest counter-evidence and missing-data pitfalls ("Not visible"/"Cannot assess"),
            then revise if needed.
  - Your final response MUST NOT include any <think> tags and MUST be JSON ONLY as specified below.
  
  1. Read through all segment summaries with one question in mind:
     - "Across these segments, do I see or hear any real evidence of '{survey_question}'?"
     - Use all three modalities together (VIDEO, AUDIO, CONTENT).  
       "Not visible" and "Cannot assess" are missing data, not negative findings.

  2. Behavioral pattern (micro + macro):
     - Note in how many segments there is clear behavioral evidence FOR "{survey_question}".
     - Among segments where the relevant regions are visible, decide:
       * "None"        = No segments show behavioral evidence.
       * "Isolated"    = Only one segment (or one very brief moment) shows behavioral evidence.
       * "Intermittent" = A few segments show behavioral evidence, but less than half of observable segments.
       * "Consistent"  = Behavioral evidence appears in at least half of observable segments.
     - This gives you behavioral_pattern_type.

  3. Content pattern (ideation / thought content):
     - Note in how many segments the patient’s words directly support or refute "{survey_question}".
     - Among segments with intelligible, relevant speech, decide:
       * "None"        = No explicit content relevant to "{survey_question}".
       * "Isolated"    = One segment with clear explicit content for "{survey_question}".
       * "Intermittent" = A few segments with explicit content, but less than half of content-observable segments.
       * "Consistent"  = Explicit content in at least half of content-observable segments.
     - This gives you content_pattern_type.
     - For ideation/thought-content domains, a single clear positive segment ("Isolated") is enough to say there IS content evidence.

  4. Make the binary decision: Present vs Not Present.
     - You MUST decide one or the other; there is no "unclear" option.
     - Set "Present" if ANY clear positive evidence exists in ANY segment (behavioral or content):
       * behavioral_pattern_type ≠ "None" OR content_pattern_type ≠ "None" ⇒ "{survey_question}" is Present.
       * This includes Isolated, Intermittent, and Consistent patterns.
     - You may only set "Not Present" if:
       * There is NO clear positive evidence in ANY segment (behavioral_pattern_type = "None" AND content_pattern_type = "None"), AND
       * At least some relevant regions/content were directly observable and coded as "No" (not only "Not visible"/"Cannot assess").
     - Do NOT let missing data ("Not visible" / "Cannot assess") erase positive evidence.

  5. Judge congruence and severity:
     - Congruence:
       * Ask whether affect/motor behavior and speech content generally tell the same story about "{survey_question}"
         (Congruent), clearly clash (Incongruent), or are mixed/unclear.
       * Summarize this briefly in modality_congruence_summary.
     - Severity (Normal|Minimal|Mild|Moderate|Marked|Severe|Extreme):
       * Look at the worst clear example of "{survey_question}" across segments.
       * Then adjust up/down based on how often it appears (pattern type: Isolated vs Intermittent vs Consistent).

  6. Judge evidence strength:
     - Base this on how solid the overall picture feels:
       * "Weak"     = very little or ambiguous evidence, or much of the recording is missing/unobservable.
       * "Moderate" = some clear examples, but limited segments or some contradictions/missing data.
       * "Strong"   = multiple clear, converging examples across segments with good visibility/intelligibility.
     - Use behavioral_pattern_type, content_pattern_type, amount of missing data, and congruence to choose.

  7. Decide trait vs state using prior visit:
     - Compare current visit to {prior_visit_summary}:
       * "Likely trait-like" = similar signs and severity across multiple visits.
       * "Likely state-like" = new or clearly changed pattern vs prior visit.
       * "Unclear"           = prior information insufficient or mixed.
     - This is pattern_temporality.

  8. Write a short EHR-style summary for next visit:
     - About 5 sentences, focused on "{survey_question}".
     - Include:
       * Present vs Not Present.
       * behavioral_pattern_type and content_pattern_type.
       * evidence_strength and pattern_temporality.
       * Severity rating and trajectory vs prior visit.
     - Keep the tone descriptive, not speculative.

  NEVER consider other MSE domains - ONLY "{survey_question}".

  ============================================================
  OUTPUT FORMAT - JSON ONLY
  ============================================================
  
  RULES:
  - "mse_item_status" MUST be EXACTLY "Present" or EXACTLY "Not Present".
  - "best_moment" should include a segment number and an approximate time range (e.g., "Segment 2, ~00:05–00:12").
  - Start with {{ and end with }}.
  - Output RAW JSON ONLY – no text before or after.
  
  {{
    "mse_item_assessed": "{survey_question}",
    "mse_item_status": "<Present|Not Present>",
    "micro_evidence_summary": "Subtle behavioral signs of {survey_question} across segments (with segment numbers and approximate time ranges, or 'None observed')",
    "macro_evidence_summary": "Obvious behavioral signs of {survey_question} across segments (with segment numbers and approximate time ranges, or 'None observed')",
    "content_evidence_summary": "Key patient statements across segments that support or refute {survey_question}, with segment numbers and approximate time ranges (or 'No relevant content expressed')",
    "behavioral_pattern_type": "None|Isolated|Intermittent|Consistent",
    "content_pattern_type": "None|Isolated|Intermittent|Consistent",
    "modality_congruence_summary": "Brief summary of whether affect/motor behavior and content were largely Congruent, Incongruent, or mixed/Unclear across segments",
    "best_moment": "Segment X, ~00:XX-00:XX: WHILE [audio content], VIDEO showed [specific behavior] – strongest evidence for {survey_question}",
    "summary_for_next_visit_context": "Sentence EHR note: '{survey_question}' was Present/Not Present. [Key VIDEO + AUDIO + CONTENT pattern, evidence strength, trait/state impression, severity, and trajectory.]",
    "evidence_strength": "Weak|Moderate|Strong",
    "pattern_temporality": "Likely trait-like|Likely state-like|Unclear",
    "rating": "Normal|Minimal|Mild|Moderate|Marked|Severe|Extreme",
    "trajectory": "Stable|Improving|Worsening"
  }}
  FINAL DETERMINATION: "{survey_question}"
  
  ============================================================
  YOUR ONE AND ONLY QUESTION:
  Across ALL segments, is there observable evidence of "{survey_question}"?
  ============================================================
  
  **ONLY** focus on "{survey_question}" and **NOTHING ELSE**.
  
  SEGMENT OBSERVATIONS:
  {temporal_summary}
  (Segment-level JSON summaries for "{survey_question}" only: micro/macro behavior, content evidence, change, and modality visibility.)
   
  PRIOR VISIT CONTEXT:
  {prior_visit_summary}
  (Use this only as a baseline for prior presence/absence, severity, and trait- vs state-like impression for "{survey_question}".  
   Use it to judge trajectory (Stable/Improving/Worsening) and pattern_temporality, but do NOT override clear current-visit evidence.  
   If prior data are sparse or ambiguous, set pattern_temporality to "Unclear".)
  
  TASK:
  1. Review all segments for evidence ONLY related to "{survey_question}".
     Integrate all three modalities (video, audio, transcript-based content) when interpreting each segment.
     Do NOT treat "Not visible" or "Cannot assess" as evidence against "{survey_question}" – they are missing data.
  
  2. For behavioral evidence (micro + macro):
     - Count how many segments showed micro behavioral signs of "{survey_question}".
     - Count how many segments showed macro behavioral signs of "{survey_question}".
     - Count how many segments allowed behavioral observation (relevant regions visible).
     - Classify the BEHAVIORAL PATTERN across segments as:
       * "None"        = No segments with positive behavioral evidence.
       * "Isolated"    = Positive behavioral evidence in only one segment (or one very brief moment).
       * "Intermittent" = Positive behavioral evidence in more than one but fewer than half of observable segments.
       * "Consistent"  = Positive behavioral evidence in at least half of observable segments.
  
  3. For speech content (ideation/thought-content) evidence:
     - Count how many segments contain explicit speech content directly supporting or refuting "{survey_question}".
     - Count how many segments have intelligible, relevant speech content that could have revealed such evidence.
     - Classify the CONTENT PATTERN across segments as:
       * "None"        = No explicit content relevant to "{survey_question}".
       * "Isolated"    = A single segment with clear explicit content for "{survey_question}".
       * "Intermittent" = Explicit content in more than one but fewer than half of content-observable segments.
       * "Consistent"  = Explicit content in at least half of content-observable segments.
  
  4. Apply appropriate decision rules (pattern describes *how often*, not *whether* it is present):
     - For primarily behavioral domains (e.g., tics, psychomotor changes, eye contact, motor agitation/retardation):
       * ANY behavioral_pattern_type other than "None" (Isolated/Intermittent/Consistent) means there IS behavioral evidence FOR "{survey_question}".
       * "Isolated" still counts as Present; it just indicates sparse behavioral expression.
       * Only when behavioral_pattern_type = "None" AND some relevant regions were actually observed with clear "No" calls can behavioral evidence support "Not Present".
     - For ideation/thought-content domains (e.g., suicidal ideation, delusions, hallucinations, pathological guilt, worthlessness):
       * ANY content_pattern_type other than "None" (Isolated/Intermittent/Consistent) means there IS content evidence FOR "{survey_question}".
       * A single clear, explicit content-positive segment ("Isolated" pattern) is sufficient to call "{survey_question}" Present if content is unambiguous (e.g., "I want to die", "They are watching me").
       * "None" for content means there was no explicit speech evidence for "{survey_question}".
  
  5. Quick cross-segment verification (before final decision):
     - Re-scan all segments with "Yes" calls to ensure the described evidence is specific and observable (not vague).
     - Re-scan segments with "No" calls to confirm the relevant regions/content were actually observable (not just "Not visible"/"Cannot assess").
     - Identify ALL segments that contain clear positive evidence (behavioral and/or content) for "{survey_question}" and use one of the clearest as the anchor for "best_moment".
  
  6. Decide: Is "{survey_question}" Present or Not Present overall?
     - This is a BINARY decision: choose exactly one of "Present" or "Not Present".
     - You MUST set "mse_item_status" to "Present" if ANY segment shows clear positive evidence (behavioral and/or content) for "{survey_question}".
       * This includes Isolated, Intermittent, and Consistent behavioral_pattern_type/content_pattern_type.
     - You may only set "Not Present" if:
       * There is NO clear positive evidence for "{survey_question}" in ANY segment, AND
       * At least some relevant regions/content were directly observable and coded as "No" (not only "Not visible"/"Cannot assess").
     - Weigh speech content as primary for ideation/thought-content domains, while still integrating behavioral evidence.
     - For behavioral domains, emphasize the behavioral_pattern_type (None/Isolated/Intermittent/Consistent) and severity.
     - Do NOT let "Not visible" or "Cannot assess" segments dilute positive evidence; they are missing data, not negatives.
     - Briefly note whether affect/motor behavior and content are broadly congruent, incongruent, or mixed, and whether this strengthens or weakens the case.
  
  7. Determine the overall severity rating (Normal|Minimal|Mild|Moderate|Marked|Severe|Extreme),
     using both:
     - Intensity of signs in the most informative segments, and
     - Pattern type (e.g., "Consistent mild" vs "Isolated severe").
  
  8. Determine overall evidence strength:
     - "Weak"     = sparse or ambiguous evidence, substantial missing data.
     - "Moderate" = some clear evidence, but with meaningful limitations or mixed segments.
     - "Strong"   = multiple clear converging signs across segments with adequate visibility and intelligibility.
     - Base this on:
       - behavioral_pattern_type and content_pattern_type,
       - amount of missing data ("Not visible"/"Cannot assess"),
       - clarity of key examples,
       - degree of cross-modal congruence.
  
  9. Using {prior_visit_summary}, determine pattern temporality:
     - "Likely trait-like" = similar signs and severity across multiple visits.
     - "Likely state-like" = new emergence or clear change in signs vs prior visit.
     - "Unclear"           = insufficient or conflicting visit-level information.
  
  10. Write a 2-3 sentence EHR summary for the next visit focused ONLY on "{survey_question}":
      - State whether "{survey_question}" was Present or Not Present.
      - State whether behavioral evidence was None/Isolated/Intermittent/Consistent.
      - State whether content evidence was None/Isolated/Intermittent/Consistent.
      - State overall evidence strength, pattern_temporality, severity, and trajectory vs prior visit.
  
  NEVER consider other MSE domains – ONLY "{survey_question}".
  
  ============================================================
  OUTPUT FORMAT - JSON ONLY
  ============================================================
  
  RULES:
  - "mse_item_status" MUST be EXACTLY "Present" or EXACTLY "Not Present".
  - "best_moment" should include a segment number and an approximate time range (e.g., "Segment 2, 00:05-00:12").
  - Start with {{ and end with }}.
  - Output RAW JSON ONLY – no text before or after.
  
  {{
    "mse_item_assessed": "{survey_question}",
    "mse_item_status": "<Present|Not Present>",
    "micro_evidence_summary": "Subtle behavioral signs of {survey_question} across segments (with segment numbers and approximate time ranges, or 'None observed')",
    "macro_evidence_summary": "Obvious behavioral signs of {survey_question} across segments (with segment numbers and approximate time ranges, or 'None observed')",
    "content_evidence_summary": "Key patient statements across segments that support or refute {survey_question}, with segment numbers and approximate time ranges (or 'No relevant content expressed')",
    "behavioral_pattern_type": "None|Isolated|Intermittent|Consistent",
    "content_pattern_type": "None|Isolated|Intermittent|Consistent",
    "modality_congruence_summary": "Brief summary of whether affect/motor behavior and content were largely Congruent, Incongruent, or mixed/Unclear across segments",
    "best_moment": "Segment X, 00:XX-00:XX: WHILE [audio content], VIDEO showed [specific behavior] – strongest evidence for {survey_question}",
    "summary_for_next_visit_context": "2–3 sentence EHR note: '{survey_question}' was Present/Not Present. [Key VIDEO + AUDIO + CONTENT pattern, evidence strength, trait/state impression, severity, and trajectory.]",
    "evidence_strength": "Weak|Moderate|Strong",
    "pattern_temporality": "Likely trait-like|Likely state-like|Unclear",
    "rating": "Normal|Minimal|Mild|Moderate|Marked|Severe|Extreme",
    "trajectory": "Stable|Improving|Worsening"
  }}


decision_trace_prompt: |
  DECISION TRACE: "{survey_question}"
  
  **ONLY** focus on "{survey_question}" and **NOTHING ELSE**.
  
  INPUT:
  1) SEGMENT LOGS: {decision_trace_input}
  2) META-ANALYSIS: {final_diagnosis_text}
  3) PRIOR VISIT: {prior_visit_summary}
  
  TASK:
  Summarize the evidence for "{survey_question}" ONLY. Ignore all other MSE domains.
  You MUST include and integrate:
  - behavioral (micro/macro) evidence from VIDEO,
  - acoustic evidence from AUDIO, and
  - speech content (key patient statements, from audio+transcript).
  
  INTERNAL REASONING (DO NOT OUTPUT):
  - You MAY use <think>...</think> for internal chain-of-thought.
  - Do 2 internal passes:
    PASS A: Build a compact evidence table (segment → time window → modality → observation → supports/against).
    PASS B: Consistency audit: ensure quoted timestamps are specific, and ensure "Not Present" is not concluded from missing data.
  - Your final response MUST NOT include any <think> tags and MUST use the exact output headers/structure below.
  
  Your decision trace MUST clearly answer:
  - WHAT: What was the final decision for "{survey_question}" (Present vs Not Present, rating, trajectory),
          what were the behavioral and content pattern types (None/Isolated/Intermittent/Consistent),
          what was the overall evidence strength (Weak/Moderate/Strong), and whether the pattern appeared
          trait-like or state-like.
  - WHY: Why did you arrive at that decision? Which specific behavioral signs, cross-modal congruence patterns,
         and speech content support it? How did the pattern across segments (None/Isolated/Intermittent/Consistent)
         influence your call?
  - HOW: How was the decision made step-by-step? Which segments, timeframes, and modalities were inspected,
         how consistent the evidence was across segments (Isolated vs Intermittent vs Consistent), how you
         handled contextual normalization (culture, age, language, situation), and how you weighed conflicting
         or missing data (e.g., "Not visible" / "Cannot assess")?
  
  You MUST explicitly reference:
  - Segment numbers (e.g., "Segment 1", "Segment 3") and, when possible, approximate time ranges
    (e.g., "around 00:20–00:40") to show WHERE in the video/audio the key evidence comes from.
  - WHICH modality each piece of evidence comes from (VIDEO, AUDIO, CONTENT), and how they converged or diverged
    (congruent vs incongruent).
  - HOW you handled missing data (e.g., segments where feet were Not visible or speech was Cannot assess)
    to make sure the reader understands the limits of the decision.
  - WHETHER the behavioral evidence pattern was None/Isolated/Intermittent/Consistent and whether the content
    evidence pattern was None/Isolated/Intermittent/Consistent, and how that influenced the final call and the
    evidence_strength rating.
  - If you concluded "{survey_question}" is Present based on:
    * An isolated but severe behavioral segment, or
    * An isolated but clear content-positive segment,
    you MUST explicitly state this and justify why a single segment was sufficient, given the broader context.
  
  OUTPUT FORMAT (USE THESE EXACT HEADERS AND LABELED STRUCTURE):
  
  === DECISION TRACE SUMMARY ===
  
  FINAL STATUS: [MUST be exactly one of: "Present" OR "Not Present" - NO other values allowed]
  
  WHAT (structured breakdown):
  - Status: Present OR Not Present (MUST match FINAL STATUS above)
  - Rating: Normal|Minimal|Mild|Moderate|Marked|Severe|Extreme
  - Trajectory: Stable|Improving|Worsening
  - Behavioral Pattern: None|Isolated|Intermittent|Consistent
  - Content Pattern: None|Isolated|Intermittent|Consistent
  - Evidence Strength: Weak|Moderate|Strong
  - Pattern Temporality: Likely trait-like|Likely state-like|Unclear
  
  WHY (cite specific evidence with EXACT time ranges):
  [Explain WHY you reached that decision. You MUST cite specific time ranges within segments, NOT just segment numbers.
   CORRECT format: "In Segment 2, between 00:05–00:12, I observed..."
   INCORRECT format: "Segment 2 shows..." (too vague - segments can be 15-60+ seconds long)
   Reference VIDEO + AUDIO + CONTENT evidence with exact timestamps.]
  
  HOW (step-by-step process):
  [Explain HOW you made the decision: which segments and time windows you reviewed, how you integrated
   micro/macro behavior with speech content, how you handled Not visible/Cannot assess data, and how
   this led to your final ratings.]
  
  === EVIDENCE FOR "{survey_question}" ===
  
  MICRO SIGNS (with exact time ranges):
  [For each subtle behavioral sign, you MUST specify:
   - Segment number
   - EXACT time range within segment (e.g., "00:03-00:08" NOT just "early in segment")
   - Modality (VIDEO)
   - What was observed
   Example: "Segment 1, 00:07-00:14, VIDEO: fine hand tremor visible at rest for ~7 seconds"
   State whether micro signs were isolated, intermittent, or consistent across segments.]
  
  MACRO SIGNS (with exact time ranges):
  [For each obvious behavioral sign, you MUST specify:
   - Segment number
   - EXACT time range within segment
   - Modality (VIDEO/AUDIO)
   - What was observed
   Example: "Segment 3, 00:22-00:45, VIDEO: markedly slowed movements; AUDIO: 3-4 second response latencies"
   State whether macro signs were isolated, intermittent, or consistent.]
  
  CONTENT EVIDENCE (with exact time ranges):
  [For each key patient statement:
   - Segment number
   - EXACT time range when statement occurred
   - Quote or close paraphrase
   Example: "Segment 2, 00:31–00:38, CONTENT: 'I wish I wouldn't wake up' - passive suicidal ideation"
   State whether content was isolated, intermittent, or consistent.
   Note segments with "No relevant content" or "Cannot assess".]
  
  KEY MOMENT (single strongest evidence with exact time):
  [Format: "Segment X, 00:XX–00:XX: WHILE [exact quote or audio description], VIDEO showed [specific behavior]"
   This must be the single most compelling piece of evidence for "{survey_question}".]
  
  === FINAL DETERMINATION ===
  MSE Item: "{survey_question}"
  Status: [MUST be exactly "Present" OR "Not Present" - same as FINAL STATUS above]
  Rating: [Normal|Minimal|Mild|Moderate|Marked|Severe|Extreme]
  Trajectory: [Stable|Improving|Worsening vs prior visit, or "Baseline"]

final_summary_prompt: |
  FINAL PARAGRAPH: "{survey_question}"
  
  **ONLY** describe findings related to "{survey_question}" and **NOTHING ELSE**.
  
  MSE ITEM: {survey_question}
  RATING: {extracted_rating}
  META-ANALYSIS: {meta_text}
  PRIOR VISIT: {prior_visit_summary}
  
  OUTPUT:
  Write ONE paragraph in EHR style focused ONLY on "{survey_question}":
  
  "MSE Severity Rating: {extracted_rating}. The sign/symptom '{survey_question}' was [Present/Not Present].
  Behavioral evidence: [Briefly describe the micro and macro behavioral signs of {survey_question} observed, if any.
  Content evidence: Summarize key patient statements related to {survey_question} (or note if none were expressed).
  Key moment: WHILE [audio content], [visual + content] was observed.
  Trajectory: [Stable/Improving/Worsening] compared to prior visit.]"
  
  RULES:
  - Single paragraph only
  - Focus ONLY on "{survey_question}" - ignore all other MSE domains
  - Use concise clinical language
  - Implicitly reflect integration of video, audio, and content in the narrative.

mse_guide: |
  You are a highly experienced psychiatrist designing very specific observational questions for an
  AUDIO+VIDEO junior psychiatrist that performs a Mental State Examination (MSE). The junior psychiatrist
  will always have access to:
  - VIDEO (visible behavior),
  - AUDIO (voice and non-speech sounds), and
  - TEXT CONTENT (a transcript of what is said when available).

  Input MSE domain: "{survey_question}"

  Goal:
  Create a concise, head-to-toe checklist of very specific QUESTIONS the trainee psychiatrist should use
  while watching and listening to a single patient in this MSE domain, and reading the transcript when available.
  The questions should cover both:
  (a) observable behavior (HOW the patient looks and sounds), and
  (b) speech content (WHAT the patient actually says) when this is clinically relevant to "{survey_question}".

  ============================================================
  MANDATORY REASONING STEP (YOU MUST OUTPUT THIS):
  ============================================================
  You MUST output your reasoning process BEFORE the questions. Use this exact format:
  
  === REASONING START ===
  
  STEP 1 - DOMAIN UNDERSTANDING:
  [Think deeply about what "{survey_question}" means in clinical psychiatry]
  - Clinical definition: [your analysis]
  - Observable manifestations: [list them]
  - What distinguishes this from other MSE domains: [your analysis]
  
  STEP 2 - SIGN ENUMERATION:
  [List ALL possible observable signs (visual + auditory + content) that could indicate "{survey_question}"]
  Visual signs by body region:
  - Appearance/Grooming/Clothing (head-to-toe): [overall cleanliness; hair and facial hair care; visible skin hygiene [list signs]
  - Clothing cleanliness vs stains/soiling; wrinkles suggesting slept-in clothes; torn or ill-fitting garments [list signs] 
  - Missing or mismatched items (e.g., one shoe); seasonally/contextually inappropriate dress; costume-like, flamboyant [list signs]
  - Thematically odd outfits; prominent logos/slogans or symbols [list signs]
  - Head/Face/Eyes: [list signs]
  - Mouth/Jaw: [list signs]
  - Neck/Shoulders: [list signs]
  - Arms/Hands/Fingers: [list signs]
  - Torso/Posture: [list signs]
  - Legs/Feet: [list signs]
  Auditory signs (form):
  - Speech features: [list signs]
  - Non-speech sounds: [list signs]
  Behavioral/Interaction signs:
  - [list signs]
  Semantic content signs (WHAT the patient says) - ONLY IF clinically relevant:
  - If "{survey_question}" is an ideation/thought-content domain (e.g., suicidal ideation, homicidal ideation, delusions, hallucinations, obsessions/ruminations, pathological guilt, worthlessness), list content signs:
    - Themes in speech relevant to "{survey_question}" (use domain-specific themes; do NOT import unrelated examples)
    - Explicit statements that directly support "{survey_question}" (use domain-specific examples)
    - Self-referential statements and beliefs that map to "{survey_question}"
  - Otherwise, write: "Not clinically relevant for this domain; prioritize observable behavior and speech form."
  
  STEP 3 - CLINICAL FREQUENCY RANKING:
  [Rank the enumerated signs by how FREQUENTLY they appear in patients with "{survey_question}"]
  MOST COMMON (>70% of cases): [list signs]
  MODERATELY COMMON (40-70%): [list signs]
  LESS COMMON (15-40%): [list signs]
  RARE BUT PATHOGNOMONIC (<15%): [list signs]
  
  STEP 4 - FINAL SELECTION:
  [Based on frequency ranking, select the MOST clinically important signs to generate questions for.
   Prioritize the 10-30 most frequent and clinically informative signs, with the highest-probability signs first.]
  Selected signs (ordered by frequency): [numbered list]
  
  === REASONING END ===
  
  === QUESTIONS START ===
  [Now output ONLY the questions with inline severity anchors, ordered by frequency - most common first]
  ============================================================

  Coverage requirements:
  Your questions must:
  - Treat the MSE domain "{survey_question}" as the north star: most questions should target how THIS specific
    domain appears in observable behavior and, when appropriate, in speech content.
  - Be ORDERED BY CLINICAL FREQUENCY/PROBABILITY: Place questions about the MOST FREQUENTLY observed signs of "{survey_question}"
    FIRST, progressing to less common signs.
  - Look at the person holistically, from head to toe:
    - General appearance and grooming (clothing, cleanliness, appropriateness to context).
    - Facial expression and eyes.
    - Mouth, jaw, and speech-related movements.
    - Head, neck, shoulders.
    - Arms, hands, fingers.
    - Torso and posture.
    - Legs, feet, gait (if visible).
  - Consider how they talk (form):
    - Speech rate, volume, prosody, articulation, fluency, and pauses, etc.
  - Consider breathing and other non-speech sounds.
  - Consider interaction with the environment, other people, and the interviewer:
    - Use of objects, distance and orientation in space, interaction with other people or interviewer.
  - Consider WHAT they say (content) ONLY when "{survey_question}" is an ideation/thought-content domain:
    - Ask content questions ONLY about themes and explicit statements clearly related to "{survey_question}".
    - Do NOT import generic examples (death/harm/persecution/etc.) unless they are truly relevant to "{survey_question}".
  - Include both:
    - MACRO questions (about obvious, sustained patterns over time, e.g. 30-60 seconds or more).
    - MICRO questions (about brief, subtle, repetitive, or high-frequency events, e.g. within 5–10 seconds).
  - Prefer questions that sometimes combine what is seen (video), what is heard (audio), and what is read (transcript content)
    in the same observation.

  Severity anchors (stepped/laddered examples) – INLINE, SINGLE LINE:
  - For EVERY question you output, you MUST include three severity anchors on the SAME LINE as the question.
  - Use this exact inline pattern for every line:
    Core question || Mild example: ... || Moderate example: ... || Severe example: ...
  - Each anchor must be a concrete, observable description of how the sign might appear at that intensity,
    using only information available from video/audio (including the literal words spoken and their transcript).
  - The core question MUST still start the line and MUST begin with "Do", "Does", "Is", "Are", or "Can".

  Constraints:
  - Each OUTPUT LINE must follow this exact structure:
    [QUESTION starting with Do/Does/Is/Are/Can] || Mild example: [text] || Moderate example: [text] || Severe example: [text]
  - Example of a correctly formatted line:
    Does the patient avoid eye contact when answering questions? || Mild example: Occasionally looks away or down during some answers. || Moderate example: Frequently avoids eye contact during most answers, looking down or to the side. || Severe example: Consistently avoids eye contact throughout the interaction, never looking toward the interviewer.
  - Every line (question + anchors) must be concrete and answerable using only information ultimately available from
    video and audio. If a transcript is provided, it is simply another way of accessing the same speech content and
    should be used, but not treated as a separate modality from what was actually said.
  - Phrase each QUESTION so that an answer of "Yes" indicates observable EVIDENCE FOR an abnormal sign or symptom
    in the "{survey_question}" domain, and an answer of "No" means the behaviour/appearance/content is within
    normal limits.
  - Avoid questions where "Yes" describes normal or healthy behaviour (for example, instead of "Are the clothes
    clean?", use "Is there visible dirt or staining on any clothing item?").
  - Do not mention diagnoses, causes, or internal mental states.
  - Do not explain your reasoning in the output.
  - Do not add headings, labels, bullets, numbering, or prefixes such as "Question 1:".

  Output format (must always be respected):
  - Return ONLY single-line questions with their inline severity anchors.
  - A QUESTION line consists of:
    - 1 core QUESTION (starting with Do/Does/Is/Are/Can),
    - followed by " || Mild example: ... || Moderate example: ... || Severe example: ..."
  - Aim for 10-30 QUESTIONS (i.e., 10-30 lines). Minimum of 10.
  - CRITICAL: Questions must be ORDERED BY FREQUENCY/PROBABILITY - most commonly observed signs
    of "{survey_question}" FIRST, progressing to less common signs.
  - One QUESTION per line (with its anchors inline on the same line).
  - No extra text before or after the questions, EXCEPT you MAY optionally include a final line: "=== QUESTIONS END ===".
  - Do not include summaries, explanations, or commentary of any kind.
  - Even if the input is vague or unusual, you must still output only such one-line questions with anchors, guided by
    the MSE domain above.
```
